# Germline variants associated with immunotherapy-related adverse events

**DOI:** 10.1101/2022.04.10.22273627

**Authors:** Stefan Groha, Sarah Abou Alaiwi, Wenxin Xu, Vivek Naranbhai, Amin H. Nassar, Ziad Bakouny, Elio Adib, Pier V. Nuzzo, Andrew L. Schmidt, Chris Labaki, Talal El Zarif, Biagio Ricciuti, Joao Victor Alessi, David A. Braun, Sachet A. Shukla, Tanya E. Keenan, Eliezer Van Allen, Mark M. Awad, Michael Manos, Osama Rahma, Leyre Zubiri, Alexandra-Chloe Villani, Christian Hammer, Zia Khan, Kerry Reynolds, Yevgeniy Semenov, Deborah Schrag, Kenneth L. Kehl, Matthew L. Freedman, Toni K. Choueiri, Alexander Gusev

**Affiliations:** Division of Population Sciences, Department of Medical Oncology, Dana-Farber Cancer Institute, Boston, MA, USA; Broad Institute of Harvard & MIT, Cambridge, MA, USA; Lank Center for Genitourinary Oncology, Dana-Farber Cancer Institute, Boston, MA, USA; Department of Medicine, Brigham and Women’s Hospital, Boston, MA, USA; Department of Medical Oncology, Dana-Farber Cancer Institute, Harvard Medical School, Boston, MA, USA; Department of Internal Medicine and Medical Specialties (DIMI), School of Medicine, University of Genoa, Genoa, Italy; Translational Immunogenomics Lab, Dana-Farber Cancer Institute, Boston, MA, USA; Lowe Center for Thoracic Oncology, Dana-Farber Cancer Institute, Boston, MA, USA; Breast Oncology Program, Dana-Farber/Brigham and Women’s Cancer Center, Boston, MA, USA; Center for Cancer Precision Medicine, Dana-Farber Cancer Institute, Boston, MA, USA; Massachusetts General Hospital, Boston, MA, USA; Center for Immunology and Inflammatory Diseases, Department of Medicine, Massachusetts General Hospital, Boston, MA, USA; Harvard Medical School, Boston, MA, USA; Division of Medical Oncology, Bartlett, Massachusetts General Hospital, Boston, MA, USA; Department of Dermatology, Massachusetts General Hospital, Boston, MA, USA; Division of Genetics, Brigham and Women’s Hospital, Boston, MA, USA; Genentech, South San Francisco, CA, USA

## Abstract

Immune checkpoint inhibitors (ICIs) have yielded remarkable responses in patients across multiple cancer types, but often lead to immune related adverse events (irAEs). Although a germline cause for irAEs has been hypothesized, no systematic genome wide association study (GWAS) has been performed and no individual variants associated with the overall likelihood of developing irAEs have yet been identified. We carried out a Genome-Wide Association Study (GWAS) of 1,751 patients on ICIs across 12 cancer types, with replication in an independent cohort of 196 patients and independent clinical trial data from 2275 patients. We investigated two irAE phenotypes: (i) high-grade (3-5) events defined through manual curation and (ii) all detectable events (including high-grade) defined through electronic health record (EHR) diagnosis followed by manual confirmation. We identified three genome-wide significant associations (p<5×10^−8^) in the discovery cohort associated with all-grade irAEs: rs16906115 near *IL7* (combined p=1.6×10^−11^; hazard ratio (HR)=2.1), rs75824728 near *IL22RA1* (combined p=6.6×10^−9^; HR=1.9), and rs113861051 on 4p15 (combined p=1.3×10^−8^, HR=2.0); with rs16906115 replicating in two independent studies. The association near *IL7* colocalized with the gain of a novel cryptic exon for *IL7*, a critical regulator of lymphocyte homeostasis. Patients carrying the *IL7* germline variant exhibited significantly increased lymphocyte stability after ICI initiation than non-carriers, and this stability was predictive of downstream irAEs and improved survival.

## Introduction

Cancer immunotherapy has revolutionized cancer care by harnessing the patient’s own immune system against tumors^1^. However, because immune checkpoint inhibitors (ICIs) block the body’s natural safeguards that prevent immune overactivation, treatment can also affect non-malignant tissues and cause autoimmune-like side effects^2–5^. Patients on ICIs thus commonly experience immune-related adverse events (irAEs)^4,6,7^. High-grade irAEs can lead to hospitalization and treatment cessation in 15-30% of patients^7^, emphasizing the urgent need to understand mechanisms and predictors of irAEs. Recent studies have also shown that irAEs correlate with positive anticancer responses^8^, highlighting their relevance to broader therapy outcomes.

One hypothesis for the heterogeneity in irAE onset and severity is the impact of germline genetic determinants of immune activity^6^. Recent work has shown that polygenic germline risk for autoimmune conditions is correlated with onset of cutaneous and thyroid irAEs^9,10^. Prior studies of response to ICIs have also highlighted both individual germline HLA alleles^11^ and MHC diversity^12^ as predictors of overall survival. However, no individual genetic variants associated with irAEs or response have so far been established. In this work, we hypothesized that individual germline variants may influence the broad spectrum of irAEs by modulating the general excitability of the immune system, as recently observed for somatic alterations^13,14^. We carried out a Genome-Wide Association Study (GWAS) of irAEs for patients on ICIs at a single institution, followed by replication in patients treated at an independent institution and on clinical trials.

## Results

### Genome-wide Association Study of irAEs

We carried out a GWAS for two irAE phenotypes in 1,751 patients of European ancestry across 12 cancer types treated with ICIs at a single tertiary institution (DFCI cohort, Table 1, Figure S2). Two irAE outcomes were defined for each patient following treatment initiation: (1) “high-grade” irAEs (259 cases, 1375 controls) determined by manual curation of records following NCI CTCAE (v5) guidelines for grade 3-5 events, with attribution of AEs as being immune-related determined based on the clinical consensus of the patient’s care team; (2) “all-grade” irAEs (339 cases, 1412 controls) algorithmically identified based on autoimmune-like EHR diagnosis codes (Supplementary Table S7) and including any high-grade irAEs, followed by manual review to exclude any events that were definitively linked to other causes. Detailed chart review in a subset of 44 patients found 85% of “all-grade” irAEs to be consistent with grade 2 or higher events (see Methods, Supplementary Table S10).

**Table 1.**

| Profile cohort (discovery) |  | MGH cohort (replication) |  |
| --- | --- | --- | --- |
|  | Overall<br>(N=1751) |  | Overall<br>(N=196) |
| **irAEs** |  | **irAEs** |  |
| Yes | 339 (19.4%) | Yes | 33 (16.8%) |
| No | 1412 (80.6%) | No | 163 (83.2%) |
| **Cancer Type** |  | **Cancer Type** |  |
| Non-Small Cell Lung Cancer | 539 (30.8%) | GU | 31 (15.8%) |
| Melanoma | 241 (13.8%) | Melanoma | 44 (22.4%) |
| Other | 236 (13.5%) | Other | 72 (36.7%) |
| Glioma | 112 (6.4%) | Thoracic | 49 (25.0%) |
| Breast Carcinoma | 111 (6.3%) |  |  |
| Esophagogastric Carcinoma | 111 (6.3%) |  |  |
| Renal Cell Carcinoma | 109 (6.2%) |  |  |
| Bladder Cancer | 94 (5.4%) |  |  |
| Head and Neck Carcinoma | 90 (5.1%) |  |  |
| Ovarian Cancer | 40 (2.3%) |  |  |
| Cancer of Unknown Primary | 34 (1.9%) |  |  |
| Colorectal Cancer | 34 (1.9%) |  |  |
| **Sex** |  | **Sex** |  |
| Female | 814 (46.5%) | Female | 82 (41.8%) |
| Male | 937 (53.5%) | Male | 114 (58.2%) |
| **Age** |  | **Age** |  |
| Mean (SD) | 63.0 (12.4) | Mean (SD) | 64.2 (13.4) |
| Median [Min, Max] | 63.9 [19.6, 102] | Median [Min, Max] | 66.6 [22.3, 90.2] |
| **Type of Treatment** |  | **Type of Treatment** |  |
| CTLA4 | 49 (2.8%) | CTLA4 | 17 (8.7%) |
| Combination Therapy | 154 (8.8%) | Combination Therapy | 16 (8.2%) |
| PD-1/PD-L1 | 1548 (88.4%) | PD-1/PD-L1 | 163 (83.2%) |
| **Sequencing** |  |  |  |
| Prior to ICI initiation | 1363 (77.8%) |  |  |
| Following ICI initiation | 388 (22.2%) |  |  |
| **Start year** |  |  |  |
| Before 2016 | 357 (20.4%) |  |  |
| 2016 | 416 (23.8%) |  |  |
| 2017 | 557 (31.8%) |  |  |
| 2018 | 305 (17.4%) |  |  |
| After 2018 | 116 (6.6%) |  |  |

Three genome-wide significant loci (p<5×10^−8^) were associated with all-grade irAEs: one near Interleukin 7 (*IL7*) at chr8q21, one near the Interleukin 22 Receptor Subunit Alpha 1 (*IL22RA1*) at chr1p36, and the third association at chr4p15 (Figure 1, Figure S3). No genome-wide significant associations were identified for high-grade irAEs. We tested each SNP for association with individual irAE subtypes and found that all three SNPs were nominally significant across multiple irAE subtypes with no clear outliers (Figure S13, Table S8), and were significant in the 80% of patients on PD-1 ICIs (with insufficient power to test differences by drug class; Figure S5). Neither variant was associated with overall survival nor with death without irAEs, even though all all-grade irAEs were associated with longer overall survival in a time-dependent analysis (HR=0.78 [0.65-0.94], p=8.6×10^−3^; Table S6), consistent with previous findings.

**Figure 1.**
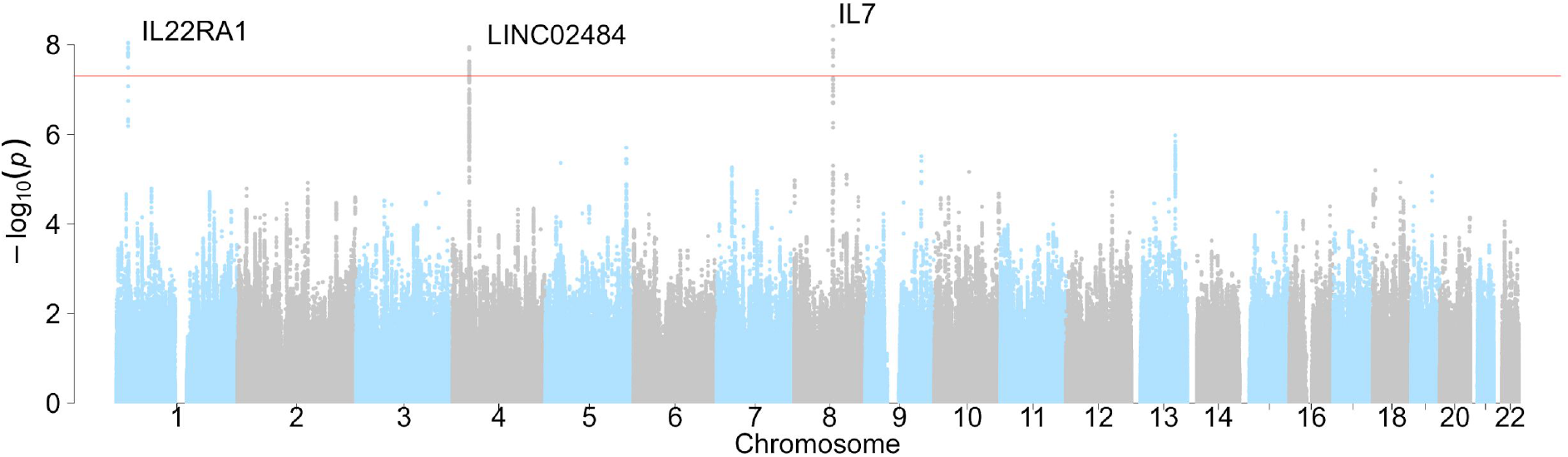
Manhattan plot of GWAS associations. Associations in the DFCI discovery cohort for all-grade irAEs. Each dot represents and associated SNP, with position of the SNP (x-axis) and p-value of the association (y-axis, -log10 scale).

The lead 8q21 SNP was rs16906115, a common variant in an intron of *IL7*, with a hazard ratio HR=2.0 [1.6-2.5] (p=3.8×10^−9^, HR corrected for imputation error, see Supplementary Note; Figure 2, Figure S7). Within individual cancer types, a consistent sign was observed in 9 out of 11 cancer types (p=2.7×10^−2^ by a one-sided binomial test) with nominal significance (p<0.05) in Non-Small Cell Lung Cancer, Melanoma, RCC, Bladder Cancer, Cancer of Unknown Primary, as well as the collection of “other” less common cancer types (Figure 2). The lead 1p36 SNP was rs75824728, a common variant in an intron of *IL22RA1*, with a hazard ratio HR=1.9 [1.5-2.4] (p=8.4×10^−9^; Figure S4). This SNP was also nominally significantly associated with high-grade irAEs with a comparable effect size (HR=1.5 [1.1-2.0], p=0.015; Figure S9). Within individual cancer types, the association was nominally significant in Non-Small Cell Lung Cancer, Melanoma, Breast Cancer, as well as the collection of “other” less common cancer types (Figure 2). The lead 4p15 SNP was rs113861051 with a hazard ratio HR= 2.0 [1.6-2.6] (p=1.1×10^−8^) (Figure S4). We carried out a broad scan for germline, clinical, and somatic features (including tumor mutational burden) associated with irAEs or interacting with the identified SNPs but observed no significant associations after multiple test correction (Table S2), underscoring the contribution of these germline findings to irAEs.

**Figure 2.**
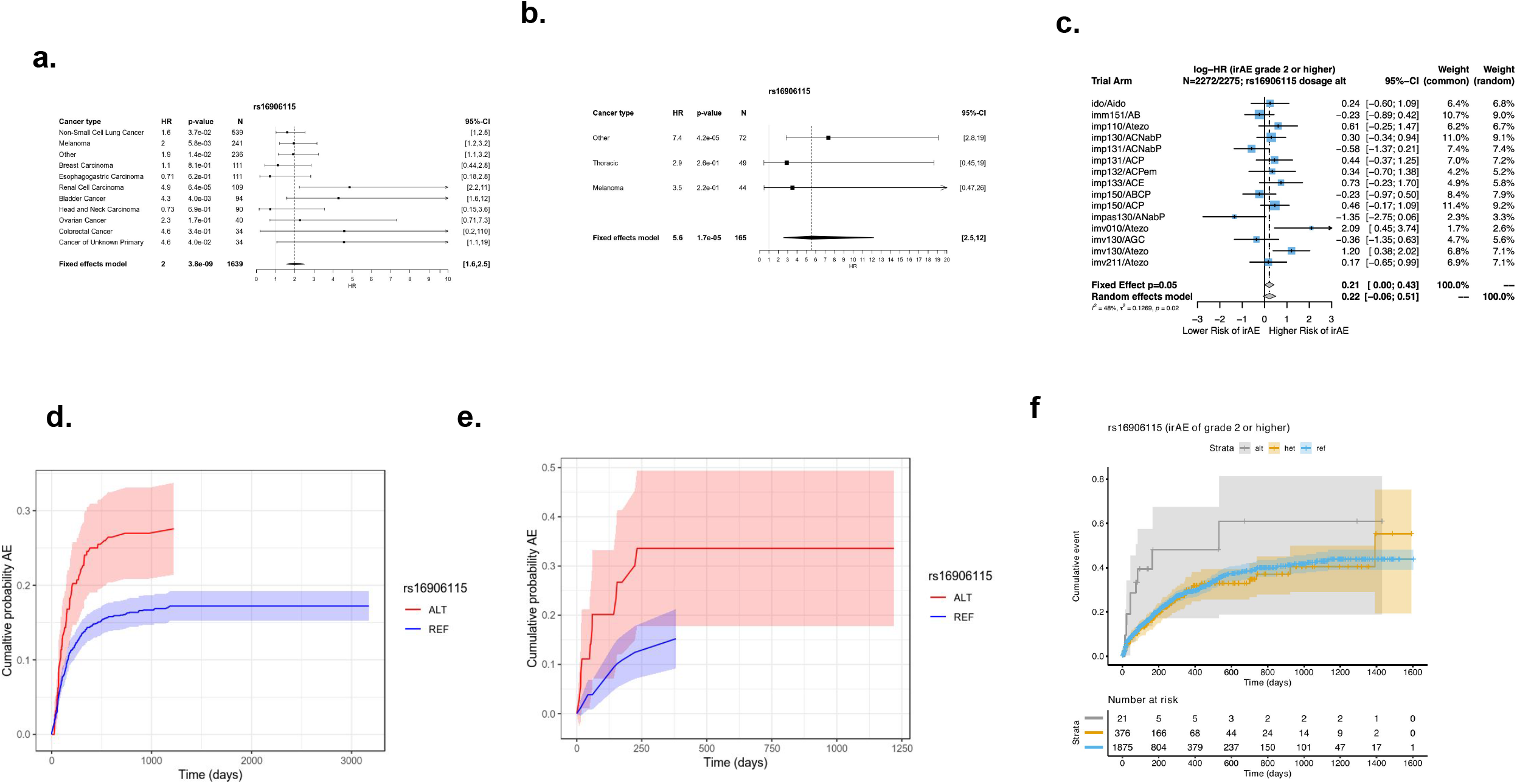
Discovery associations and replication in MGH and CT cohort. Forrest plot of genome-wide significant association (reference dosage) with all-grade irAEs at 8q21 (**a**) in the Profile cohort, (**b**) the MGH cohort and (**c**) in the CT cohort. Aalen-Johansen estimator for the cumulative incidence of adverse events following ICI initiation stratified on SNP dosage in the DFCI discovery cohort (**d**), in the MGH replication cohort (**e**) and using a Kaplan Meier estimator in the CT cohort (**f**).

We evaluated potential modifiers or interactions of the discovered associations. First, using a normative cohort of >23,000 non-ICI patients at DFCI, no significant association between any of the three SNPs and the time from sequencing to the first code-based “all-grade” event was observed (Figure S10), indicating that the SNP effects were highly specific to the ICI setting. Likewise, none of the three lead SNPs were significantly associated with prior autoimmune disease defined based on ICD codes, nor with a polygenic risk score (PRS) for autoimmune disease (see Supplementary Methods) either in the ICI cohort or in the non-ICI patients. Though ICD-based autoimmune disease definitions are likely to be incomplete, we verified that our prior autoimmune disease phenotype was significantly associated with irAEs (Figure S2) and germline polygenic risk for autoimmune disease (p=8.8×10^−4^ in the ICI cohort, see Supplementary Note). Finally, we investigated various adjustments for the competing risk of death, immortal time bias, as well as inclusion/exclusion of individuals with immune-related diagnoses at the start of treatment and observed no significant impact on these associations (Supplementary Note, Figure S6).

### Independent replication of the *IL7* variant

We evaluated the three discovery SNPs in two independent cohorts for replication (see Methods for cohort details). The rs16906115 variant near *IL7* replicated significantly (HR=3.6 [1.8-7.1], p=2.8×10^−4^) in an independent cohort of 196 patients on ICIs treated at Mass General Hospital (MGH cohort) with severe irAEs requiring hospitalization and confirmed by chart review (Figure 2, Figure S11, Figure S12). rs16906115 also replicated nominally (HR=1.2 [1.0-1.5], p=0.05) in a second cohort of 2,275 patients on clinical trials (CT cohort) for ICIs with grade 2-5 irAEs recorded as part the trial (Figure 2c). Although no significant outliers were observed, a test for heterogeneity of effect sizes across trials was nominally significant (p=0.02), primarily driven by the IMpassion130 triple-negative breast cancer study. Sub-analyses did not show significant associations with any other event grade (Figure S15, S16, S17) or irAE subtype (Figure S15, S18). The other two associations, rs75824728 near IL22RA1 and rs113861051 at 4p15, did not replicate in either independent cohorts; although all three associations remained significant in a meta-analysis with the MGH cohort (due to data constraints, we could not perform a genome-wide meta-analysis with the CT cohort). Lastly, while this manuscript was in preparation, the variant near *IL7* was independently replicated in a third cohort of 214 melanoma patients on ICIs in the UK with severe (grade 3 or above) irAEs requiring corticosteroids, which was further molecularly characterized in parallel work^42^. Thus, the *IL7* associated variant replicated in three independent cohorts (Supplementary Table S5).

### Colocalization of *IL7* GWAS variant with a novel *IL7* cryptic exon

We sought to identify a putative mechanism for the *IL7* locus by integrating our GWAS with molecular data. In tissue-specific expression quantitative trait loci (eQTLs) mapped by the GTEx consortium^20^, the lead irAE SNP was significantly associated with *IL7* exon junction usage in testis for the chr8:78740082:78749524 junction (which we call *IL7*_junc_) and had an R^2^ of 0.98 to the lead *IL7*_junc_ QTL (rs7816685), which was also in the irAE GWAS credible set (Supplementary Table S3, Supplementary Figure S8) (Figure S20). By inspection of the raw RNA-seq coverage and junction plots, we observed that carriers of the risk allele exhibited splicing and activation of a novel 250bp cryptic exon (spanning chr8:78746500-78746750, which we call *IL7*_ce_ for “cryptic exon”) that was entirely absent from all homozygous non-carriers (Figure 3a). The SNP had a stronger effect on *IL7*_ce_ and explained the association with *IL7*_junc_ in a conditional analysis, consistent with *IL7*_ce_ being the causal mediator (Figure S20). The lead *IL7*_ce_ QTL, rs7816685, was the only QTL located in the splice region of *IL7*_ce_ and was predicted to be -1bp from an Acceptor Gain region for *IL7* (delta score = 0.19 by SpliceAI^27^) further implicating rs7816685 as the likely causal variant. Despite the increased expression in Testis, we did not find any association between sex and irAEs (Figure S25) nor any sex interaction (p=0.28), and we hypothesize that the effect was observed in Testis incidentally as this tissue contributes disproportionately to both cis and trans eQTLs identified in GTEx^20^.

**Figure 3.**
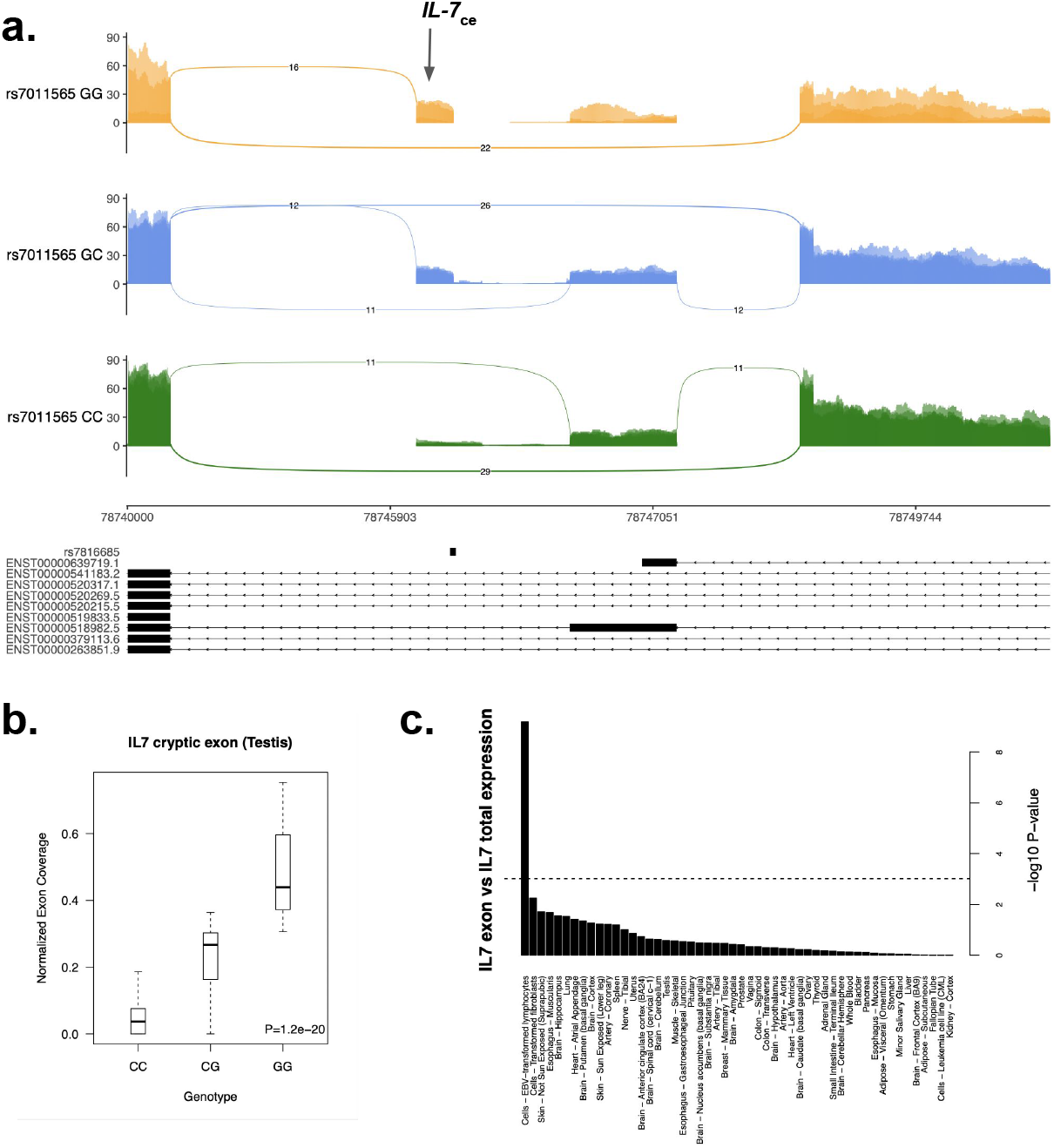
Colocalization with IL7 cryptic exon. Sashimi plot of alternative splicing of *IL7* stratified on the lead splice QTL (**a**), with the putative causal variant shown below and the cryptic exon highlighted (IL7_ce_). Cryptic exon activity stratified by lead splice QTL genotype (**b**). Significance of co-expression of *IL7* and *IL7*_ce_ across GTEx tissues (Pearson correlation) (**c**).

Considering *IL7*_ce_ as the putative functional mechanism, we next quantified its activity in a broader set of tissues and cell-types. Across the GTEx tissues, *IL7*_ce_ expression was generally low, with Testis and Lymphoblastoid Cell Lines (LCLs) exhibiting clear high outlier expression (Figure S21), the latter consistent with the role of *IL7* in lymphoid cell development. LCLs uniquely exhibited significant correlation between *IL7*_ce_ and total *IL7* expression (Figure 3c) as well as significantly higher *IL7:IL7R* co-expression in the presence of *IL7*_ce_ (P=3.4×10^−3^; Figure S21), suggesting that *IL7*_ce_ may stabilize *IL7* expression or increase *IL7R* binding in lymphocytes. To better understand the precise cell-type of action, we mapped *IL7*_ce_ in publicly available RNA-seq from sorted immune-related cells from patients with autoimmune diseases: *IL7*_ce_ was highly expressed in B-cells and moderately expressed in CD4 T-cells, with no observable expression in the other immune cell types (Figure S22). Across 11,284 tumors from TCGA, *IL7*_ce_ was associated with 13/61 previously defined immune landscape features^14^ (Bonferroni corrected p<0.05 after adjusting for cancer type and overall IL7 expression; Table S1) including: lymphocyte infiltration, BCR/TCR diversity, IFN-gamma response, and the “Wound Healing” immune cluster. In parallel work, the B cell specific effect of rs16906115 on *IL7* was confirmed in melanoma patients receiving ICIs and its influence on T cell development further characterized^42^.

### Association of *IL7* variant with lymphocyte homeostasis

Due to the known role of *IL7* in lymphocyte homeostasis^28^, we explored whether the influence of rs16906115 on irAEs was reflected in peripheral blood lymphocyte count from clinical laboratory data. As a surrogate for lymphocyte expansion/homeostasis, we defined the change in lymphocyte count 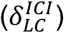 using measurements 30 days before/after ICI initiation for patients in the DFCI and MGH cohorts. In the DFCI cohort, carriers of the risk allele exhibited no significant change in lymphocytes (median 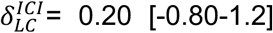, p=0.69) whereas non-carriers had significantly reduced 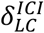 (median 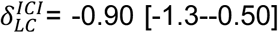, p=2.3×10^−6^ by paired Wilcoxon test); which replicated in the MGH cohort (median 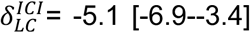 p=4.1×10^−8^ for non-carriers; median 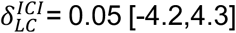, p=0.99 for carriers). The difference in 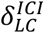 between carriers and non-carriers was significant in both the Profile cohort (difference in mean 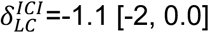, p=0.040) and the MGH cohort (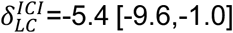, p=0.017; Figure 4), as well as the independent analysis of melanoma patients receiving immune checkpoint inhibitors^42^ Similarly, 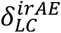 defined 30 days before vs. after irAEs was stable for carriers (p=0.49) but not for non-carriers (p=2.2×10^−3^), though this association may be complicated by steroid usage (Figure S19). The *IL7* variant thus had a consistent stabilizing effect on lymphocyte counts at the initiation of ICI therapy and at the onset of irAE. Results were similar when using absolute lymphocyte count (Supplementary Note). Lastly, we investigated whether this phenomenon pointed to broader lymphocyte dynamics irrespective of genotype status. Indeed, higher 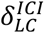 was nominally associated with increased irAE incidence (HR=1.2 per s.d., p=0.018) and a concomitant increase in overall survival for those patients not experiencing any irAE (HR=0.87, p=1.6×10^−3^) in the DFCI cohort.

**Figure 4.**
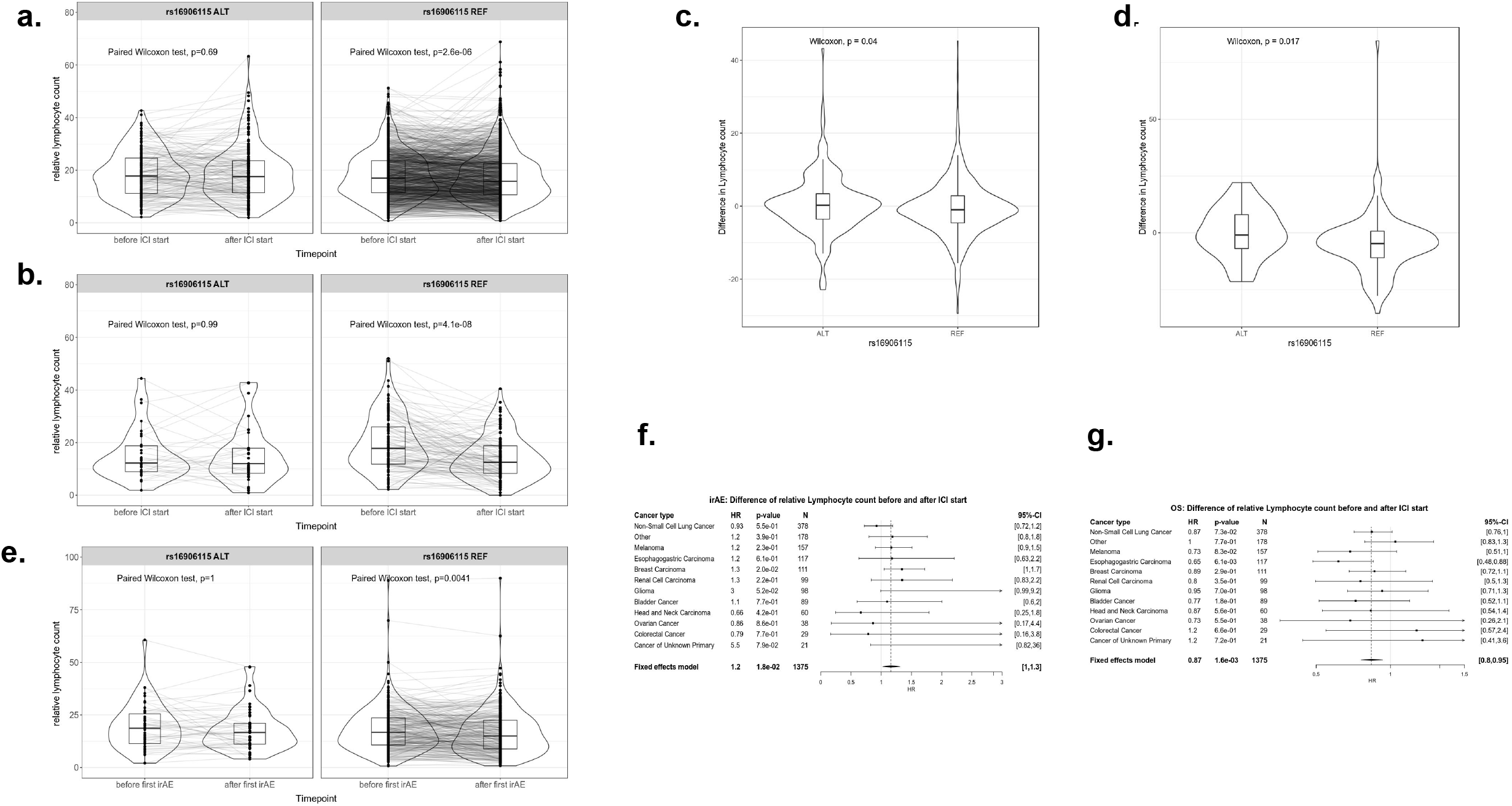
Lymphocyte counts up to 30 days before and after ICI initiation for cases and controls. Paired Wilcoxon test between time-points in carriers and non-carriers in the DFCI (a) and MGH (b) cohort. Wilcoxon test of the difference in lymphocyte counts prior versus following ICI initiation between carrier and non-carrier in the Profile (c) and MGH cohort (d). Paired Wilcoxon test between before and after first irAE in carriers and non-carriers in the DFCI cohort (e). Association between difference in lymphocyte counts before and following ICI initiation and developing and irAE (f) as well as death without an irAE (g).

## Discussion

We conducted a GWAS of irAEs in an observational pan-cancer setting, identifying three novel genome-wide significant associations, with replication of a variant near *IL7* in three independent cohorts. For the *IL7* locus, we nominated rs7816685 as the likely causal variant, residing in the splice junction of a novel cryptic exon of *IL7* and associated with broad differences in the tumor immune landscape.

Although the putative *IL7* mechanism identified in this work has not previously been linked to irAEs, *IL7* has been extensively studied for its involvement in immune response and auto-immune disease. *IL7* has a critical role in the development and maturation of T cells, limits organ toxicity during antiviral immune response, and supports aberrant immune activity in autoimmune disease^29^. There is evidence that *IL7* expression blocks PD-1, leading to type 1 diabetes^30^, as well as involvement in the development of chronic colitis^31^; functioning like a natural checkpoint inhibitor^32^. The administration of *IL7* in patients with cancer results in increased lymphocyte counts (particularly CD4+ and CD8+ T-cell counts) and reduced regulatory T-cell counts^28^. It is therefore plausible that the *IL7* risk variant results in a more facilitatory milieu for autoimmune/autoreactive immune responses in patients on ICIs, explaining its association with irAEs. Several studies have shown that *IL7* receptor blockade can reverse autoimmune response^30,33^, offering a potential therapeutic avenue for managing *IL7* mediated irAEs. These findings motivate further investigation into the influence of the *IL7* SNP and *IL7* splicing on ICI response, particularly for combination treatments.

Our study has several limitations. First, the heterogeneity of irAE presentation and severity led us to define two, partially overlapping outcomes. In the discovery GWAS, irAEs were manually abstracted from clinical notes as well as algorithmically inferred using EHR data (followed by manual quality control), and may have thus included some events with ambiguous causes, especially for irAEs that were observed well after the treatment was administered. This heterogeneity was highlighted by different associations with downstream survival, where DFCI all-grade irAEs were protective, DFCI high-grade irAEs were not associated with survival, and MGH high-grade irAEs were hazardous. The observational nature of the DFCI/MGH populations also limited the homogeneity of the cohort. Although we attempted to control for common covariates, most patients had a complex treatment history that could not be modelled. However, we expect this heterogeneity to primarily influence power and generalizability, as germline genetic variation cannot be *caused* by unmodelled confounders. Second, the influence of irAEs in risk allele carriers on downstream treatment decisions and the potential for germline-guided “decision support” is of great interest. While we attempted to annotate treatment discontinuation and steroid administration in carriers of the *IL7* risk allele (see Methods and Supplementary Table S9) we could not draw clear conclusions due to the limited data and high baseline rates of both outcomes. Ideally this relationship could be investigated in a prospective follow-up with strict monitoring of. Clinical decisions. Third, we restricted our study to individuals of European ancestry to mitigate possible population stratification, but further studies in non-European are warranted to understand the generalizability of these associations. In particular, the associated variant rs7816685 near IL7 has an allele frequency of 0.31 in East Asian populations (compared to 0.065 in Europeans) and may thus explain more variance in irAEs in Asian patients. Lastly, the use of imputation from tumor-only panel sequencing for the discovery GWAS produced imputed variants with more noise than direct genotyping and likely excluded some difficult-to-impute or rare polymorphisms. This limitation also offers an opportunity for further analysis of this variant in existing panel sequencing datasets^34^.

The identification of genetic variants associated with irAEs is consistent with a hypothesized patient-specific immunological set point and opens avenues for future analysis to inform the genetic architecture of irAEs including: genetic correlation with other complex traits^35^, polygenic risk scores for patient stratification^36^, and Mendelian Randomization to estimate the causal influence of irAEs on other cancer outcomes^37^. Larger studies will enable polygenic heritability analyses to uncover the cell types, gene sets, and pathways that drive these outcomes. Ultimately, the utility of these associations to identify high-risk patients for early monitoring or treatment modifications must be evaluated in prospective, randomized trials in conjunction with their influence on anti-tumor response.

## Methods

### Cohort definition, consent, and genotyping

Analyses were carried out across three cohorts with genotyping and clinical information (additional genotyping and phenotyping information provided in the Supplementary Note):

#### Dana-Farber Cancer Institute (DFCI) cohort

1,751 patients of European ancestry (to avoid any confounding from population stratification) treated with ICIs (90% with PD-1/PD-L1) at DFCI from 2013 to 2020 (Table 1), across 12 cancer types. Patients were biopsied and sequenced on the OncoPanel tumor sequencing platform^15^ targeting 275-447 cancer genes and germline single nucleotide polymorphisms (SNPs) were imputed using ultra low-coverage off-target reads^16^ with imputation accuracy evaluated using a partially overlapping set of directly genotyped individuals (Figure S1). For normative comparisons, a pan-cancer control cohort of 23,763 individuals treated with non-ICI therapies at DFCI was similarly sequenced and imputed through the same pipeline. Patients provided informed consent for research and IRB approval was obtained (protocol #19-033 and #19-025).

#### MGH cohort

An independent pan-cancer cohort of 196 patients on ICIs at Massachusetts General Hospital (MGH) with direct germline genotyping on the Illumina Multi-Ethnic Genotyping Array (MEGA) (Table 1). Occurrence of high-grade irAEs (33 cases, 163 controls) was obtained through the Severe Immunotherapy Complications Program at MGH, for inpatient management of high-grade irAEs. Each high-grade irAE was clinically confirmed by an oncology team with expertise in diagnosing and managing irAEs and secondarily verified by organ-specific clinical irAE experts at the corresponding disease center. Secondary analyses of previously collected data were performed with approval from the Partners IRB (IRB protocol 2020P002307).

#### Clinical Trial (CT) replication cohort

A second replication analysis of individual associations was carried out in 2275 patients that were treated with atezolizumab (anti-PD-L1) and were of European ancestry and met sample and genetic data quality control from 12 previously published clinical trials sponsored by F. Hoffmann–La Roche/Genentech (Table S4). Studies included trials of atezolizumab in renal cell carcinoma (IMmotion, imm), lung cancer (IMpower, imp), triple-negative breast cancer (IMpassion, impas), urothelial cancer (IMvigor, imv), and advanced cancers (IDO; majority lung, breast, or ovarian). All patients provided informed consent for the respective main study. A subset of patients signed an optional Research Biosample Repository (RBR) Informed Consent Form (ICF) to provide whole blood samples for future research, including study of inherited and non-inherited genetic variation from these whole blood samples. Ethics Committees (EC) and Institutional Review Boards (IRB) at each study site for each clinical trial approved the clinical trial protocol, the main study ICF, and the RBR ICF. Whole-genome sequencing data was collected from whole blood (as previously described^9^) and used to compute individual variant association statistics.

### Statistical analysis

GWAS was carried out across all variants in the DFCI and MGH cohorts for association with time to irAE separately for each irAE definition. In all cohorts, individuals were restricted to European ancestry. Due to the competing risk of death while on treatment, a cause-specific hazard rate was computed for every SNP using a mixed-effects model^17^, equivalent to censoring on death or loss-to-follow-up. In each cohort, covariates were included for ancestry, age, gender, line/type of treatment (Supplementary Note). Statistical fine-mapping of genome-wide significant loci was carried out using the SuSIE software^18^. irAE probabilities and cumulative incidence were quantified using the Aalen–Johansen estimator, a non-parametric estimator that accounts for competing risks^19^. Associations between irAEs and overall survival were evaluated using a time-dependent covariate coded as 0 for controls and as 1 starting from the time of first irAE.

### Analysis of molecular data

Associations were functionally characterized using publicly available gene expression and splicing data from multiple resources. Variants were connected to putative target genes using gene expression and splicing QTLs across 44 tissues from the GTEx consortium^20^. RNA-seq BAM files were downloaded from the GTEx repository and splice junction usage was analyzed using ggsashimi^21^. Cell sorted data across 6 immune cell subsets from individuals with autoimmune diseases and healthy controls were accessed from ref.^22^ and GEO (SRP045500). Pan-cancer RNA-seq BAM files from TCGA were used to quantify expression across tumor sites^23^ and correlated against previously defined immune populations and signals^14^. Analyses of read-level activity and cryptic splicing were carried out using the recount2 framework^24^. Clinical lab measurements were extracted from EHR data via the Oncology Data Retrieval System^25^ framework for the DFCI cohort and the Research Patient Data Registry (RPDR)^26^ for the MGH cohort.

## Data Availability

Summary statistics of the GWAS will be made available upon publication.

## Disclosures

D.A.B. reports nonfinancial support from Bristol Myers Squibb, honoraria from LM Education/Exchange Services, and personal fees from MDedge, Exelixis, Octane Global, Defined Health, Dedham Group, Adept Field Solutions, Slingshot Insights, Blueprint Partnerships, Charles River Associates, Trinity Group, and Insight Strategy, outside of the submitted work.

K.K. reports receiving honoraria from IBM and Roche.

M.M.A reports grants and personal fees from Genentech, grants and personal fees from Bristol-Myers Squibb, personal fees from Merck, grants and personal fees from AstraZeneca, grants from Lilly, personal fees from Maverick, personal fees from Blueprint Medicine, personal fees from Syndax, personal fees from Ariad, personal fees from Nektar, personal fees from Gritstone, personal fees from ArcherDX, personal fees from Mirati, personal fees from NextCure, personal fees from Novartis, personal fees from EMD Serono, personal fees from Panvaxal/NovaRx, outside the submitted work.

O.R. reports research support from Merck. Speaker for activities supported by educational grants from BMS and Merck. Consultant for Merck, Celgene, Five Prime, GSK, Bayer, Roche/Genentech, Puretech, Imvax, Sobi, Boehringer Ingelheim. Patent “Methods of using pembrolizumab and trebananib” pending.

S.A.S. reports nonfinancial support from Bristol-Myers Squibb, and equity in Agenus Inc., Agios Pharmaceuticals, Breakbio Corp., Bristol-Myers Squibb and Lumos Pharma.

T.K.C. reports research/advisory boards/consultancy/Honorarium (Institutional and personal, paid and unpaid): AstraZeneca, Aveo, Bayer, Bristol Myers-Squibb, Eisai, EMD Serono, Exelixis, GlaxoSmithKline, IQVA, Ipsen, Kanaph, Lilly, Merck, Nikang, Novartis, Pfizer, Roche, Sanofi/Aventis, Takeda, Tempest. Travel, accommodations, expenses, medical writing in relation to consulting, advisory roles, or honoraria. Stock options: Pionyr, Tempest. Other: Up-to-Date royalties, CME-related events (e.g.: OncLIve, PVI, MJH Life Sciences) honorarium. NCI GU Steering Committee. Patents filed, royalties or other intellectual properties (No income as of current date): related to biomarkers of immune checkpoint blockers and ctDNA. No speaker’s bureau.

Z.B. reports research support from the imCORE Network on behalf of Genentech, Inc. and Bristol-Myers Squibb. Honoraria from UpToDate.

## Acknowledgements

We thank all the patients who consented to participate in this study as well as the institutional data collection efforts that made this study possible. We would like to acknowledge Michael Hassett, Neil Lindeman, David Liu, Pieter Lukasse, Laura MacConnaill, Paz Polak, Scott Rodig, Noah Zaitlen, and Elad Ziv for helpful discussions; the DFCI Oncology Data Retrieval System (OncDRS) for the aggregation, management, and delivery of the clinical and operational research data used in this project; and the DFCI/BWH Data Sharing Group for the aggregation, management, and delivery of the clinical and genomics data used in this project.

AG is supported by NIH R01CA227237, NIH R01CA244569, NIH R21HG010748, the Claudia Adams Barr Foundation, the Louis B. Mayer Foundation, and the Doris Duke Charitable Foundation. SAS acknowledges support by the NCI (R50RCA211482). SG was supported by NIH R01CA227237 and a DFCI Trustee Fellowship. TKC is supported in part by the Dana-Farber/Harvard Cancer Center Kidney SPORE and Program, the Kohlberg Chair at Harvard Medical School and the Trust Family, Michael Brigham, and Loker Pinard Funds for Kidney Cancer Research at DFCI. TEK acknowledges grant support from the National Institutes of Health (T32CA009172).

## Supplementary Note

### Sample collection, genotype imputation, and quality control

#### DFCI cohort

The DFCI cohort was sequenced as part of the *Profile* project, a prospective clinical sequencing effort for consented patients undergoing routine treatment at the Dana-Farber Cancer Institute and affiliated hospitals. A custom targeted hybrid capture sequencing platform (OncoPanel) was used to assay genomic variation from tumor biopsies. Each sample was sequenced on one of three panel versions targeting the exons of 275, 300, and 447 genes respectively. Samples meet a minimum of 30X coverage for 80% of targets for analysis. Somatic variation (including single nucleotide variants, insertions/deletions, and copy number variation) was called by the Profile clinical bioinformatics pipeline and signed out by a pathologist at Brigham & Women’s Hospital after technical review, as previously described^15^. Off-target and on-target reads from the sequenced BAMs were imputed using the STITCH imputation software^16,38^. Imputed variants were restricted to minor allele frequency >1% and imputation INFO score >0.4. Genetic ancestry was inferred using principal component projection with the SNPWEIGHTS software^39^. Continental components were used to exclude non-European individuals, and within-Europe components were included as covariates.

A partly overlapping cohort of 833 individuals (126 overlapping patients on ICIs) with both OncoPanel tumor sequencing and direct germline SNP array genotyping (on the Illumina Multi-Ethnic Genotyping Array (MEGA)) was used to benchmark the imputation accuracy. Pearson correlation for each SNP was computed between the tumor-imputed and germline genotyped individuals. Mean imputed SNP correlation was 0.86 after variant quality control and highly uniform across the genome (Figure S1). Detailed analysis of variant imputation accuracy have been described separately and the imputation workflow is publicly available^16^. For visualizations where imputed patients were stratified by variant carrier/non-carrier status, the decision boundary was determined using logistic regression of carrier status on imputed dosage in the samples with both tumor sequencing and SNP array data.

#### MGH cohort

Blood samples were collected from MGH patients and genotyped on the Illumina MEGA array. Data was imputed to the 1000 Genomes reference panel using the Haplotype Reference Consortium imputation server, followed by quality control removing variants with minor allele frequency <1% and INFO score <0.9. Genetic ancestry was inferred using in-sample principal components and restricted to Europeans.

#### CT cohort

A subset of patients signed an optional Research Biosample Repository (RBR) Informed Consent Form (ICF) to provide whole blood samples for future research, including study of inherited and non-inherited genetic variation from these whole blood samples. Whole-genome sequencing data was collected from whole blood as previously described^9^. Genetic ancestry was inferred using ADMIXTURE and restricted to Europeans (ancestry >0.7). In-sample principal components were also computed to account for any remaining population structure.

### Outcome definitions in the DFCI cohort

Mortality was collected using linkage to the National Death Index (NDI) through 2019. For patients who died after 2019, a clinical death index from the electronic health record (EHR) was used (which captured 86% of occurred deaths when evaluated for patients before 12/31/2019).

The “all-grade” event definition was obtained by algorithmic abstraction using EHR diagnosis codes. A list of predefined relevant diagnosis codes was used to filter all available codes for potential adverse events after treatment start and up to 60 days after receiving the last ICI dose. Diagnosis codes, which were present in the EHR of the respective patient before treatment start were excluded. Evident false positives were excluded by inspection of the diagnosis code and manual review of the patient chart at the event date, to exclude events that did not occur or were clearly linked to non-ICI causes. The used search terms and manual exclusion list of search terms is shown in Supplementary table S7.

### Prior autoimmune disease and polygenic risk score

We investigated relationships between the identified risk variants and prior autoimmune disease and autoimmune disease risk. We defined patients with prior autoimmune disease based on the occurrence of an autoimmune related ICD10 code before ICI treatment start. Each irAE lead SNP was then tested for association with prior autoimmune disease, while adjusting for age, gender, treatment year, panel version of the sequencing panel, treatment type, line of treatment, as well as cancer type. As an alternative measure of autoimmune disease risk, we also inferred a polygenic risk score (PRS) for any autoimmune disease from a recent UK Biobank GWAS study (see Data Availability). We confirmed that the PRS was significantly associated with the previous ICD-based autoimmune disease definition in the ICI cohort (p=8.8×10^−4^). Each irAE lead SNP was again tested for association with the PRS, adjusting for cancer type, age, gender, panel version, as well as the first two principal components to control for ancestry.

### Termination of treatment and steroid administration

For a subset of 44 patients, which were selected based on highest dosage of the lead IL7 SNP, information on continuation of treatment after irAE as well as steroid administration was manually annotated through chart review.

### Survival analysis

#### GWAS discovery

In the DFCI discovery cohort, discovery of GWAS variants associated with risk of irAEs was performed using a multivariate multi-state survival framework modelling with irAE as the primary outcome and death as a competing risk. Direct modelling of competing risks is important for incidence computation and to account for potential survivor bias^40^, where individuals who live longer may develop more irAEs by chance. Due to computational constraints, the mixed-effects survival GWAS methodology did not allow for stratified covariates and flexible truncation. We thus re-estimated the top associations (p<5×10^−6^) by fixed-effect meta-analysis over the cancer types with stratification of any covariates that exhibited a proportional hazards violation. Lastly, to account for error in the imputation, we rescaled the HR based on the imputed/genotyped relationship, though we note this is a linear rescaling that does not impact the significance of the association.

Additionally to account for immortal time bias, 422 patients who were sequenced after the start of ICI treatment were left-truncated until sequencing. Left-truncation and excluding patients with allograft surgery or immunosuppressants at treatment start did not influence any of the genome-wide significant associations (Figure S6).

In the replication cohorts (MGH and CT), cause-specific hazard ratios and p-values were estimated by conventional survival analysis with censoring on death or loss to follow-up. This cause specific hazard computation (our primary measure of effect-size) is equivalent to that estimated from the multi-state model.

#### Multi-state modelling of competing risks

We employed a time-to-event analysis with irAEs as the event of interest. However, as death precludes from experiencing an irAE, death events were addressed through an illness-death model, a special case of the class of multi-state survival models. In this model patients in the “Treatment” state can either experience a transition to “irAE” or to “Death” without having experienced an irAE. Furthermore, patients who have experienced an irAE can also transition to the “Death” state. For any transition in the multi-state survival model censoring due to loss to follow-up, as well as left-truncation due to delayed sequencing was employed.

In the setting of multi-state survival models, there are two possible hazard rates one might be interested in: the cause-specific hazard and the subdistribution hazard. While the subdistribution hazard quantifies the risk for the incidence of the event in the population, the cause-specific hazard quantifies the inherent risk of a patient experiencing an event conditioned on that patient being event-free. The cause-specific hazard rate, therefore, corresponds to the infinitesimal generator of transitions in a Markov Jump process with added censoring. As we are interested in the biological mechanism of experiencing an irAE, the primary quantity of interest is the cause-specific hazard rate (see further discussion in ref.^41^). The subdistribution hazard, which takes into account the risk of the competing death event given from the same covariate, is of secondary interest primarily from an epidemiological perspective.

To address the challenge of estimating the cumulative population-level incidence/probability of an irAE in the multi-state setting, we employed the Aalen-Johansen estimator^19^. We treated irAEs as a transient state to obtain the probability over time to have experienced an irAE but be alive, and irAEs as an absorbing state to obtain the cumulative incidence of irAEs over time.

### Covariate adjustment

In the DFCI discovery cohort, covariates were included for: two within-Europe ancestry components (after restricting to European individuals, see above); age at treatment start; gender; line of treatment as determined from the EHR medication records; start year of treatment; type of treatment (PD1/PD-L1 or CTLA4 monotherapy, combination); concurrent alternate treatment (chemotherapy, targeted therapy); as well as two technical covariates adjusting for the version of the targeted panel and an indicator for sequencing after treatment start. Patients were grouped into cancer types with >30 individuals, and the analyses were stratified or meta-analyzed over cancer types (as indicated). In the MGH cohort, covariates were included for: cancer type, type of immune checkpoint inhibitor, age at treatment start, gender, and genetic ancestry. Cancer type was included as a covariate rather than a stratifying variable due to the relatively small sample size of each type and the assumption that common covariate effects could be better learned across all samples. In the CT cohort, covariates were included for five genetic ancestry components, and stratified on treatment arms (which also capture cancer types).

### Data availability

Full summary association statistics for the discovery cohort will be made available upon publication.

UK Biobank association statistics for autoimmune disease were previously computed by BOLT-LMM v2.3 and used to estimate the autoimmune disease PRS (accessed from: https://data.broadinstitute.org/alkesgroup/UKBB/UKBB_409K/). RNA-seq data from GTEx and TCGA was accessed through the Receount2 interface and API (https://jhubiostatistics.shinyapps.io/recount/).

## Figure Legends

**Supplementary Figure S1**. Distribution of SNP imputation accuracy (Pearson correlation) from panel sequencing for 833 samples with available direct germline genotyping.

**Supplementary Figure S2**. Cumulative number of patients experiencing high-grade irAEs (**a**) or all-grade irAEs (**b**) stratified by therapy class.

**Supplementary Figure S3**. QQ-Plot for all-grade adverse events.

**Supplementary Figure S4**. Discovery associations with rs75824728 (**a**) and rs113861051 (**b**) stratified by cancer type.

**Supplementary Figure S5**. Discovery associations with (a) rs16906115, (b) rs75824728 and (c) rs113861051 stratified by treatment class.

**Supplementary Figure S6**. Association of rs16906115 (**a**) without left-truncation of the patients and (**b**) excluding patients with previous allograft or on immunosuppressant drugs.

**Supplementary Figure S7**. Correlation to true genotype for the genome-wide significant SNPs rs16906115 (**a**) and rs75824728 (**b**) as well as rs113861051 (**c**) association in 833 patients where both panel and array sequencing was available.

**Supplementary Figure S8**. Finemapped 95% credible set of associations with at the 8q21 (rs16906115) locus.

**Supplementary Figure S9**. Forrest plot for the top all-grade irAE SNP association in 1p36 locus, tested against high-grade irAE definition.

**Supplementary Figure S10**. Incidence of all-grade irAEs over time for the DFCI ICI cohort (**a**) compared to incidence of the corresponding diagnosis codes in the DFCI non-ICI cohort (**b**).

**Supplementary Figure S11**. (**a**) Logarithmic hazard rates (effect sizes) and (**b**) p-values for association in the discovery DFCI cohort and the MGH cohort for the 8q21 locus, restricted to nominally significant associations in the discovery cohort (p<0.05). (**c**) Comparison of the association strengths of variants around the top association locus in DFCI and MGH. The 95% credible set in the DFCI cohort is colored in blue. The upper red line signifies genome wide significance, the lower red line bonferroni corrected significance for SNPs tested in the MGH cohort.

**Supplementary Figure S12**. Aalen-Johansen estimator for the probability of adverse events following ICI initiation taking into account death (**a**), as well as cumulative incidence of irAEs following ICI initiation (**b**) in the DFCI cohort; and in the MGH cohort (**c**,**d**).

**Supplementary Figure S13**. Association of r(a) rs16906115, (b) rs75824728 and (c) rs113861051 by type of irAE in the Profile cohort.

**Supplementary Figure S14**. Association of rs16906115 by type of irAE in (a) Profile and (b) CT cohort.

**Supplementary Figure S15**. Recorded irAE event type distribution in CT cohort for (a) grade >=1, (b) grade >=2 and (c) grade >=3.

**Supplementary Figure S16**. Cumulative irAE probability distributions for in CT cohort grade >=1, grade >=2 and grade >=3 irAEs for carriers and non-carriers of the (a) rs16906115, (b) rs75824728 and (c) rs113861051 reference allele.

**Supplementary Figure S17**. Forest plots over trial arms in CT cohort for association with grade >=1, grade >=2 and grade >=3 irAEs for carriers of the (a) rs16906115, (b) rs75824728 and (c) rs113861051 reference allele on the hazard scale.

**Supplementary Figure S18**. Association with irAE subtypes in CT cohort for grade >=1, grade >=2 and grade >=3 irAEs for carriers of the (a) rs16906115, (b) rs75824728 and (c) rs113861051 reference allele on the log-hazard scale.

**Supplementary Figure S19**. As steroids influence lymphocyte counts, we conditioned on the patient receiving steroids 30 days before or after irAE. We observed a significant effect for carriers when steroids were given (p=0.012) with no effect in non-carriers (p=0.84). There was no significant effect in either carriers (p=0.23) nor non-carriers (p=0.63) when no steroids were given.

**Supplementary Figure S20**. Association of 8q21 SNP with (**a**) IL7_junc_ and (**b**) IL7_ce_ in GTEx testis. The risk SNP was significantly associated with *IL7*_ce_ (P=1.2×10^−20^, R^2^=0.63) and remained significant after conditioning on *IL7*_junc_ (P_cond_=4.6×10^−15^), whereas the risk SNP had a much weaker association with *IL7*_junc_ (P=1.4×10^−9^, R^2^=0.11) that was no longer significant after conditioning on *IL7*_ce_ (P_cond_=0.05).

**Supplementary Figure S21**. Expression of *IL7ce* in GTEx tissues (**a**). *IL7-IL7R* co-expression as function of *IL7ce* in GTEx LCLs (**b**).

**Supplementary Figure S22**. Expression of *IL7*_ce_ in cell sorted immune cell data.

**Supplementary Figure S23**. *IL7*_ce_ expression in TCGA by site.

**Supplementary Figure S24**. Association between previous auto-immune disease as defined by auto-immune ICD-10 codes, and all-grade irAEs.

**Supplementary Figure S25**. Association between sex assigned at birth and irAEs.

**Supplementary Table S1**. Association of *IL7*_ce_ expression with TCGA immune landscape features. TGF-beta response, TCR diversity, BCR diversity, and proliferation were not significantly associated with overall *IL7* expression but were significant for *IL7*_ce_. In contrast, increases in Th1 and Th2 cells were highly significantly associated with total *IL7* expression (P<10^−30^) but not with *IL7*_ce_ (P>0.3).

**Supplementary Table S2**. Hazard ratios and significance of interaction term in interaction analysis of germline, somatic and clinical features with 8q21 SNP and with irAE as outcome.

**Supplementary Table S3**. 95% credible set of fine-mapping the 8q21 locus.

**Supplementary Table S4**. Names and references for the clinical trial studies.

**Supplementary Table S5**. List of genome-wide significant loci in the discovery cohort with effect sizes and p-values in the replication cohort, as well as allele frequency of the reference SNP in both Profile and MGH cohorts.

**Supplementary Table S6**. Overall survival association of adverse events in the Profile and MGH cohort.

**Supplementary Table S7**. Search terms and exclusion criteria for “all-grade”, EHR based irAE outcome definition

**Supplementary Table S8**. Frequency of irAE for carriers and non-carriers of the risk alleles by type of irAE.

**Supplementary Table S9**. Frequency of termination of therapy and steroid administration after irAE in the carrier and non-carrier group. The patients were ascertained by highest dosage of rs16906115 for curation.

**Supplementary Table S10**. Frequency and number of grades of irAEs. The patients were ascertained by highest dosage of rs16906115 for curation.

## Notes

### Author Declarations

Ethics committee/IRB of Dana Farber Cancer Institute gave ethical approval for this work.

### Summary of Updates

Light formatting changes

