## Supplemental Materials for "Germline variants associated with immunotherapy-related adverse events"

### Supplemental Material

### Figure S1

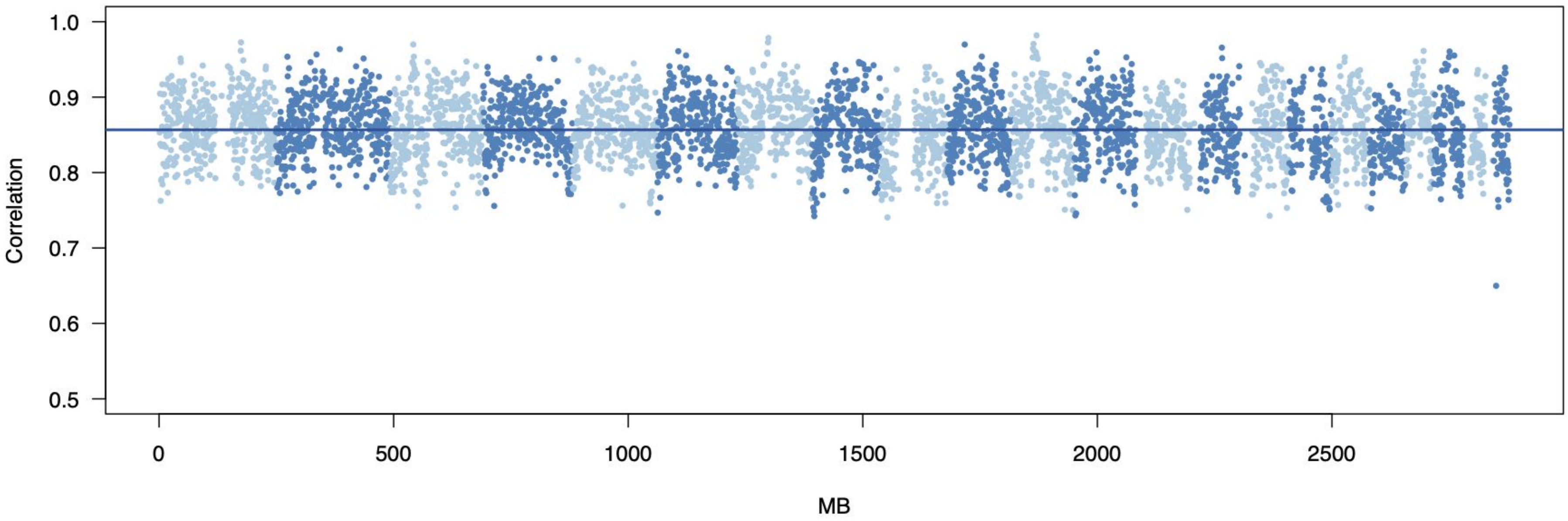

### Figure S2

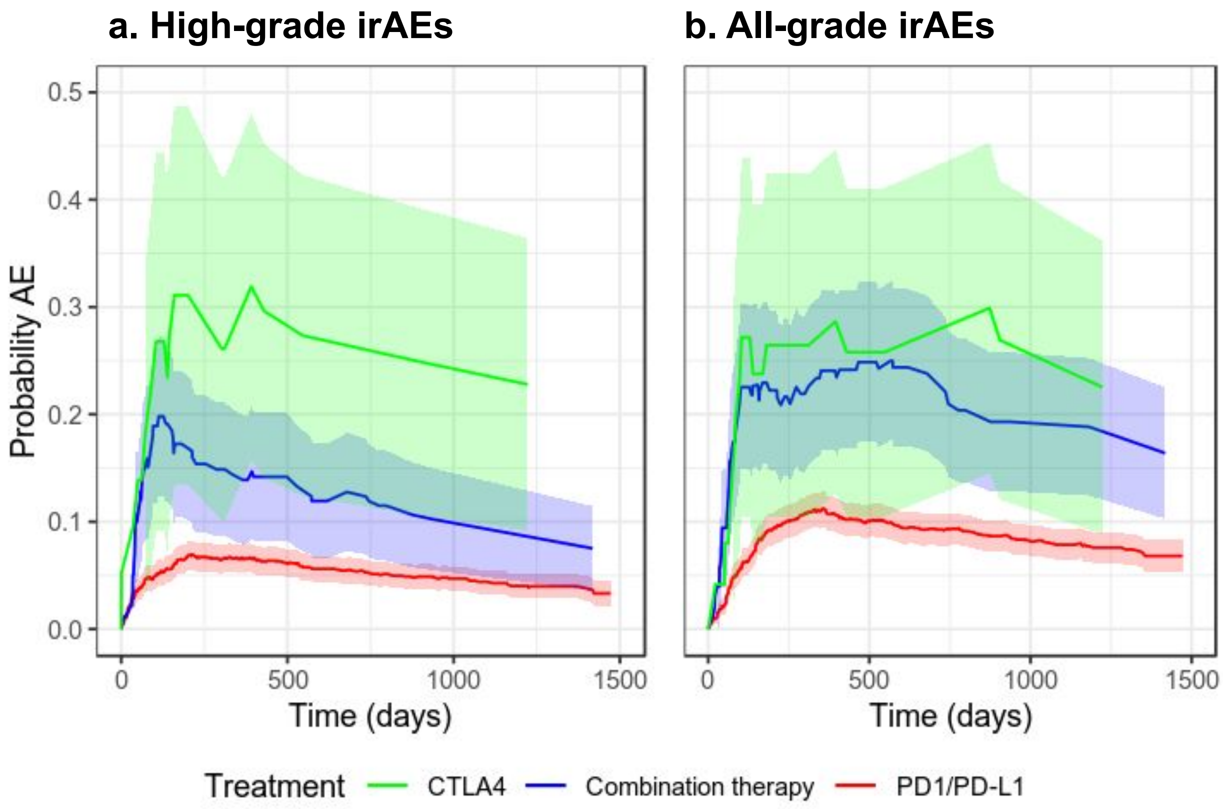
a. High-grade irAEs
b. All-grade irAEs
Probability AE
0.0
0.1
0.2
0.3
0.4
0.5
0
500
1000
1500
Time (days)
Time (days)
Treatment
CTLA4
Combination therapy
PD1/PD-L1

### Figure S3

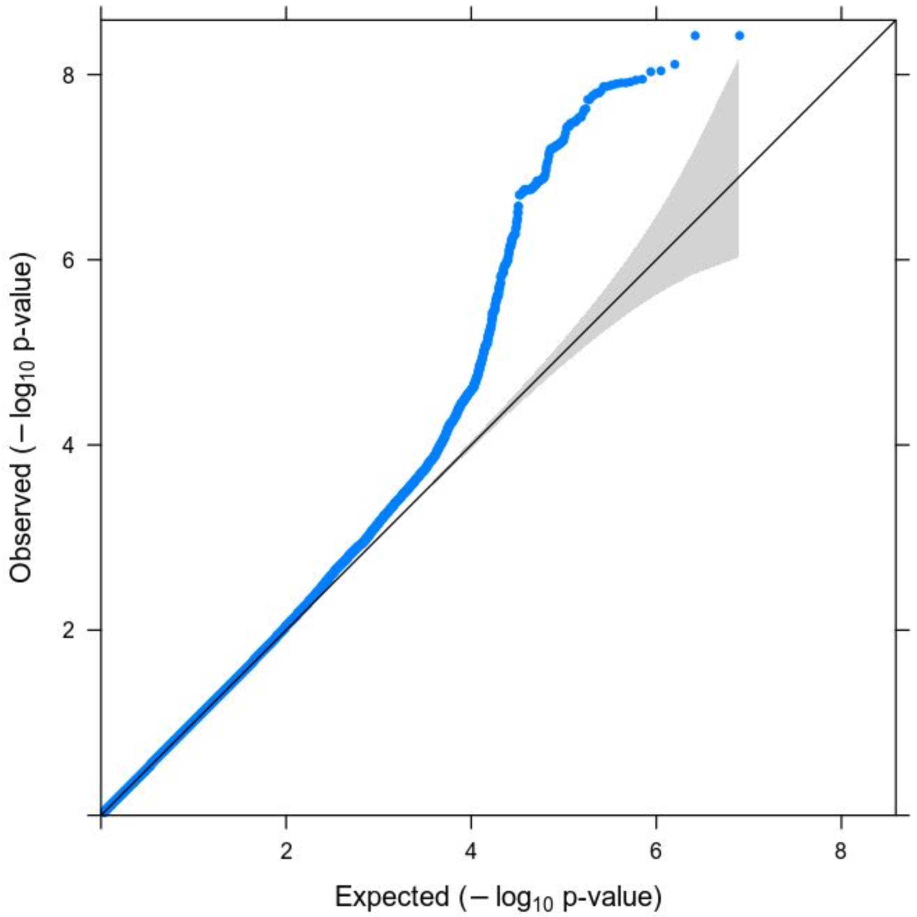

### Figure S4

**a.**

rs75824728

| Cancer type | HR | p-value | N | 95%-CI |
|---|---|---|---|---|
| Non-Small Cell Lung Cancer | 1.9 | 1.4e-03 | 539 | [1.3,2.9] |
| Melanoma | 1.6 | 3.1e-02 | 241 | [1,2.5] |
| Other | 2.5 | 3.3e-04 | 236 | [1.5,4.2] |
| Glioma | 4.1e-05 | 2.7e-01 | 112 | [5.9e-13,2900] |
| Breast Carcinoma | 2.6 | 1.4e-02 | 111 | [1.2,5.4] |
| Esophagogastric Carcinoma | 2.1 | 1.0e-01 | 111 | [0.87,4.9] |
| Renal Cell Carcinoma | 0.86 | 8.0e-01 | 109 | [0.28,2.7] |
| Bladder Cancer | 0.23 | 2.3e-01 | 94 | [0.021,2.5] |
| Head and Neck Carcinoma | 1.8 | 3.0e-01 | 90 | [0.58,5.8] |
| Colorectal Cancer | 60 | 4.9e-02 | 34 | [1,3500] |
| Cancer of Unknown Primary | 1.7 | 5.3e-01 | 34 | [0.32,9] |
| **Fixed effects model** | **1.9** | **8.4e-09** | **1711** | **[1.5,2.4]** |

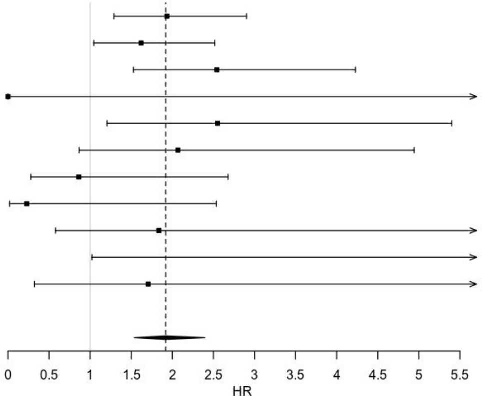


**b.**

rs113861051

| Cancer type | HR | p-value | N | 95%-CI |
|---|---|---|---|---|
| Non-Small Cell Lung Cancer | 1.8 | 2.9e-03 | 539 | [1.2,2.7] |
| Melanoma | 2.1 | 8.1e-04 | 241 | [1.4,3.3] |
| Other | 0.65 | 5.2e-01 | 236 | [0.18,2.4] |
| Glioma | 0.026 | 6.3e-01 | 112 | [1.2e-08,58000] |
| Breast Carcinoma | 4.7 | 3.2e-04 | 111 | [2,11] |
| Esophagogastric Carcinoma | 0.35 | 5.1e-01 | 111 | [0.016,7.7] |
| Renal Cell Carcinoma | 2.1 | 1.5e-01 | 109 | [0.77,5.7] |
| Bladder Cancer | 1.1 | 9.3e-01 | 94 | [0.19,6.3] |
| Head and Neck Carcinoma | 3.2 | 3.7e-02 | 90 | [1.1,9.3] |
| Ovarian Cancer | 3.5 | 7.3e-02 | 40 | [0.89,14] |
| Colorectal Cancer | 11 | 1.5e-01 | 34 | [0.41,290] |
| Cancer of Unknown Primary | 0.36 | 3.2e-01 | 34 | [0.048,2.7] |
| **Fixed effects model** | **2** | **1.1e-08** | **1751** | **[1.6,2.6]** |

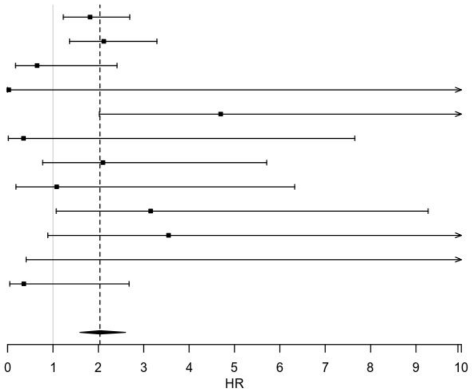

### Figure S5

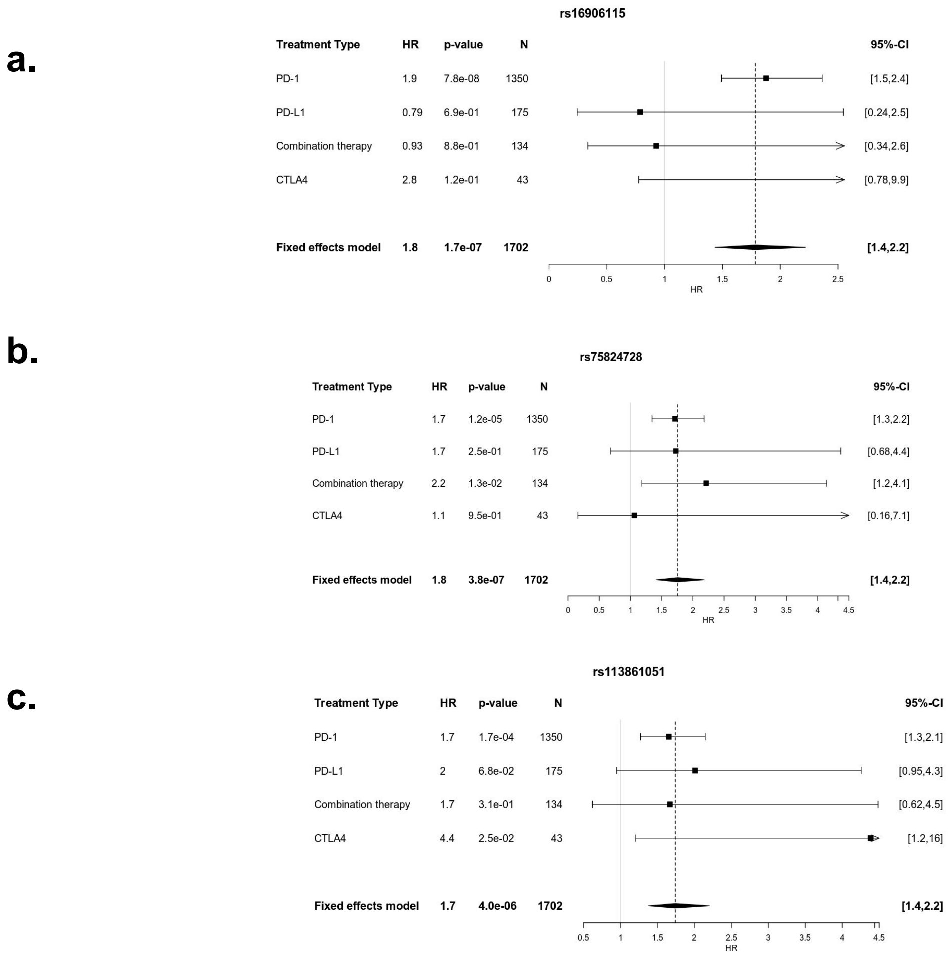
a.
rs16906115
Treatment Type HR p-value N 95%-CI
PD-1 1.9 7.8e-08 1350 [1.5,2.4]
PD-L1 0.79 6.9e-01 175 [0.24,2.5]
Combination therapy 0.93 8.8e-01 134 [0.34,2.6]
CTLA4 2.8 1.2e-01 43 [0.78,9.9]
Fixed effects model 1.8 1.7e-07 1702 [1.4,2.2]
0 0.5 1 1.5 2 2.5
HR
b.
rs75824728
Treatment Type HR p-value N 95%-CI
PD-1 1.7 1.2e-05 1350 [1.3,2.2]
PD-L1 1.7 2.5e-01 175 [0.68,4.4]
Combination therapy 2.2 1.3e-02 134 [1.2,4.1]
CTLA4 1.1 9.5e-01 43 [0.16,7.1]
Fixed effects model 1.8 3.8e-07 1702 [1.4,2.2]
0 0.5 1 1.5 2 2.5 3 3.5 4 4.5
HR
c.
rs113861051
Treatment Type HR p-value N 95%-CI
PD-1 1.7 1.7e-04 1350 [1.3,2.1]
PD-L1 2 6.8e-02 175 [0.95,4.3]
Combination therapy 1.7 3.1e-01 134 [0.62,4.5]
CTLA4 4.4 2.5e-02 43 [1.2,16]
Fixed effects model 1.7 4.0e-06 1702 [1.4,2.2]
0.5 1 1.5 2 2.5 3 3.5 4 4.5
HR

### Figure S6

**a.**

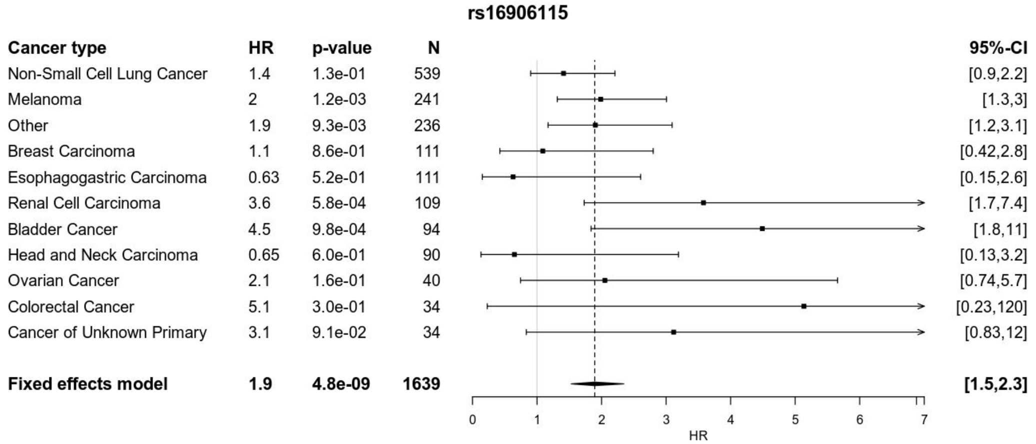
rs16906115
Cancer type
HR
p-value
N
95%-CI
Non-Small Cell Lung Cancer 1.4 1.3e-01 539 [0.9,2.2]
Melanoma 2 1.2e-03 241 [1.3,3]
Other 1.9 9.3e-03 236 [1.2,3.1]
Breast Carcinoma 1.1 8.6e-01 111 [0.42,2.8]
Esophagogastric Carcinoma 0.63 5.2e-01 111 [0.15,2.6]
Renal Cell Carcinoma 3.6 5.8e-04 109 [1.7,7.4]
Bladder Cancer 4.5 9.8e-04 94 [1.8,11]
Head and Neck Carcinoma 0.65 6.0e-01 90 [0.13,3.2]
Ovarian Cancer 2.1 1.6e-01 40 [0.74,5.7]
Colorectal Cancer 5.1 3.0e-01 34 [0.23,120]
Cancer of Unknown Primary 3.1 9.1e-02 34 [0.83,12]
Fixed effects model 1.9 4.8e-09 1639 [1.5,2.3]
0 1 2 3 4 5 6 7
HR

**b.**

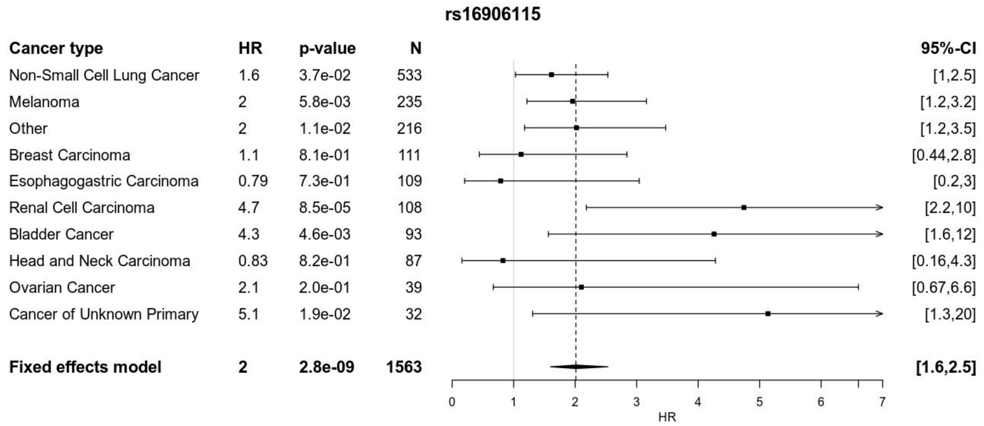
rs16906115
Cancer type
HR
p-value
N
95%-CI
Non-Small Cell Lung Cancer 1.6 3.7e-02 533 [1,2.5]
Melanoma 2 5.8e-03 235 [1.2,3.2]
Other 2 1.1e-02 216 [1.2,3.5]
Breast Carcinoma 1.1 8.1e-01 111 [0.44,2.8]
Esophagogastric Carcinoma 0.79 7.3e-01 109 [0.2,3]
Renal Cell Carcinoma 4.7 8.5e-05 108 [2.2,10]
Bladder Cancer 4.3 4.6e-03 93 [1.6,12]
Head and Neck Carcinoma 0.83 8.2e-01 87 [0.16,4.3]
Ovarian Cancer 2.1 2.0e-01 39 [0.67,6.6]
Cancer of Unknown Primary 5.1 1.9e-02 32 [1.3,20]
Fixed effects model 2 2.8e-09 1563 [1.6,2.5]
0 1 2 3 4 5 6 7
HR

### Figure S7

a.

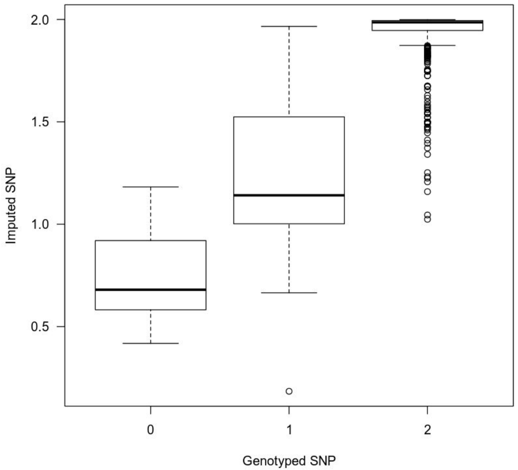


b.

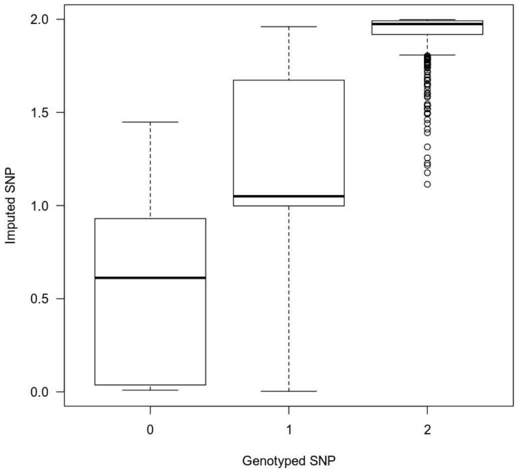


c.

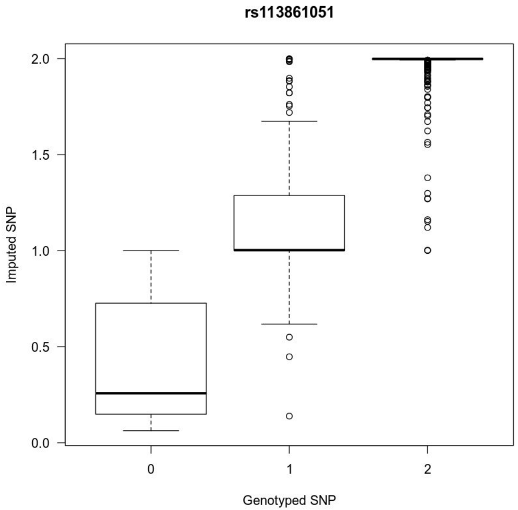

### Figure S8

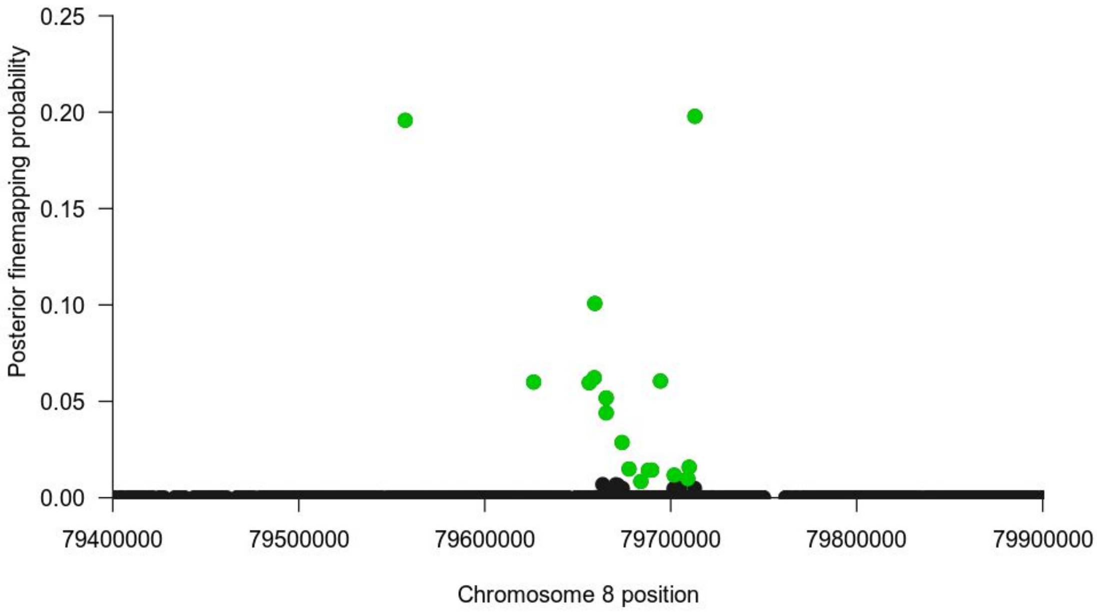

Posterior finemapping probability
0.25
0.20
0.15
0.10
0.05
0.00
79400000
79500000
79600000
79700000
79800000
79900000
Chromosome 8 position

### Figure S9

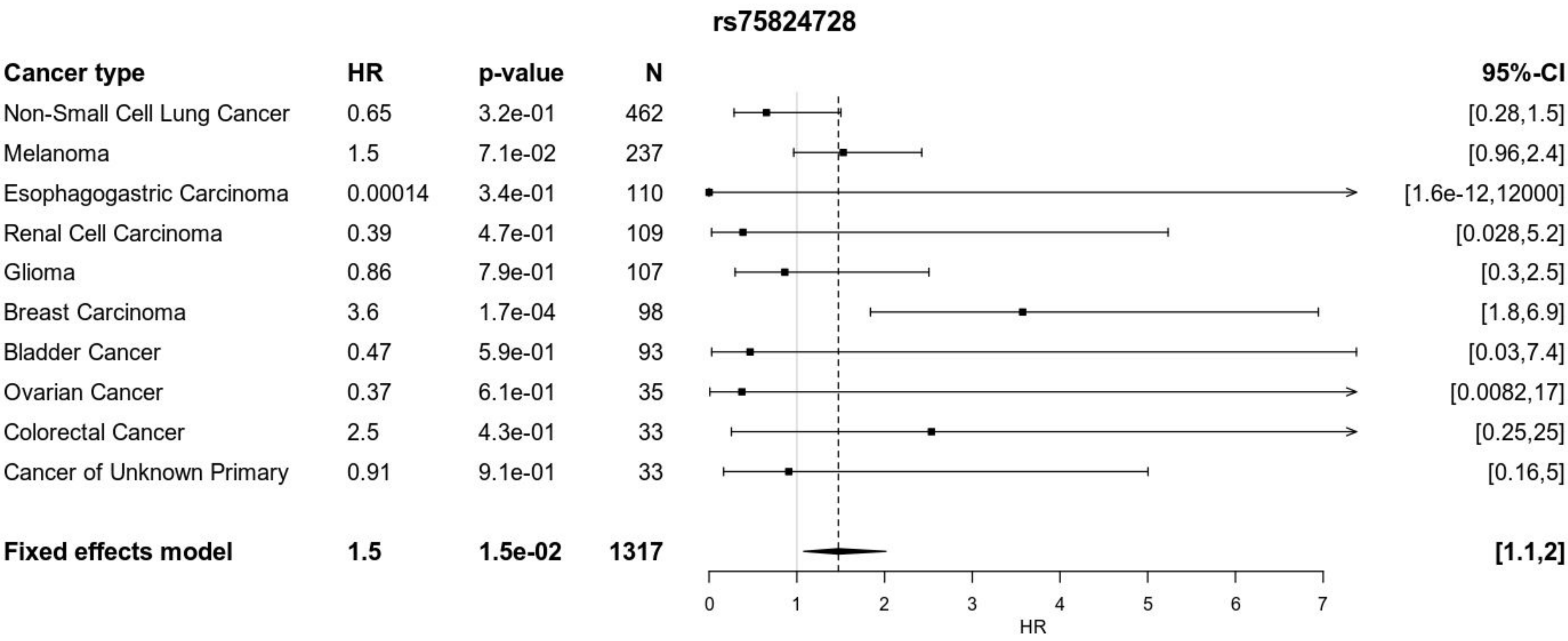
rs75824728
Cancer type
HR
p-value
N
95%-CI
Non-Small Cell Lung Cancer 0.65 3.2e-01 462 [0.28,1.5]
Melanoma 1.5 7.1e-02 237 [0.96,2.4]
Esophagogastric Carcinoma 0.00014 3.4e-01 110 [1.6e-12,12000]
Renal Cell Carcinoma 0.39 4.7e-01 109 [0.028,5.2]
Glioma 0.86 7.9e-01 107 [0.3,2.5]
Breast Carcinoma 3.6 1.7e-04 98 [1.8,6.9]
Bladder Cancer 0.47 5.9e-01 93 [0.03,7.4]
Ovarian Cancer 0.37 6.1e-01 35 [0.0082,17]
Colorectal Cancer 2.5 4.3e-01 33 [0.25,25]
Cancer of Unknown Primary 0.91 9.1e-01 33 [0.16,5]
Fixed effects model 1.5 1.5e-02 1317 [1.1,2]
0
1
2
3
4
5
6
7
HR

**Figure S10**

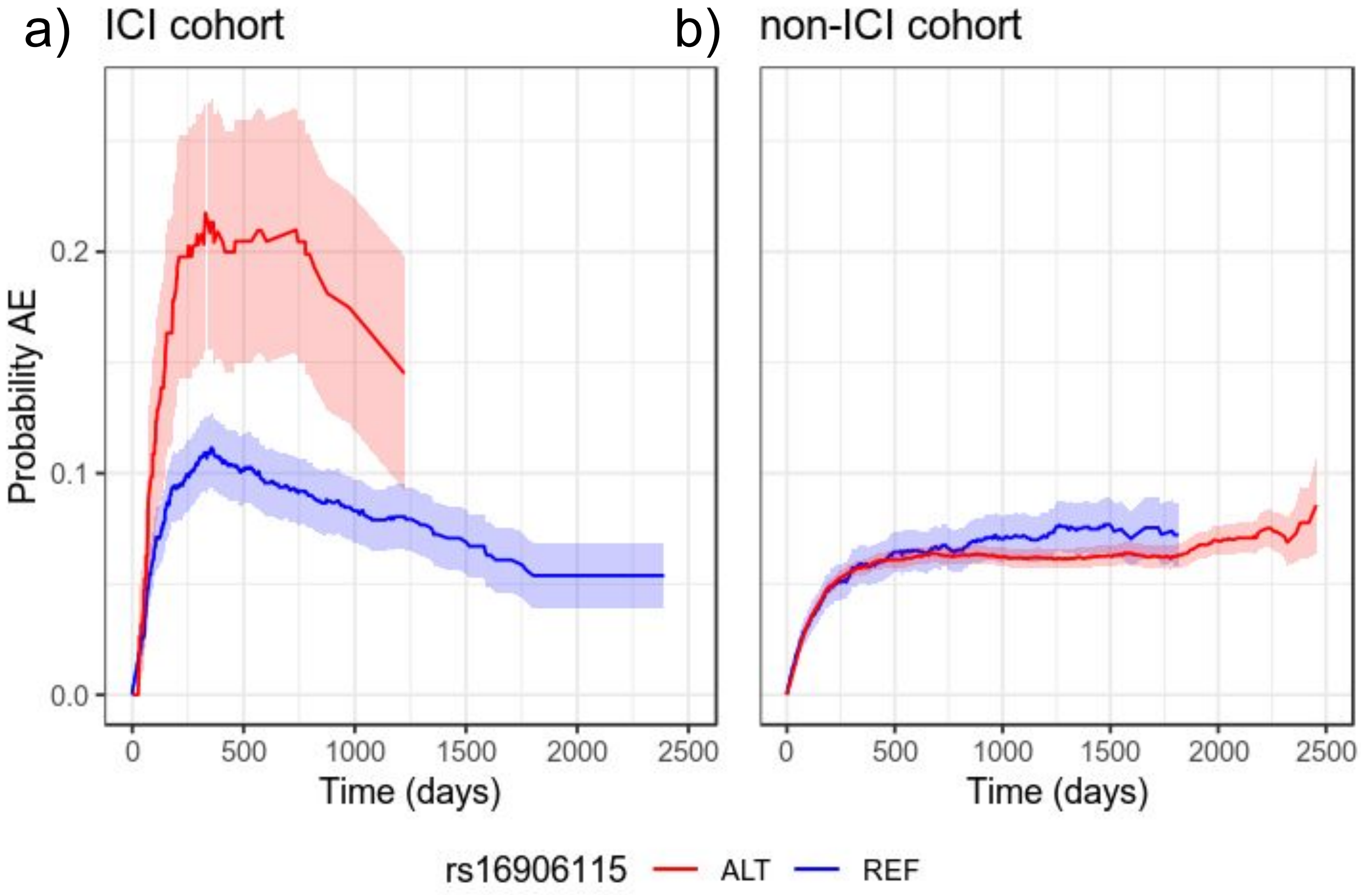

a) ICI cohort
b) non-ICI cohort
Probability AE
0.0
0.1
0.2
0
500
1000
1500
2000
2500
Time (days)
rs16906115
ALT
REF

### Figure S11

**a.**

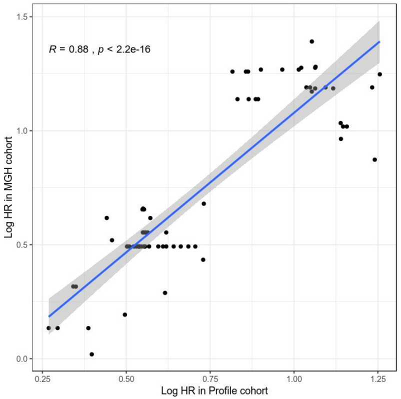


**b.**

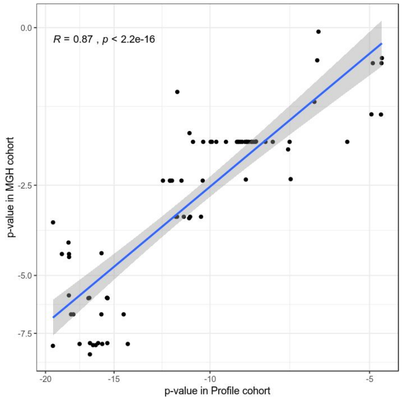


**c.**

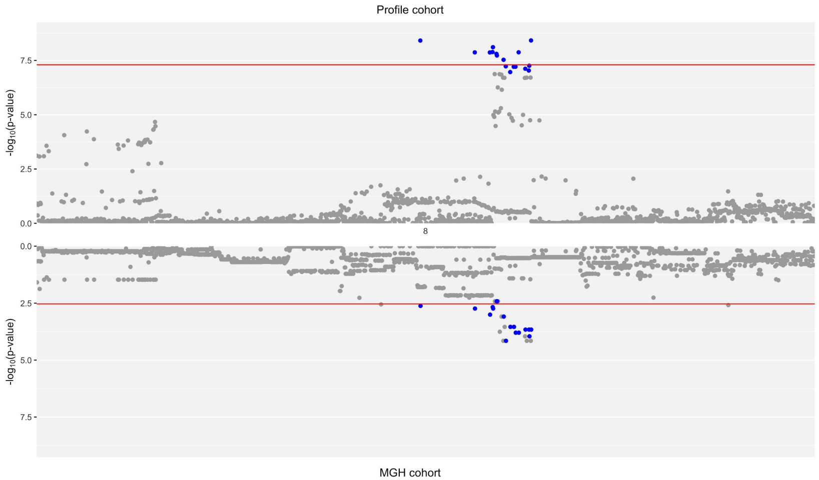

### Figure S12

**a.**

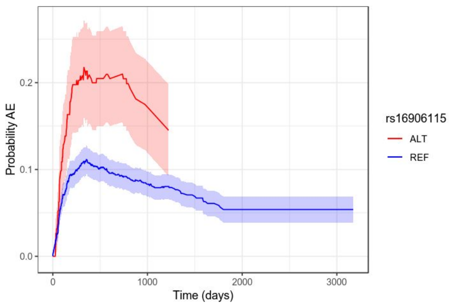


**b.**

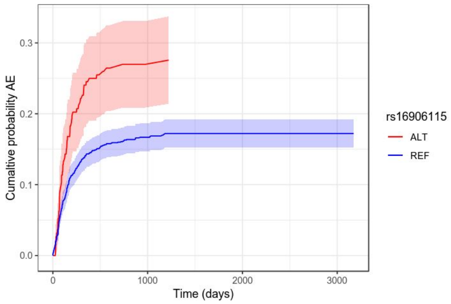


**c.**

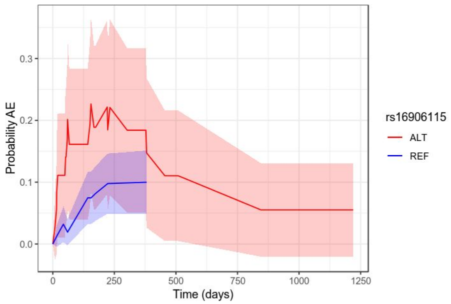


**d.**

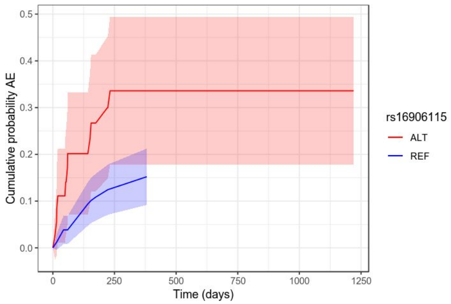

### Figure S13

(a)

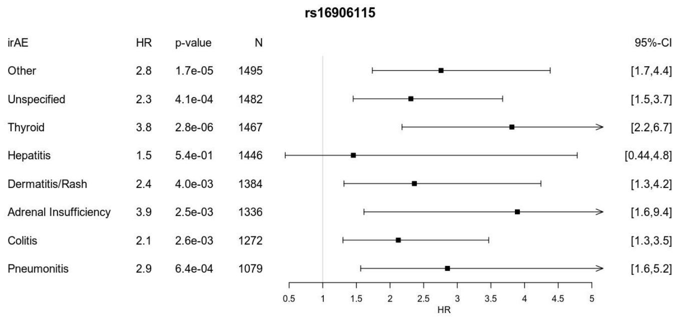


(b)

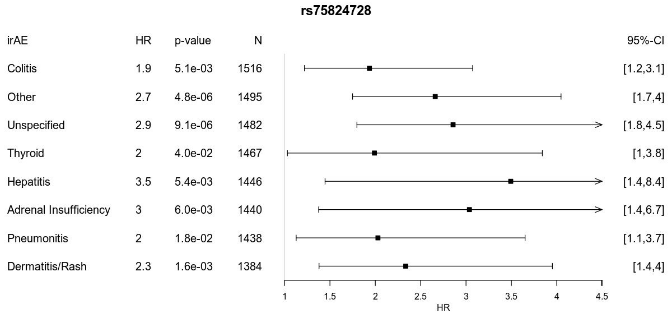


(c)

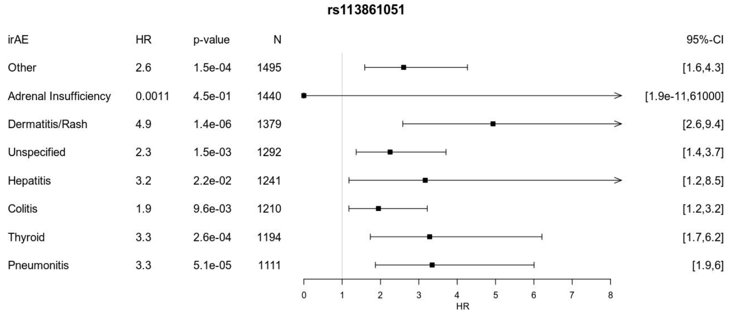

### Figure S14

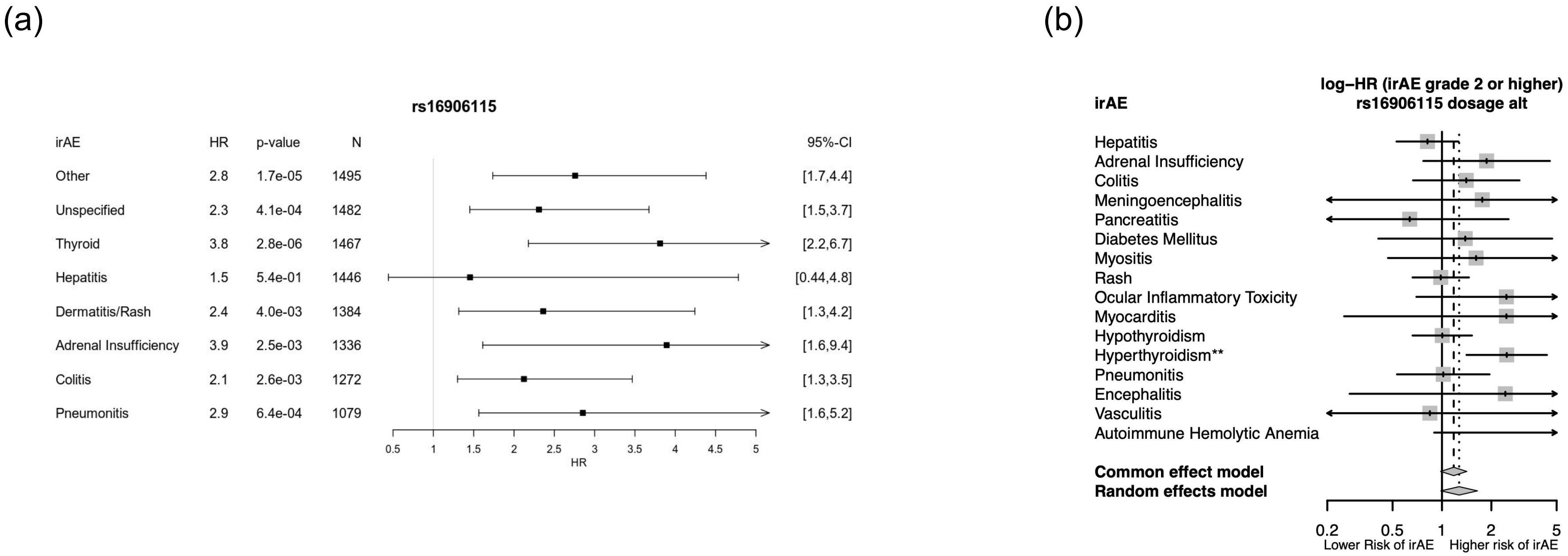

(a)
rs16906115
irAE HR p-value N 95%-CI
Other 2.8 1.7e-05 1495 [1.7,4.4]
Unspecified 2.3 4.1e-04 1482 [1.5,3.7]
Thyroid 3.8 2.8e-06 1467 [2.2,6.7]
Hepatitis 1.5 5.4e-01 1446 [0.44,4.8]
Dermatitis/Rash 2.4 4.0e-03 1384 [1.3,4.2]
Adrenal Insufficiency 3.9 2.5e-03 1336 [1.6,9.4]
Colitis 2.1 2.6e-03 1272 [1.3,3.5]
Pneumonitis 2.9 6.4e-04 1079 [1.6,5.2]
0.5 1 1.5 2 2.5 3 3.5 4 4.5 5
HR
(b)
log–HR (irAE grade 2 or higher)
rs16906115 dosage alt
irAE
Hepatitis
Adrenal Insufficiency
Colitis
Meningoencephalitis
Pancreatitis
Diabetes Mellitus
Myositis
Rash
Ocular Inflammatory Toxicity
Myocarditis
Hypothyroidism
Hyperthyroidism**
Pneumonitis
Encephalitis
Vasculitis
Autoimmune Hemolytic Anemia
Common effect model
Random effects model
0.2 0.5 1 2 5
Lower Risk of irAE
Higher risk of irAE

### Figure S15

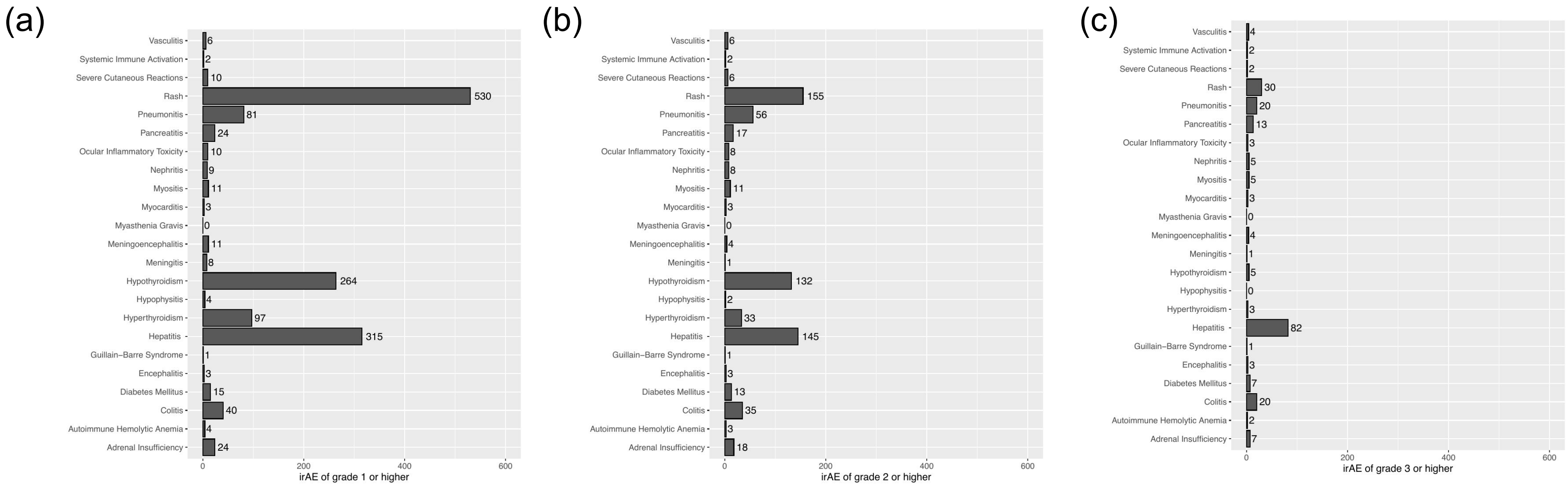
(a)
Vasculitis 6
Systemic Immune Activation 2
Severe Cutaneous Reactions 10
Rash 530
Pneumonitis 81
Pancreatitis 24
Ocular Inflammatory Toxicity 10
Nephritis 9
Myositis 11
Myocarditis 3
Myasthenia Gravis 0
Meningoencephalitis 11
Meningitis 8
Hypothyroidism 264
Hypophysitis 4
Hyperthyroidism 97
Hepatitis 315
Guillain–Barre Syndrome 1
Encephalitis 3
Diabetes Mellitus 15
Colitis 40
Autoimmune Hemolytic Anemia 4
Adrenal Insufficiency 24
0
200
400
600
irAE of grade 1 or higher
(b)
Vasculitis 6
Systemic Immune Activation 2
Severe Cutaneous Reactions 6
Rash 155
Pneumonitis 56
Pancreatitis 17
Ocular Inflammatory Toxicity 8
Nephritis 8
Myositis 11
Myocarditis 3
Myasthenia Gravis 0
Meningoencephalitis 4
Meningitis 1
Hypothyroidism 132
Hypophysitis 2
Hyperthyroidism 33
Hepatitis 145
Guillain–Barre Syndrome 1
Encephalitis 3
Diabetes Mellitus 13
Colitis 35
Autoimmune Hemolytic Anemia 3
Adrenal Insufficiency 18
0
200
400
600
irAE of grade 2 or higher
(c)
Vasculitis 4
Systemic Immune Activation 2
Severe Cutaneous Reactions 2
Rash 30
Pneumonitis 20
Pancreatitis 13
Ocular Inflammatory Toxicity 3
Nephritis 5
Myositis 5
Myocarditis 3
Myasthenia Gravis 0
Meningoencephalitis 4
Meningitis 1
Hypothyroidism 5
Hypophysitis 0
Hyperthyroidism 3
Hepatitis 82
Guillain–Barre Syndrome 1
Encephalitis 3
Diabetes Mellitus 7
Colitis 20
Autoimmune Hemolytic Anemia 2
Adrenal Insufficiency 7
0
200
400
600
irAE of grade 3 or higher

### Figure S16

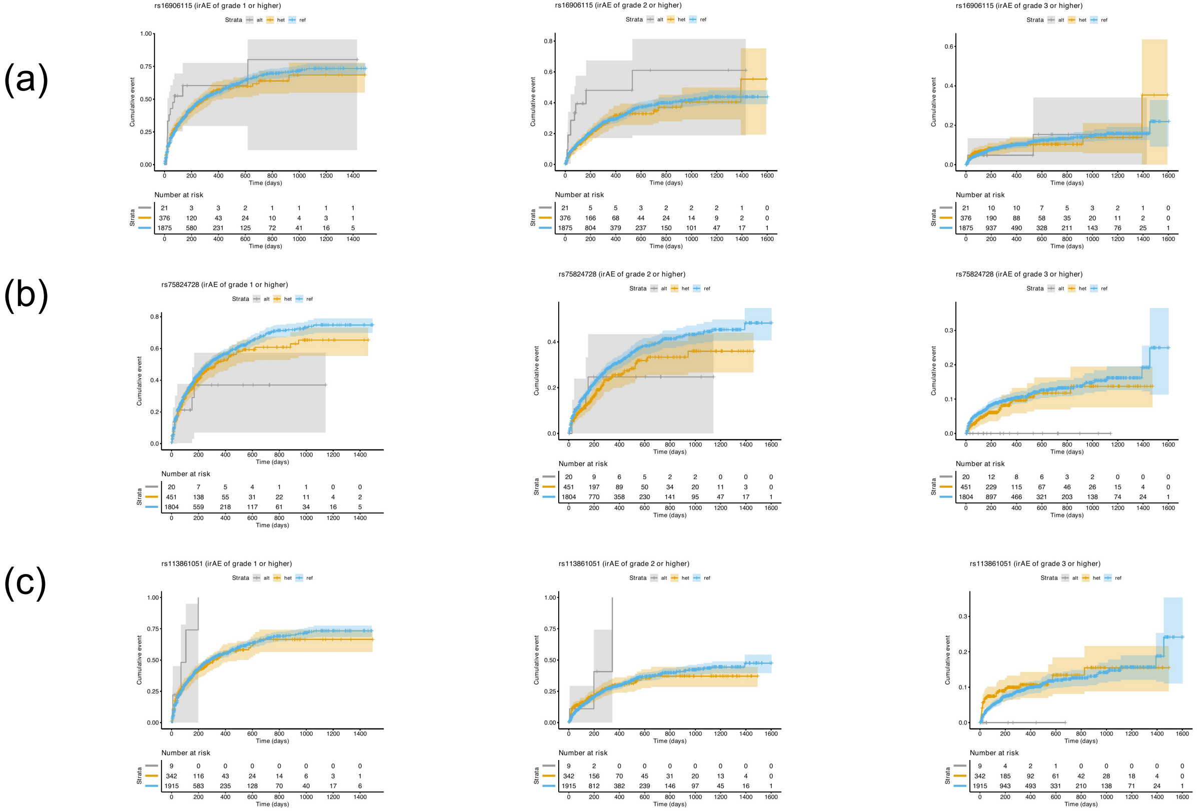

(a)
rs16906115 (irAE of grade 1 or higher)
rs16906115 (irAE of grade 2 or higher)
rs16906115 (irAE of grade 3 or higher)
Strata alt het ref
Cumulative event
Time (days)
Number at risk
Strata
21 3 3 2 1 1 1 1
376 120 43 24 10 4 3 1
1875 580 231 125 72 41 16 5
21 5 5 3 2 2 2 1 0
376 166 68 44 24 14 9 2 0
1875 804 379 237 150 101 47 17 1
21 10 10 7 5 3 2 1 0
376 190 88 58 35 20 11 2 0
1875 937 490 328 211 143 76 25 1
(b)
rs75824728 (irAE of grade 1 or higher)
rs75824728 (irAE of grade 2 or higher)
rs75824728 (irAE of grade 3 or higher)
20 7 5 4 1 1 0 0
451 138 55 31 22 11 4 2
1804 559 218 117 61 34 16 5
20 9 6 5 2 2 0 0 0
451 197 89 50 34 20 11 3 0
1804 770 358 230 141 95 47 17 1
20 12 8 6 3 2 0 0 0
451 229 115 67 46 26 15 4 0
1804 897 466 321 203 138 74 24 1
(c)
rs113861051 (irAE of grade 1 or higher)
rs113861051 (irAE of grade 2 or higher)
rs113861051 (irAE of grade 3 or higher)
9 0 0 0 0 0 0 0
342 116 43 24 14 6 3 1
1915 583 235 128 70 40 17 6
9 2 0 0 0 0 0 0 0
342 156 70 45 31 20 13 4 0
1915 812 382 239 146 97 45 16 1
9 4 2 1 0 0 0 0 0
342 185 92 61 42 28 18 4 0
1915 943 493 331 210 138 71 24 1

### Figure S17

(a)

(b)

(c)

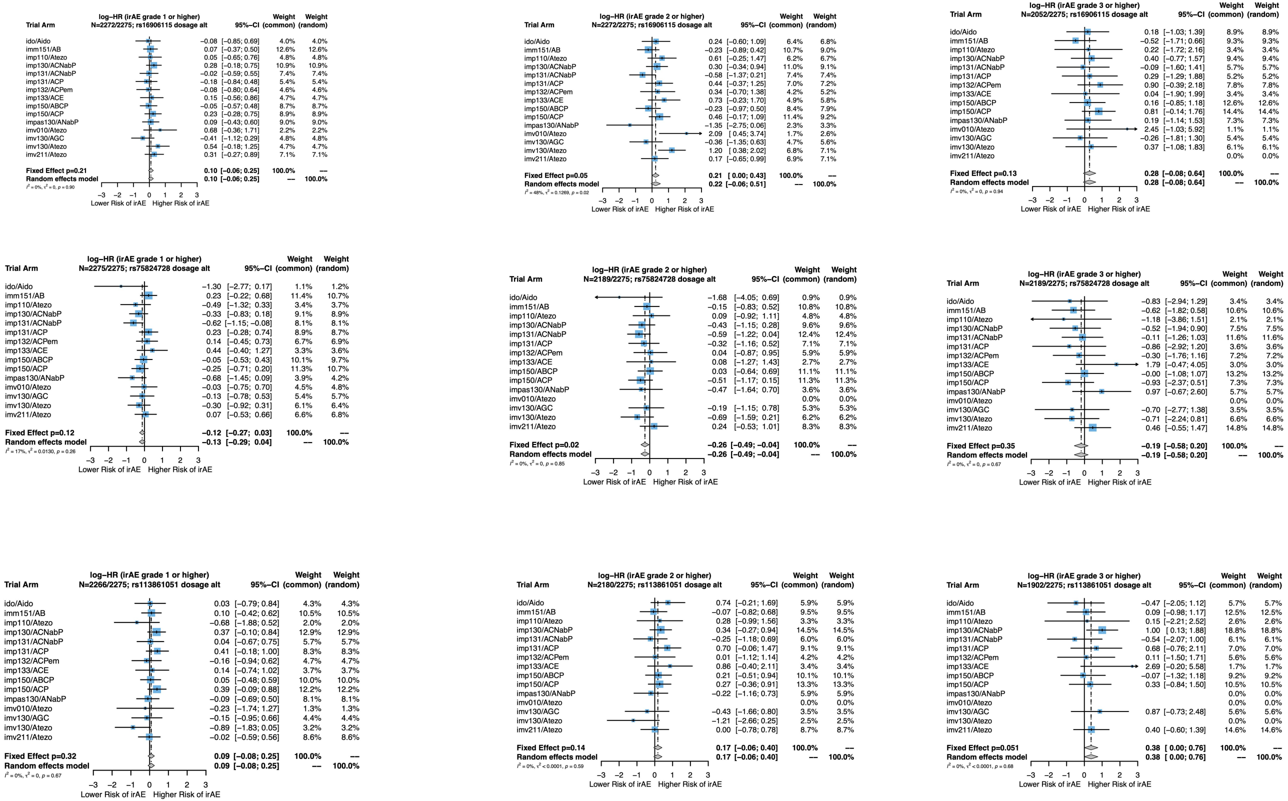

### Figure S18

(a)

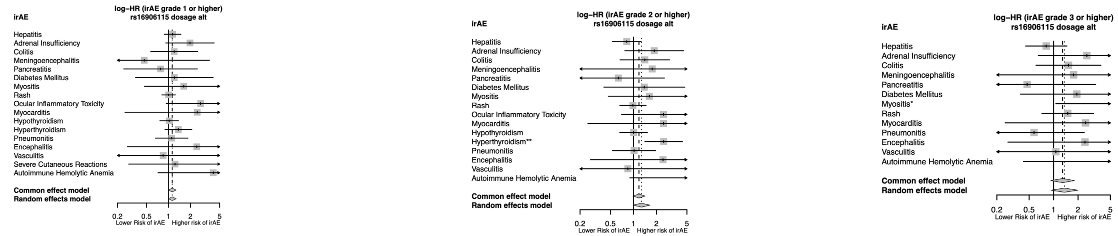

(b)

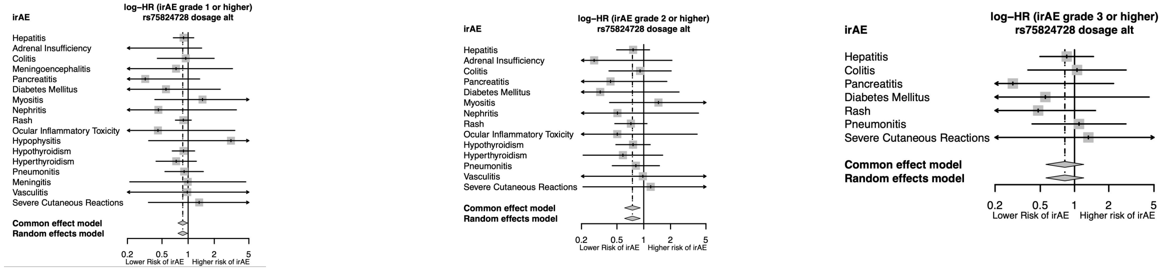

(c)

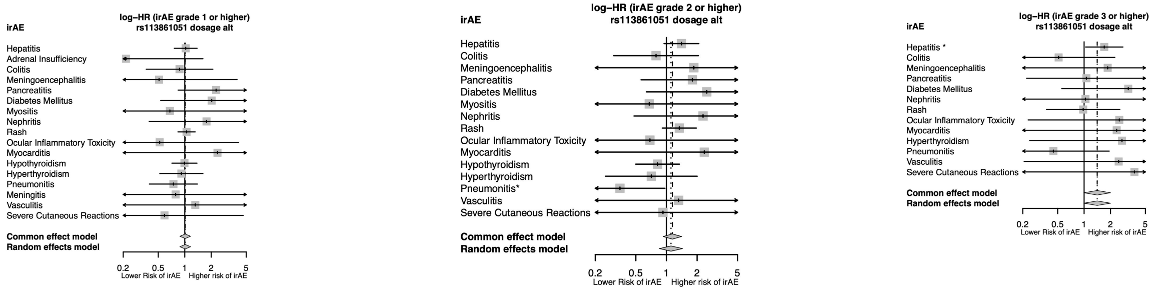

### Figure S19

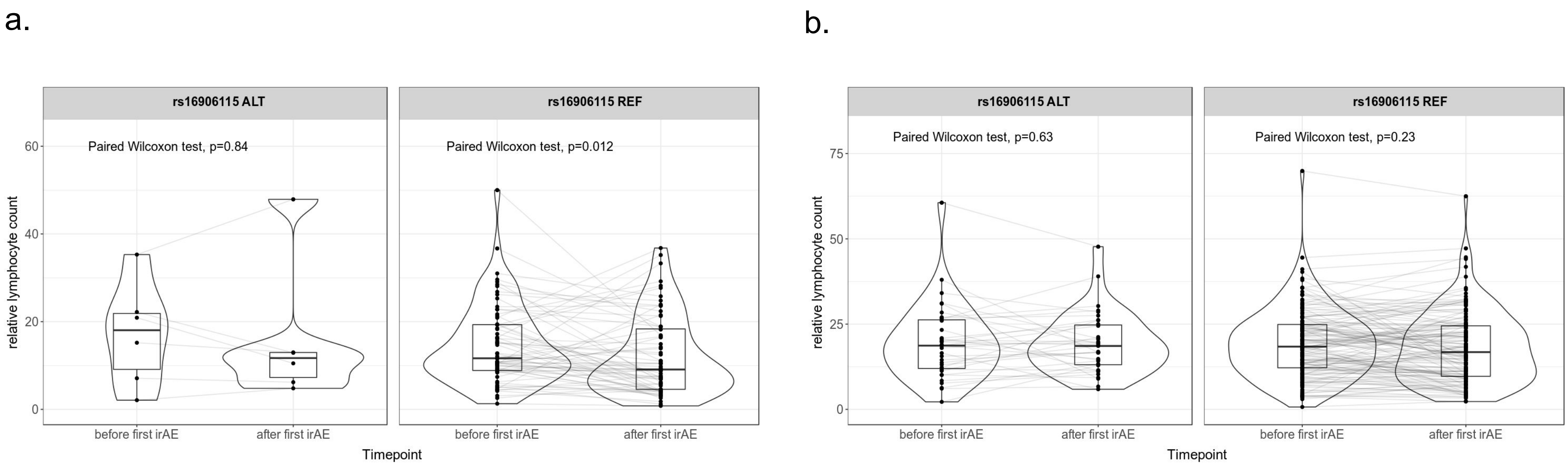
a.
rs16906115 ALT
rs16906115 REF
Paired Wilcoxon test, p=0.84
Paired Wilcoxon test, p=0.012
relative lymphocyte count
0
20
40
60
before first irAE
after first irAE
before first irAE
after first irAE
Timepoint
b.
rs16906115 ALT
rs16906115 REF
Paired Wilcoxon test, p=0.63
Paired Wilcoxon test, p=0.23
relative lymphocyte count
0
25
50
75
before first irAE
after first irAE
before first irAE
after first irAE
Timepoint

### Figure S20

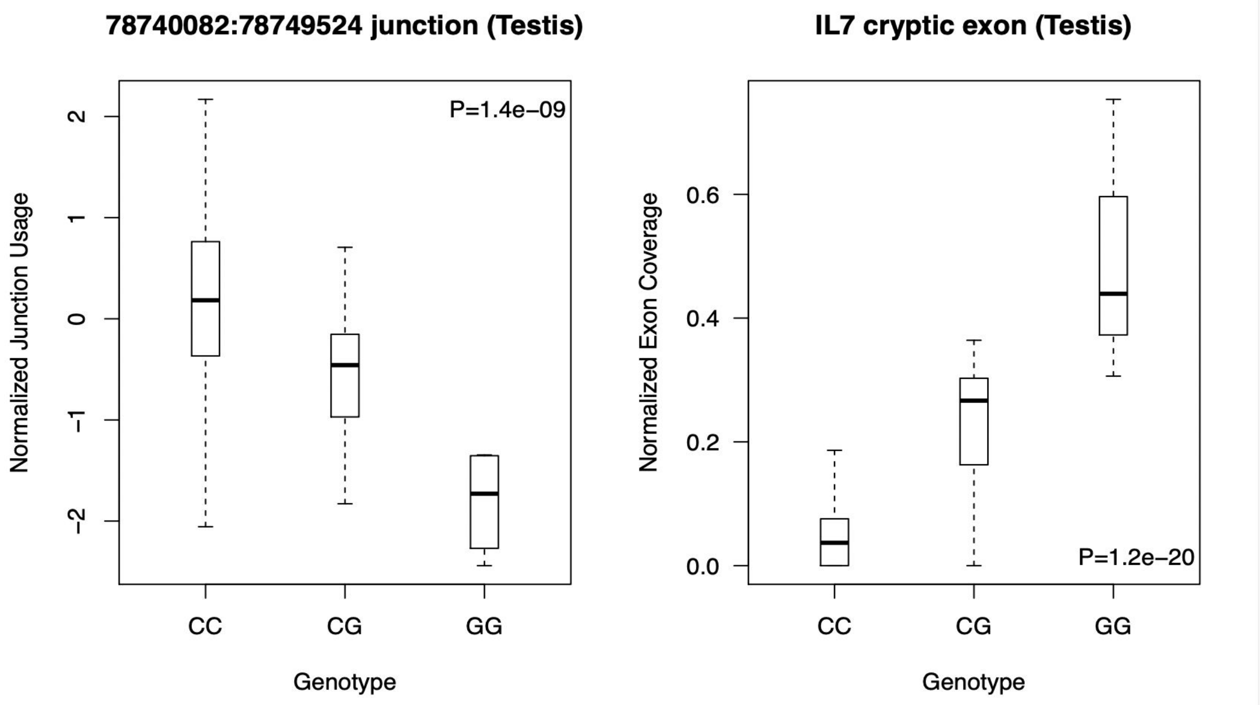
78740082:78749524 junction (Testis)
P=1.4e−09
Normalized Junction Usage
2
1
0
−1
−2
CC
CG
GG
Genotype
IL7 cryptic exon (Testis)
P=1.2e−20
Normalized Exon Coverage
0.6
0.4
0.2
0.0
CC
CG
GG
Genotype

### Figure S21

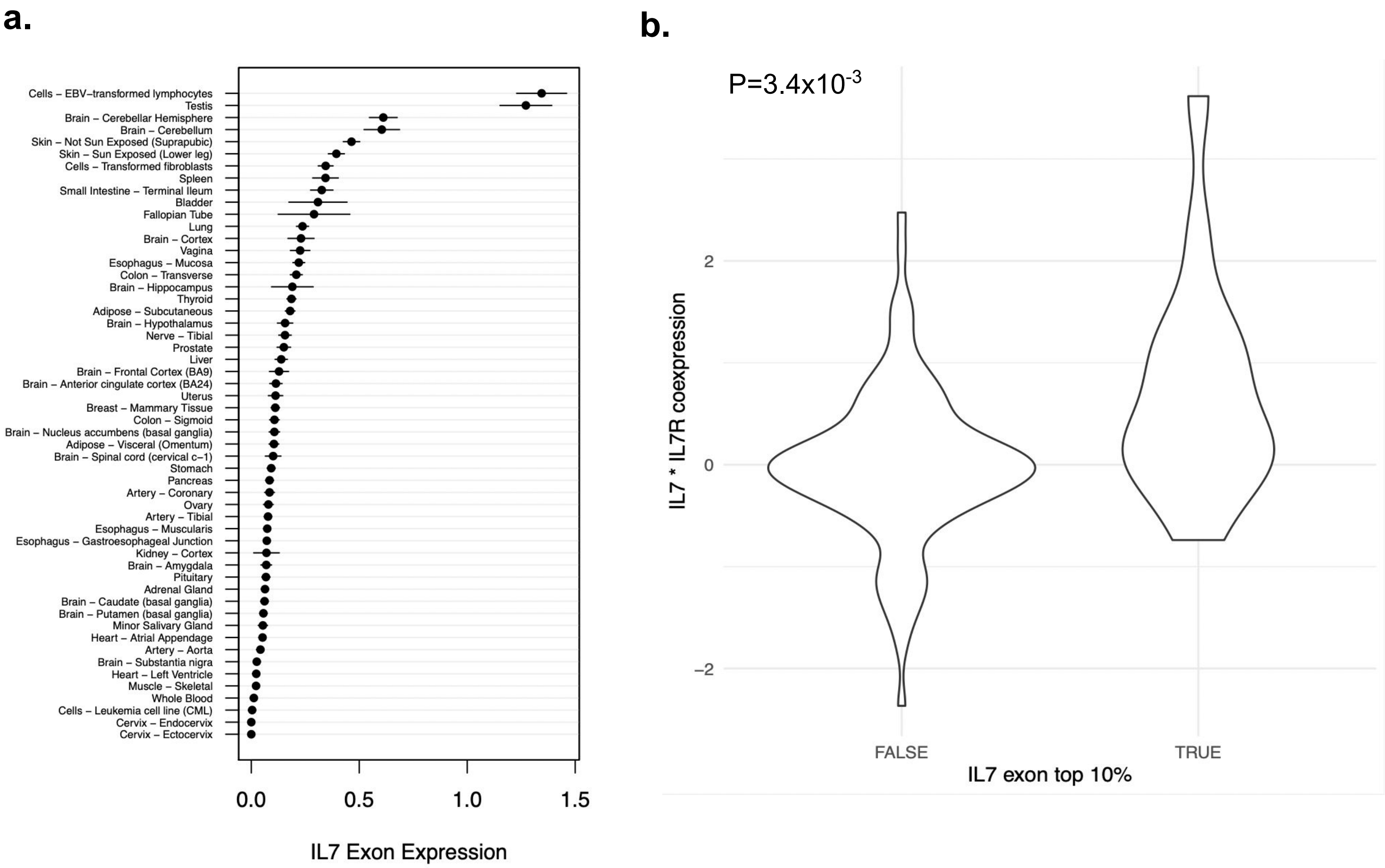
a.
Cells – EBV–transformed lymphocytes
Testis
Brain – Cerebellar Hemisphere
Brain – Cerebellum
Skin – Not Sun Exposed (Suprapubic)
Skin – Sun Exposed (Lower leg)
Cells – Transformed fibroblasts
Spleen
Small Intestine – Terminal Ileum
Bladder
Fallopian Tube
Lung
Brain – Cortex
Vagina
Esophagus – Mucosa
Colon – Transverse
Brain – Hippocampus
Thyroid
Adipose – Subcutaneous
Brain – Hypothalamus
Nerve – Tibial
Prostate
Liver
Brain – Frontal Cortex (BA9)
Brain – Anterior cingulate cortex (BA24)
Uterus
Breast – Mammary Tissue
Colon – Sigmoid
Brain – Nucleus accumbens (basal ganglia)
Adipose – Visceral (Omentum)
Brain – Spinal cord (cervical c–1)
Stomach
Pancreas
Artery – Coronary
Ovary
Artery – Tibial
Esophagus – Muscularis
Esophagus – Gastroesophageal Junction
Kidney – Cortex
Brain – Amygdala
Pituitary
Adrenal Gland
Brain – Caudate (basal ganglia)
Brain – Putamen (basal ganglia)
Minor Salivary Gland
Heart – Atrial Appendage
Artery – Aorta
Brain – Substantia nigra
Heart – Left Ventricle
Muscle – Skeletal
Whole Blood
Cells – Leukemia cell line (CML)
Cervix – Endocervix
Cervix – Ectocervix
0.0
0.5
1.0
1.5
IL7 Exon Expression
b.
P=3.4x10-3
IL7 * IL7R coexpression
2
0
−2
FALSE
TRUE
IL7 exon top 10%

### Figure S22

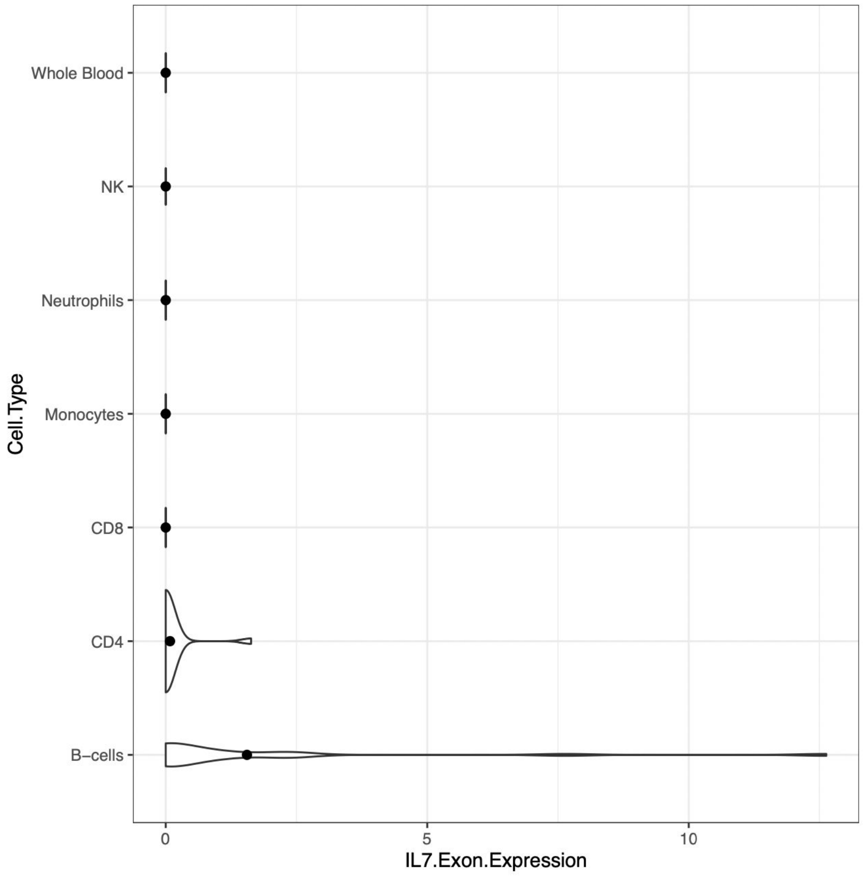

Whole Blood
NK
Neutrophils
Monocytes
CD8
CD4
B−cells
Cell.Type
0
5
10
IL7.Exon.Expression

### Figure S23

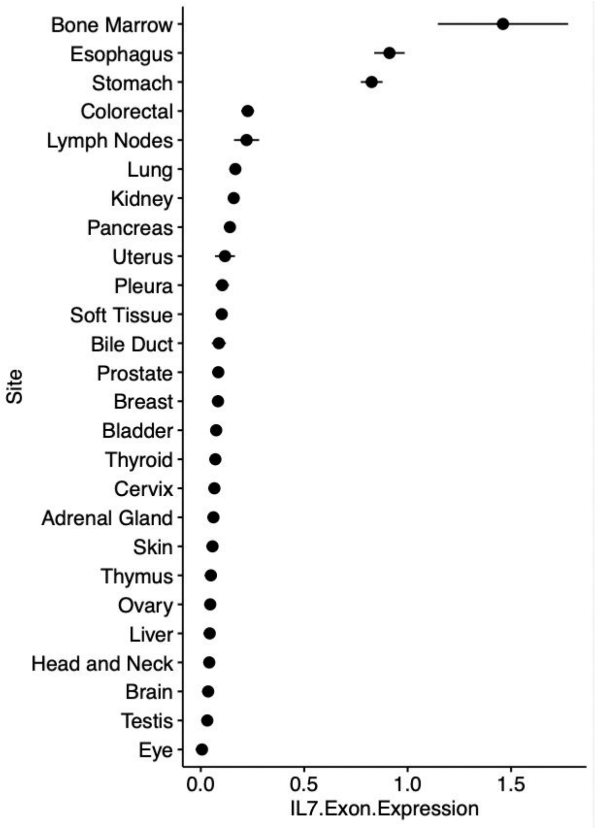

Bone Marrow
Esophagus
Stomach
Colorectal
Lymph Nodes
Lung
Kidney
Pancreas
Uterus
Pleura
Soft Tissue
Bile Duct
Prostate
Breast
Bladder
Thyroid
Cervix
Adrenal Gland
Skin
Thymus
Ovary
Liver
Head and Neck
Brain
Testis
Eye
Site
0.0
0.5
1.0
1.5
IL7.Exon.Expression

#### Previous autoimmune disease

| Cancer type | HR | p-value | N | 95%-CI |
|---|---|---|---|---|
| Non-Small Cell Lung Cancer | 1.2 | 5.0e-01 | 539 | [0.69,2.2] |
| Melanoma | 1.1 | 8.1e-01 | 241 | [0.56,2.1] |
| Other | 1.8 | 2.0e-01 | 236 | [0.74,4.3] |
| Breast Carcinoma | 0.52 | 5.3e-01 | 111 | [0.068,4] |
| Esophagogastric Carcinoma | 4.8 | 9.2e-03 | 111 | [1.5,16] |
| Renal Cell Carcinoma | 1.5 | 4.6e-01 | 109 | [0.54,3.9] |
| Bladder Cancer | 1.7 | 3.5e-01 | 94 | [0.55,5.4] |
| Head and Neck Carcinoma | 1.9 | 4.5e-01 | 90 | [0.36,10] |
| Ovarian Cancer | 11 | 2.8e-02 | 40 | [1.3,100] |
| Colorectal Cancer | 2.1e-07 | 1.0e+00 | 34 | [0,Inf] |
| Cancer of Unknown Primary | 6.1 | 1.0e-01 | 34 | [0.71,53] |
| **Fixed effects model** | **1.5** | **6.4e-03** | **1639** | **[1.1,2.1]** |

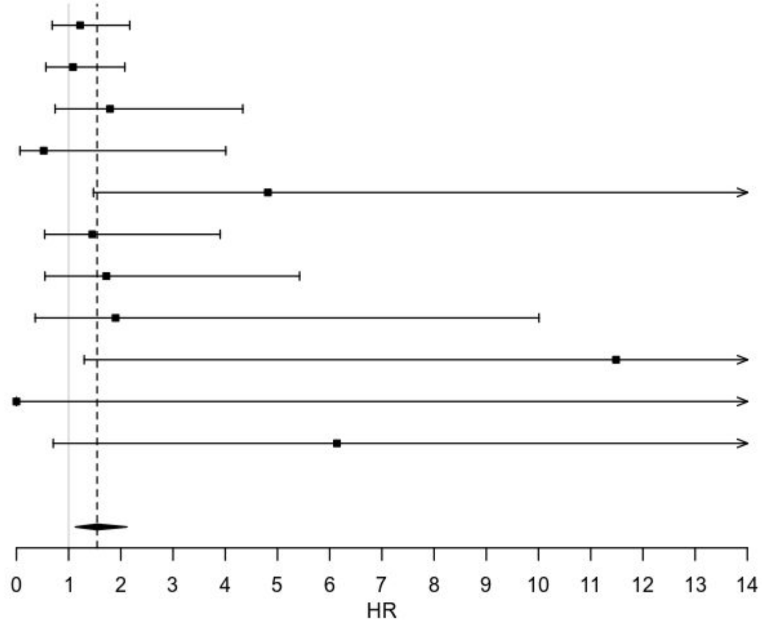


Figure S24: Previous autoimmune disease

**Male sex**

| Cancer type | HR | p-value | N | 95%-CI |
|---|---|---|---|---|
| Non-Small Cell Lung Cancer | 0.78 | 3.0e-01 | 539 | [0.49,1.2] |
| Melanoma | 1.2 | 3.7e-01 | 241 | [0.78,2] |
| Other | 1.4 | 4.1e-01 | 236 | [0.65,2.9] |
| Glioma | 1.1 | 8.9e-01 | 112 | [0.19,6.9] |
| Breast Carcinoma | | | 111 | |
| Esophagogastric Carcinoma | 0.99 | 9.9e-01 | 111 | [0.21,4.7] |
| Renal Cell Carcinoma | 0.59 | 2.8e-01 | 109 | [0.22,1.6] |
| Bladder Cancer | 0.89 | 8.3e-01 | 94 | [0.31,2.6] |
| Head and Neck Carcinoma | 0.7 | 6.7e-01 | 90 | [0.14,3.6] |
| Ovarian Cancer | | | 40 | |
| Colorectal Cancer | 140 | 1.7e-02 | 34 | [2.4,7900] |
| Cancer of Unknown Primary | 2.8 | 2.1e-01 | 34 | [0.55,15] |
| **Fixed effects model** | **1** | **8.6e-01** | **1751** | **[0.79,1.3]** |

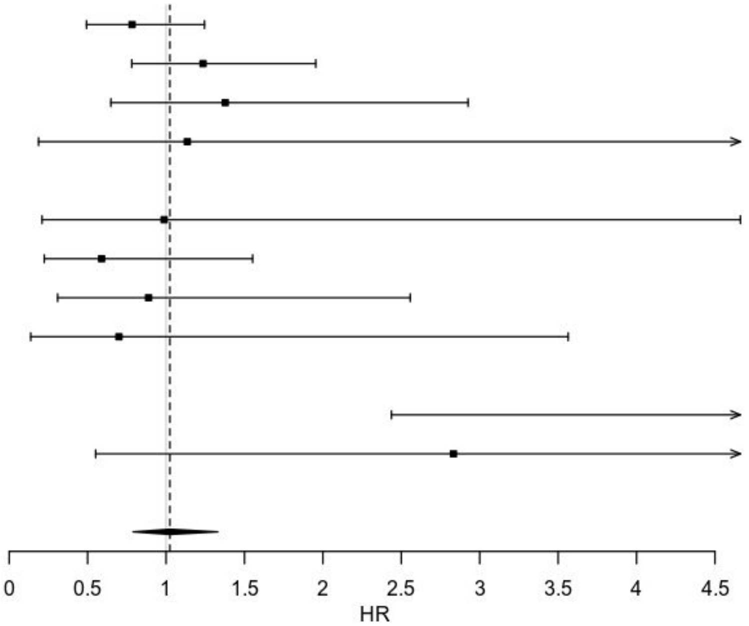


Figure S25: Sex difference

### Table S1

| Immune Feature | IL7 Exon Usage Z-Score | IL7 Exon Usage P-value | IL7 Expression Z-Score | IL7 Expression P-value | IL7 Exon \| IL7 expression Z-Score | IL7 Exon \| IL7 expression P-value |
|---|---|---|---|---|---|---|
| Leukocyte.Fraction | 9.2 | 7E-20 | 8.2 | 3E-16 | **7.7** | 2E-14 |
| Stromal.Fraction | 8.1 | 6E-16 | 3.4 | 7E-04 | **7.6** | 4E-14 |
| Proliferation | 7.2 | 5E-13 | -1.4 | 2E-01 | **7.7** | 2E-14 |
| Wound.Healing | 7.1 | 1E-12 | 16.4 | 2E-59 | **4.0** | 6E-05 |
| Macrophage.Regulation | 7.1 | 1E-12 | -5.2 | 2E-07 | **8.3** | 9E-17 |
| Lymphocyte.Infiltration.Signature.Score | 6.6 | 4E-11 | 4.1 | 4E-05 | **5.9** | 4E-09 |
| IFN.gamma.Response | 5.6 | 3E-08 | 7.2 | 9E-13 | **4.2** | 2E-05 |
| TGF.beta.Response | 5.5 | 4E-08 | 1.5 | 1E-01 | **5.3** | 1E-07 |
| BCR.Shannon | 5.3 | 1E-07 | 2.0 | 5E-02 | **5.0** | 8E-07 |
| TCR.Shannon | 4.0 | 7E-05 | 1.5 | 1E-01 | **3.8** | 2E-04 |
| TCR.Richness | 3.9 | 1E-04 | 7.7 | 2E-14 | 2.4 | 2E-02 |
| CTA.Score | 3.5 | 4E-04 | 1.8 | 8E-02 | 3.2 | 1E-03 |
| Th1.Cells | 3.3 | 1E-03 | 12.8 | 4E-37 | 0.8 | 4E-01 |
| Th2.Cells | 3.3 | 1E-03 | 21.8 | 5E-103 | -1.0 | 3E-01 |
| Th17.Cells | 2.8 | 6E-03 | -5.3 | 2E-07 | **3.9** | 8E-05 |
| B.Cells.Naive | 2.3 | 2E-02 | 4.0 | 6E-05 | 1.5 | 1E-01 |
| Dendritic.Cells.Resting | 1.0 | 3E-01 | -3.9 | 1E-04 | 1.8 | 7E-02 |
| Macrophages.M0 | 0.5 | 6E-01 | -9.9 | 4E-23 | 2.6 | 9E-03 |
| Macrophages.M1 | 0.4 | 7E-01 | 3.8 | 2E-04 | -0.4 | 7E-01 |
| Macrophages.M2 | 0.2 | 8E-01 | -4.8 | 2E-06 | 1.2 | 2E-01 |
| Mast.Cells.Activated | -0.1 | 1E+00 | 6.6 | 4E-11 | -1.4 | 2E-01 |
| NK.Cells.Activated | -0.6 | 5E-01 | -4.2 | 3E-05 | 0.2 | 8E-01 |
| Plasma.Cells | -0.7 | 5E-01 | -11.5 | 3E-30 | 1.7 | 9E-02 |
| T.Cells.CD4.Memory.Resting | -1.1 | 3E-01 | -3.7 | 2E-04 | -0.3 | 7E-01 |
| T.Cells.CD8 | -1.3 | 2E-01 | -6.7 | 2E-11 | 0.1 | 1E+00 |
| T.Cells.Follicular.Helper | -2.6 | 9E-03 | -4.7 | 3E-06 | -1.7 | 9E-02 |
| T.Cells.Regulatory.Tregs | -3.6 | 3E-04 | -1.2 | 2E-01 | **-3.4** | 7E-04 |
| Lymphocytes | -3.6 | 3E-04 | -6.6 | 4E-11 | -2.3 | 2E-02 |
| Macrophages | -4.2 | 2E-05 | -2.8 | 5E-03 | **-3.8** | 2E-04 |

**Positive association**

**Negative association**

Table S2: Interaction Analysis

| SNP | Chromosome | Position | Posterior probability | P-value in Profile cohort |
|---|---|---|---|---|
| rs16906115 | 8 | 79712998 | 0.1978415408 | 3.79E-09 |
| rs7011565 | 8 | 79557237 | 0.1957790135 | 3.83E-09 |
| rs16906062 | 8 | 79659182 | 0.1007706209 | 7.75E-09 |
| rs7816685 | 8 | 79658906 | 0.06223026669 | 1.29E-08 |
| rs73251644 | 8 | 79694464 | 0.06051930009 | 1.33E-08 |
| rs73249933 | 8 | 79626275 | 0.05998693589 | 1.34E-08 |
| rs74197776 | 8 | 79656182 | 0.05961872189 | 1.35E-08 |
| rs117355070 | 8 | 79665307 | 0.05176578818 | 1.57E-08 |
| rs116228140 | 8 | 79665329 | 0.04399633208 | 1.87E-08 |
| rs141792902 | 8 | 79673847 | 0.02865189034 | 2.95E-08 |
| rs4739139 | 8 | 79709926 | 0.01583309901 | 5.54E-08 |
| rs59286098 | 8 | 79677592 | 0.01490514512 | 5.91E-08 |
| rs16906090 | 8 | 79689755 | 0.01434440485 | 6.16E-08 |
| rs10110519 | 8 | 79688026 | 0.01428458798 | 6.18E-08 |
| rs2717536 | 8 | 79701876 | 0.01170948825 | 7.64E-08 |
| rs9942766 | 8 | 79709182 | 0.009834019143 | 9.20E-08 |
| rs2919935 | 8 | 79683948 | 0.008440074784 | 1.08E-07 |

Table S3: 95% credible set.

| Study name | Reference |
| --- | --- |
| IDO inhibitor trial (ido) | Jung KH, LoRusso P, Burris H, et al., Clin Cancer Res, 2019 |
| imm151 / IMmotion151 | Rini BI, Powles T, Atkins MB, et al, Lancet, 2019 |
| imp110 / IMpower110 | Herbst RS, Giaccone G, de Marinis F, et al., N Engl J Med, 2020 |
| imp130 / IMpower130 | West H, McCleod M, Hussein M, et al., Lancet Oncol., 2019 |
| imp131 / IMpower131 | Jotte R, Cappuzzo F, Vynnychenko I, et al., J Thorac Oncol., 2020 |
| imp132 / IMpower132 | Nishio M, Barlesi F, West H, et al., J Thorac Oncol., 2021 |
| imp133 / IMpower133 | Horn L, Mansfield AS, Szczęsna A, et al., N Engl J Med., 2018 |
| imp150 / IMpower150 | Socinski MA, Jotte RM, Cappuzzo F, et al., N Engl J Med., 2018 |
| impas130 /IMpassion130 | Schmid P, Adams S, Rugo HS, et al., N Engl J Med., 2018 |
| | Schmid P, Rugo HS, Adams S, et al., Lancet Oncol., 2020 |
| imv010 / IMvigor010 | Bellmunt J, Hussain M, Gschwend JE, et al., Lancet Oncol., 2021 |
| imv130 / IMvigor130 | Galsky MD, Arija JÁA, Bamias A, et al., Lancet, 2020 |
| imv211 / IMvigor211 | Powles T, Durán I, van der Heijden MS, et al., Lancet, 2018 |

Table S4: Clinical trial replication cohorts

| | | Profile cohort | | | MGH cohort | | | Clinical trial cohort | |
|---|---|---|---|---|---|---|---|---|---|
| SNP | Ref>Alt | HR [95% conf int] | p-value | Frequency | HR [95% conf int] | p-value | Frequency | HR [95% conf int] | p-value |
| rs16906115 | G>A | 2.0 [1.6-2.5] | 3.8x10-9 | 0.08 | 3.6 [1.8-7.1] | 0.00028 | 0.10 | 1.2 [1.0-1.5] | 0.05 |
| rs75824728 | G>A | 1.9 [1.5-2.4] | 8.4x10-9 | 0.10 | 1.49 [0.67-3.2] | 0.4 | 0.08 | 0.77 [0.61-0.96] | 0.02 |
| rs113861051 | A>(G/T) | 2.0 [1.6-2.6] | 1.1x10-8 | 0.07 | 1.4 [0.50-3.7] | 0.55 | 0.08 | 1.18 [0.94-1.5] | 0.14 |

Table S5: Genome wide significant loci

| Cohort | Hazard ratio [95% conf int] | p-value |
|---|---|---|
| DFCI | 1.2 [0.98,1.5] | 0.083 |
| MGH | 1.3 [1.1-1.5] | 0.007 |

Table S6: Overall survival association

Table S7: Search terms

| | rs16906115 | | rs75824728 | | rs113861051 | |
|---|---|---|---|---|---|---|
| irAE type | ALT | REF | ALT | REF | ALT | REF |
| Colitis | 0.075 | 0.057 | 0.07 | 0.058 | 0.095 | 0.056 |
| Other | 0.086 | 0.042 | 0.11 | 0.039 | 0.11 | 0.04 |
| Thyroid | 0.08 | 0.025 | 0.042 | 0.03 | 0.061 | 0.029 |
| Hepatitis | 0.024 | 0.014 | 0.036 | 0.013 | 0.024 | 0.015 |
| Adrenal insufficiency | 0.024 | 0.016 | 0.036 | 0.015 | 0.008 | 0.018 |
| Pneumonitis | 0.042 | 0.031 | 0.048 | 0.03 | 0.11 | 0.024 |
| Dermatitis/Rash | 0.07 | 0.04 | 0.075 | 0.039 | 0.081 | 0.04 |

Table S8: irAE frequency in carriers and non-carriers by irAE type

| | | relative occurrence | | absolute occurrence | |
|---|---|---|---|---|---|
| | | Stopped therapy | Steroid given | Stopped therapy | Steroid given |
| **rs16906115** | ALT | 0.45 | 0.48 | 20 | 21 |
| | REF | 0.5 | 0.5 | 1 | 1 |
| **rs75824728** | ALT | 0.36 | 0.36 | 4 | 4 |
| | REF | 0.49 | 0.51 | 17 | 18 |
| **rs113861051** | ALT | 0 | 0 | 0 | 0 |
| | REF | 0.54 | 0.56 | 21 | 22 |

Table S9: Occurrence of terminated therapy or steroid administration after irAE

| grade of irAE | Number of events | Frequency |
|---|---|---|
| 1 | 8 | 0.18 |
| 2 | 19 | 0.42 |
| 3 | 15 | 0.33 |
| 4 | 2 | 0.044 |
| 5 | 1 | 0.022 |

Table S10

| test_name | HR | pval |
|---|---|---|
| ETV4 deletion | 5.731719668 | 0.003699400454 |
| PRS: PRSWEB_PHE | 0.7550949468 | 0.01001543655 |
| PRS: BRCA.ERPOS | 0.7198298798 | 0.01044343632 |
| SNV in PBRM1 | 3.785056795 | 0.01161673183 |
| PRS: UKB_460K.bp_ | 1.341783947 | 0.01183205383 |
| FKBP9 deletion | 23.06985775 | 0.01268981956 |
| PRS: PRSWEB_PHE | 0.7614820971 | 0.01491898934 |
| RAD21 amplificatior | 0.5100807525 | 0.01581436951 |
| MYC amplification | 0.5282454208 | 0.01641313591 |
| EXT1 amplification | 0.5224209595 | 0.01805440023 |
| HLA_B_3502 | 0.1824060558 | 0.01885730417 |
| TSC1 deletion | 2.267507641 | 0.01910335487 |
| RIT1 amplification | 0.3546174109 | 0.02092329033 |
| SNV in KEAP1 | 18.62636694 | 0.02224104713 |
| PRS: UKB_460K.bp_ | 1.297755261 | 0.02338238714 |
| MYBL1 amplificatio | 0.5130646042 | 0.02466486936 |
| PRS: MEDS_PMID3 | 1.271415031 | 0.02550118264 |
| SNV in BAP1 | 5.212250631 | 0.02665151397 |
| SNV in KDM5C | 8.984942194 | 0.02892369297 |
| NBN amplification | 0.5245835871 | 0.0290023963 |
| MCL1 amplification | 0.5476359034 | 0.02942844719 |
| EED deletion | 0.2025410208 | 0.02984498504 |
| GLI3 deletion | 20.15783465 | 0.03060921184 |
| PRS: PRSWEB_PHE | 0.8122150141 | 0.03282254513 |
| JAK1 deletion | 2.763798906 | 0.03376495058 |
| PRS: UKB_460K.dis | 1.301543093 | 0.03412615435 |
| GBA amplification | 0.3876058761 | 0.03639044686 |
| PTCH1 deletion | 2.058935475 | 0.03642655925 |
| RSPO2 amplificatior | 0.4444248628 | 0.03847852215 |
| PRKDC amplificatio | 0.5333109362 | 0.04039814261 |
| Lab value: MPV | 1.304653328 | 0.04075217595 |
| SNV in DNMT3A | 264.1946866 | 0.04083390644 |
| PRDM1 deletion | 0.4961774501 | 0.04085103024 |
| FANCC deletion | 1.998357002 | 0.04190215746 |
| HLA_A_0101 | 0.6695153848 | 0.04210819356 |
| NPRL2 amplificatior | 8.530256906 | 0.04329046232 |
| CDC73 deletion | 11.65016524 | 0.04347320142 |
| BCL2 amplification | 0.2117517967 | 0.04408131728 |
| FANCG amplificatio | 0.1693113757 | 0.04820008356 |
| BRCC3 amplificatio | 7.221922411 | 0.04832622552 |
| HLA_B_4403 | 1.801459696 | 0.04855363658 |
| PRS: Stratified_rect | 0.7949677598 | 0.04992908715 |

| | | |
|---|---|---|
| **HLA_B_3503** | 2.854659263 | 0.05046039981 |
| **Lab value: GLOB** | 0.6540182794 | 0.05388394331 |
| **NTRK2 deletion** | 1.939829268 | 0.05388878251 |
| **NOTCH1 deletion** | 1.922227665 | 0.05456673281 |
| **PRS: MEDS_PMID3** | 1.226524685 | 0.05507243575 |
| **PRS: UKB_460K.blo** | 1.253378239 | 0.05557997345 |
| **RECQL4 amplificati** | 0.5551500377 | 0.05757071479 |
| **PRKDC deletion** | 2.82974404 | 0.05790906468 |
| **PRS: PMID2786325** | 0.8226974173 | 0.06043442068 |
| **KRAS deletion** | 2.959313964 | 0.06174435205 |
| **POLE amplification** | 3.146149137 | 0.06404531867 |
| **XPA deletion** | 1.895261676 | 0.0643297655 |
| **MPL amplification** | 0.2806639645 | 0.06455896714 |
| **FLT4 amplification** | 2.165435643 | 0.06551916053 |
| **PRS: PMID2786325** | 0.8276926911 | 0.06593843775 |
| **PRS: PMID2786325** | 1.203457168 | 0.06594616918 |
| **SNV in RBM10** | 3.912205743 | 0.06603365102 |
| **Lab value: CA** | 1.784857218 | 0.0660394233 |
| **RHOA deletion** | 2.631459671 | 0.0666891275 |
| **Lab value: MG** | 0.1760334704 | 0.06669127974 |
| **COL7A1 deletion** | 2.644680642 | 0.06756404418 |
| **KCNQ1 deletion** | 3.100325708 | 0.06863997054 |
| **RBBP8 deletion** | 2.949557127 | 0.06866020384 |
| **PRKCZ deletion** | 0.3210232092 | 0.06878303899 |
| **AURKA deletion** | 7.291338306 | 0.06880891996 |
| **BRAF amplification** | 0.5271258845 | 0.06907776553 |
| **GNAS deletion** | 5.565842771 | 0.06995396938 |
| **Lab value: LDH** | 1.001399607 | 0.07021888922 |
| **ZNF217 deletion** | 7.174919959 | 0.07176635775 |
| **PRS: GLIOMA** | 1.19285215 | 0.07249755917 |
| **PRKCI deletion** | 0.1842391603 | 0.07411007153 |
| **PRS: TRICL.sma_ns** | 1.269822053 | 0.07416063287 |
| **Lab value: PT** | 1.178014941 | 0.07510240999 |
| **PRS: PMID2786325** | 0.8339632996 | 0.07599485474 |
| **PRS: PRSWEB_PHE** | 0.8052972789 | 0.07704299224 |
| **CDKN2C amplificati** | 0.19279148 | 0.07822485453 |
| **PRS: PMID2786325** | 0.8344714008 | 0.07826029488 |
| **FANCG deletion** | 1.599678077 | 0.07850422435 |
| **GATA3 deletion** | 0.4902290496 | 0.07905322108 |
| **PRSS1 amplificatior** | 0.347412307 | 0.08135115968 |
| **LMO3 deletion** | 2.568873752 | 0.08185959706 |
| **PRKAR1A amplifica** | 0.5315628231 | 0.08349579877 |
| **PRS: UKB_460K.pig** | 1.243722681 | 0.08383145173 |

| | | |
|---|---|---|
| **HLA_B_4901** | 2.308193489 | 0.08523468158 |
| **PTK2 amplification** | 0.4946650055 | 0.08544568222 |
| **AKT3 deletion** | 10.58248775 | 0.08609381146 |
| **HLA_DQA1_0101** | 1.448295165 | 0.08654387136 |
| **H3F3A deletion** | 10.16762276 | 0.08761854298 |
| **SNV in ATRX** | 5.329002489 | 0.08822866096 |
| **HLA_B_4402** | 1.741640375 | 0.08914783597 |
| **RET amplification** | 0.06368784432 | 0.08926650942 |
| **Lab value: PTI** | 4.806807542 | 0.08972257681 |
| **NFKBIZ deletion** | 0.3655016423 | 0.08977617148 |
| **GATA6 amplificatior** | 0.3664670786 | 0.08996892522 |
| **PRS: UKB_460K.pig** | 1.288058711 | 0.090142109 |
| **EZH2 amplification** | 0.557674872 | 0.09059778242 |
| **STAT6 amplification** | 1.602141793 | 0.09080937937 |
| **CDKN2B amplificati** | 0.108056249 | 0.0914153669 |
| **UROD deletion** | 2.481003468 | 0.09345393877 |
| **PRS: MEDS_PMID3** | 1.209619647 | 0.09357057229 |
| **XRCC2 amplificatio** | 0.3724134655 | 0.09371341093 |
| **H19 deletion** | 2.831319992 | 0.094255384 |
| **CD79B amplificatior** | 0.5406625004 | 0.0950325489 |
| **RAD54B deletion** | 8.128090453 | 0.0961090609 |
| **PNKP deletion** | 3.738719085 | 0.0964542084 |
| **PAXIP1 amplificatio** | 0.3765426628 | 0.09651399535 |
| **SNV in PTEN** | 0.1095280502 | 0.09720108059 |
| **BRCA1 deletion** | 2.240355019 | 0.09808250266 |
| **Lab value: DMNEUT** | 0.9401686264 | 0.09962000499 |
| **FGFR4 amplificatior** | 1.856037611 | 0.100754695 |
| **PAX5 deletion** | 1.55975703 | 0.1013987112 |
| **NOTCH2 amplificati** | 0.4384470419 | 0.1016554388 |
| **RAD54B amplificati** | 0.5208354521 | 0.1017498174 |
| **SNV in SMARCA4** | 6.421912533 | 0.1030657903 |
| **PIK3CA deletion** | 0.248452819 | 0.1037086854 |
| **SS18 deletion** | 2.501580408 | 0.1044256229 |
| **ZNF708 deletion** | 2.485487631 | 0.1055731267 |
| **SOX2 deletion** | 0.2126715635 | 0.1062244564 |
| **TAL1 deletion** | 2.325867004 | 0.1062909055 |
| **SMARCE1 deletion** | 3.438469465 | 0.1068030433 |
| **SNV in CDKN2A** | 2.118233133 | 0.1073466443 |
| **Lab value: TRIG** | 1.012839638 | 0.1086882129 |
| **ARID1A amplificatio** | 0.001439682594 | 0.1097630862 |
| **MAP2K1 deletion** | 0.3839499477 | 0.109966931 |
| **MDM4 deletion** | 8.452981815 | 0.1110168502 |
| **SNV in BCORL1** | 11.55405179 | 0.1115742241 |

| | | |
|---|---|---|
| **RNF8 amplification** | 0.4579930383 | 0.1125851536 |
| **UBE2T amplificatio** | 0.4993317583 | 0.1128806326 |
| **TMB_binned2** | 1.444272276 | 0.1152372152 |
| **PRS: PMID2786325** | 1.172975511 | 0.1167462006 |
| **HLA_B_0801** | 0.6811132887 | 0.1170249746 |
| **CDKN2A amplificati** | 0.03431131703 | 0.1175979173 |
| **FANCA deletion** | 0.5427618478 | 0.1191636076 |
| **PRS: UKB_460K.blo** | 1.187820096 | 0.1194976189 |
| **SNV in ERBB4** | 2.734600831 | 0.1203804142 |
| **BRCA1 amplificatio** | 0.4365078861 | 0.1206084923 |
| **SNV in B2M** | 3.446887976 | 0.121491715 |
| **Lab value: AMY** | 1.014124515 | 0.1215101605 |
| **PRS: CKD_overall_E** | 1.43410216 | 0.1232957887 |
| **PRS: UKB_460K.lun** | 0.8415881278 | 0.123581724 |
| **FH deletion** | 8.971862957 | 0.1244166929 |
| **SNV in KDM6A** | 2.497325248 | 0.1248416694 |
| **BRAF deletion** | 0.2719016698 | 0.1257642374 |
| **HOXB13 deletion** | 4.113098928 | 0.1260492115 |
| **CYLD deletion** | 0.5469151155 | 0.1263361435 |
| **NFKBIA deletion** | 1.730224382 | 0.1277828652 |
| **TMB** | 1.023056694 | 0.1283360624 |
| **CTCF amplification** | 4.966895085 | 0.1283992483 |
| **totalper_genes** | 0.24529636 | 0.1302080162 |
| **PRS: PRSWEB_PHE** | 1.173344237 | 0.1309304075 |
| **RUNX1T1 amplificat** | 0.5263056705 | 0.1309441763 |
| **DAXX amplification** | 0.4874495042 | 0.1328429064 |
| **ERBB3 amplificatio** | 1.526492431 | 0.1338525222 |
| **CBFB amplification** | 4.758534114 | 0.134543108 |
| **ABL1 deletion** | 1.669055611 | 0.1351306811 |
| **PRS: UKB_460K.rep** | 0.8629191045 | 0.1352651271 |
| **SDHAF2 amplificati** | 0.08405596059 | 0.1357863283 |
| **MYCL deletion** | 2.336241655 | 0.1364824274 |
| **PRS: UKB_460K.bo** | 1.188626429 | 0.1366484401 |
| **ATR amplification** | 0.1338770307 | 0.1375638043 |
| **RMRP amplification** | 0.1882802466 | 0.1376978016 |
| **UROD amplification** | 0.03727998327 | 0.1389957678 |
| **NF2 amplification** | 0.2513317265 | 0.1390132213 |
| **POLQ deletion** | 0.3833246686 | 0.1391742322 |
| **RPTOR deletion** | 6.261418414 | 0.1398083995 |
| **PRS: PMID2786325** | 1.149336033 | 0.1399370578 |
| **POT1 amplification** | 0.4182666293 | 0.1402582878 |
| **CBLB deletion** | 0.4079493278 | 0.1406624059 |
| **PRS: UKB_460K.ca** | 0.8819937605 | 0.1422306834 |

| | | |
|---|---|---|
| **SPOP deletion** | 3.751507309 | 0.1424629366 |
| **RAD51D amplificati** | 0.3745611711 | 0.1430658689 |
| **ARID1A deletion** | 0.5502556849 | 0.1442037099 |
| **RET deletion** | 0.5770640521 | 0.1463767836 |
| **SMARCB1 amplifica** | 0.3782417814 | 0.1479160453 |
| **MAPK1 amplificatio** | 0.3301999589 | 0.1479672587 |
| **SMAD4 amplificatio** | 0.3532792711 | 0.1492929281 |
| **HLA_DQB1_0602** | 1.436659576 | 0.1504885309 |
| **ETV4 amplification** | 0.4714068449 | 0.1512499896 |
| **BRE amplification** | 0.06935156907 | 0.1517987489 |
| **ERBB2 amplificatio** | 0.5333844239 | 0.1523638703 |
| **MYB amplification** | 0.5020951577 | 0.15272354 |
| **TET1 deletion** | 0.4695766784 | 0.1532651135 |
| **IGF2 deletion** | 2.454389384 | 0.1537131707 |
| **SF1 deletion** | 0.3451128123 | 0.1537223208 |
| **MECOM deletion** | 0.03551938671 | 0.1537515815 |
| **PTEN amplification** | 0.00174013901 | 0.1541756847 |
| **KIF1B amplification** | 0.01334552323 | 0.1546832667 |
| **MBD4 deletion** | 0.2972238541 | 0.1554024385 |
| **PRF1 deletion** | 0.6179517029 | 0.1569198945 |
| **ARID2 amplificatio** | 1.575690127 | 0.1581922909 |
| **BCL6 deletion** | 0.3034331679 | 0.159388803 |
| **SMO deletion** | 0.3060760468 | 0.1605972785 |
| **TERC amplification** | 0.5571226539 | 0.1610564139 |
| **PVRL4 amplificatio** | 0.5508922656 | 0.1610579538 |
| **RHPN2 deletion** | 5.990116226 | 0.1637419224 |
| **RHEB amplification** | 0.6129913209 | 0.1639162892 |
| **PRS: TRICL.sma.HM** | 1.191992733 | 0.1647178931 |
| **CDK8 deletion** | 0.3796539909 | 0.165737541 |
| **CIC amplification** | 0.3556205111 | 0.1659536307 |
| **NRAS amplification** | 0.3504256822 | 0.1672004304 |
| **STAT3 amplification** | 0.5146224695 | 0.1672304665 |
| **GATA2 deletion** | 0.4043847955 | 0.1690481716 |
| **MLH1 amplification** | 2.321758929 | 0.1694052013 |
| **CDH1 deletion** | 0.5253432953 | 0.1700091104 |
| **PPP2R1A deletion** | 3.196683391 | 0.1709527724 |
| **IGF1R deletion** | 0.4502363783 | 0.1711090161 |
| **PRS: PRSWEB_PHE** | 0.8626338142 | 0.1712469011 |
| **TMB_rank** | 1.15356967 | 0.1716284338 |
| **MYCL amplification** | 0.3275646431 | 0.171919864 |
| **GEN1 amplification** | 0.2459840174 | 0.1744705073 |
| **YAP1 deletion** | 0.3925406233 | 0.176567283 |
| **RMRP deletion** | 1.680271351 | 0.1766908652 |

| | | |
|---|---|---|
| **GATA6 deletion** | 1.693509709 | 0.1767138417 |
| **PTPN11 amplificatio** | 1.548353912 | 0.1773709269 |
| **KMT2D amplificatio** | 3.194497226 | 0.1776008002 |
| **GLI1 amplification** | 1.469259454 | 0.1798546336 |
| **XRCC1 amplificatio** | 0.3937971758 | 0.1798761458 |
| **MUTYH amplificatio** | 0.3035189172 | 0.1828414911 |
| **HLA_A_0201** | 1.256948489 | 0.1828729884 |
| **SOX9 amplification** | 0.6482161297 | 0.1829474575 |
| **HLA_DPB1_0101** | 0.5019007254 | 0.1833707199 |
| **HLA_C_0401** | 0.7308478156 | 0.1834704265 |
| **MAX deletion** | 2.494359067 | 0.1849259964 |
| **HLA_DQB1_0501** | 1.347249532 | 0.1867637441 |
| **CREBBP deletion** | 0.2479112195 | 0.1869932807 |
| **PRS: Stratified_left** | 0.8621703175 | 0.187391747 |
| **HLA_C_0701** | 0.761855325 | 0.1881304 |
| **STAG1 amplification** | 1.518132386 | 0.1882544238 |
| **PRS: Stratified_rect** | 0.8711488284 | 0.1893136414 |
| **ETV5 deletion** | 0.3068740478 | 0.1898332532 |
| **HLA_DRB1_0101** | 1.529235284 | 0.1900012535 |
| **PRS: PMID2786325** | 0.858067108 | 0.1907018167 |
| **HLA_A_2902** | 1.528584742 | 0.1914934029 |
| **SUZ12 amplification** | 0.5778248801 | 0.1916364931 |
| **GREM1 amplificatio** | 0.0110833975 | 0.1916433267 |
| **EME1 deletion** | 3.104781765 | 0.1920163715 |
| **TSC2 deletion** | 0.2707713445 | 0.1921472657 |
| **NTRK1 amplification** | 0.7060782426 | 0.1928150197 |
| **HLA_C_0202** | 1.598131492 | 0.1938864192 |
| **ROS1 deletion** | 0.630535872 | 0.194170538 |
| **DEPDC5 deletion** | 2.243914538 | 0.1941861839 |
| **RPA1 amplification** | 3.18729498 | 0.1947149283 |
| **FANCF amplification** | 0.1223561403 | 0.1947546752 |
| **PRS: TRICL.ade_ns.** | 1.199567794 | 0.1955126656 |
| **NF1 deletion** | 1.838350842 | 0.1956465004 |
| **PRS: UKB_460K.dis** | 0.8683058343 | 0.196966171 |
| **ID3 amplification** | 0.4049863934 | 0.197104553 |
| **REL deletion** | 0.0006961226388 | 0.1989607591 |
| **ZNRF3 amplification** | 0.2772878926 | 0.1990098328 |
| **PRPF8 amplification** | 0.1857966635 | 0.199074343 |
| **HLA_B_1402** | 1.537832141 | 0.1993132769 |
| **TCEB1 amplification** | 0.5998764381 | 0.1995888387 |
| **HLA_A_3101** | 1.824273385 | 0.2007150457 |
| **TERC deletion** | 0.05472630791 | 0.2017791731 |
| **PRS: PRSWEB_PHE** | 1.137170847 | 0.2019774698 |

| | | |
|---|---|---|
| **RHEB deletion** | 0.3114108604 | 0.2022046408 |
| **FAM46C amplificati** | 0.3979824806 | 0.2023673531 |
| **Lab value: T3TM** | 0.9857927273 | 0.2030590469 |
| **PIM1 amplification** | 0.6362995815 | 0.2052771508 |
| **HLA_B_4002** | 6.558420527 | 0.2057575065 |
| **SETBP1 amplificatio** | 0.4360019955 | 0.2067685785 |
| **EWSR1 amplificatio** | 0.3265596262 | 0.207130614 |
| **ALOX12B deletion** | 1.640316725 | 0.2087395263 |
| **CDK12 deletion** | 2.329741592 | 0.209885595 |
| **CHEK2 amplificatio** | 0.3903475662 | 0.2102713781 |
| **CEBPA amplificatio** | 0.6364926857 | 0.211004113 |
| **PRS: Stratified_LvR** | 1.144468058 | 0.2110229487 |
| **BRIP1 amplification** | 0.6482360311 | 0.2126890974 |
| **SMARCE1 amplifica** | 0.4332296015 | 0.2129823208 |
| **MEN1 amplification** | 0.1646257654 | 0.2130637651 |
| **B2M amplification** | 0.2756965046 | 0.2150508773 |
| **PMS2 deletion** | 2.777890087 | 0.2157152923 |
| **PRKAR1A deletion** | 3.915924132 | 0.2161219474 |
| **CBL deletion** | 0.6644547475 | 0.2166429075 |
| **CDKN1A amplificati** | 0.6364013107 | 0.2168012549 |
| **EGLN1 amplificatio** | 0.5876667692 | 0.2173660257 |
| **SRSF2 amplificatio** | 0.6393551259 | 0.2180862647 |
| **IL7R amplification** | 1.885445968 | 0.2196269938 |
| **PRS: Stratified_colo** | 0.8752067203 | 0.2200828657 |
| **HLA_DRB1_1501** | 1.429325806 | 0.2208034575 |
| **HLA_DQA1_0201** | 1.338722663 | 0.2210153176 |
| **CCND3 amplificatio** | 0.6909635657 | 0.2216589664 |
| **IDH2 deletion** | 0.5118885114 | 0.2218136641 |
| **BRCA2 amplificatio** | 0.3252875255 | 0.224431562 |
| **CCNE1 deletion** | 1.928305669 | 0.2266026489 |
| **PAX5 amplification** | 0.5061014664 | 0.2272899953 |
| **RPTOR amplificatio** | 0.5651866969 | 0.2286951223 |
| **GLI2 amplification** | 0.3622943525 | 0.2290398712 |
| **BLM deletion** | 0.5034632384 | 0.2291294737 |
| **PRPF40B amplificat** | 1.534085063 | 0.2292752899 |
| **MAF amplification** | 4.660794803 | 0.229486636 |
| **NTRK3 deletion** | 0.5122936381 | 0.2300958888 |
| **RAD51D deletion** | 2.307304709 | 0.2301724051 |
| **PRS: UKB_460K.car** | 0.878891924 | 0.2310632205 |
| **RICTOR amplificatio** | 1.886558832 | 0.2316008528 |
| **TLR4 deletion** | 1.698800929 | 0.2319623216 |
| **CBFA2T3 amplificat** | 5.098910277 | 0.2323802381 |
| **Lab value: PBUN** | 1.016799899 | 0.2329235651 |

| | | |
|---|---|---|
| **IDH1 deletion** | 1.478981464 | 0.2329833617 |
| **TAL1 amplification** | 0.01141605709 | 0.2337506271 |
| **PRS: BUN_overall_E** | 1.134236219 | 0.2341532018 |
| **PRS: PRSWEB_PHE** | 0.8667443347 | 0.2380879962 |
| **HLA_B_5001** | 3.619382843 | 0.2380911714 |
| **HIST1H3C amplifica** | 0.6136380626 | 0.2385385684 |
| **RAD52 deletion** | 0.2930095315 | 0.239925274 |
| **CRTC1 deletion** | 2.533768074 | 0.240197143 |
| **RICTOR deletion** | 0.4439647992 | 0.2408141677 |
| **Lab value: ALB** | 1.413414323 | 0.2415209581 |
| **WRN amplification** | 0.5140993137 | 0.2419333864 |
| **NTHL1 deletion** | 0.2156312892 | 0.2426035199 |
| **RPL26 deletion** | 1.638679588 | 0.2431393621 |
| **SS18 amplification** | 0.2324992802 | 0.2451459214 |
| **ZRSR2 deletion** | 0.1061392361 | 0.2459343389 |
| **KDM6B deletion** | 1.571125973 | 0.2474774695 |
| **CDK9 deletion** | 1.595215055 | 0.2475485482 |
| **ALK deletion** | 0.008833916945 | 0.2475749413 |
| **SDHB deletion** | 0.6447396782 | 0.2481693415 |
| **Lab value: PGLU** | 0.9954286728 | 0.2489313598 |
| **SMO amplification** | 0.6694423972 | 0.2509429359 |
| **HLA_C_0802** | 1.45584011 | 0.251034758 |
| **KAT6B deletion** | 0.5559874494 | 0.2511556602 |
| **BRD3 deletion** | 2.27918948 | 0.252654871 |
| **HLA_C_1601** | 1.771530996 | 0.252994078 |
| **TOPBP1 deletion** | 0.354448518 | 0.2531991572 |
| **ID3 deletion** | 0.6336896651 | 0.2536887536 |
| **GATA4 amplificatior** | 0.5051647277 | 0.2538558899 |
| **Lab value: AEOSM** | 28.1998513 | 0.2556530849 |
| **Lab value: CRE** | 1.20659424 | 0.2561325244 |
| **SYK deletion** | 1.582180625 | 0.2573261421 |
| **BARD1 deletion** | 2.033946718 | 0.2575302388 |
| **RBBP8 amplificatior** | 0.2322776158 | 0.2583919427 |
| **Lab value: VCRE** | 1.09072988 | 0.2603510649 |
| **XRCC5 deletion** | 2.021959034 | 0.260606871 |
| **CTNNA1 deletion** | 0.432869805 | 0.2618629417 |
| **POLQ amplification** | 0.2330523898 | 0.2622089858 |
| **POLE deletion** | 3.137214684 | 0.2628626173 |
| **SDHB amplification** | 0.3113225662 | 0.2648357295 |
| **HLA_B_3508** | 0.08878911836 | 0.2652012983 |
| **CDH4 deletion** | 3.772872799 | 0.2652429426 |
| **NFE2L2 deletion** | 0.5187649092 | 0.2655119898 |
| **SH2B3 amplificatior** | 1.450715537 | 0.2661680995 |

| | | |
|---|---|---|
| **STAT3 deletion** | 1.697637791 | 0.2662446402 |
| **ERBB4 deletion** | 1.409914782 | 0.2668098865 |
| **TCF7L1 amplificatio** | 2.129294655 | 0.2675246938 |
| **PRS: UKB_460K.blo** | 0.8825458153 | 0.2679902502 |
| **ERCC1 amplificatio** | 0.4509665465 | 0.2689245489 |
| **PRS: UKB_460K.dis** | 1.136204967 | 0.269775985 |
| **PRS: TRICL.nonsmo** | 1.142426559 | 0.2703414859 |
| **EZH2 deletion** | 0.3292096985 | 0.2718160325 |
| **BMPR1A amplificati** | 0.05028226249 | 0.2724784332 |
| **PRS: MEDS_PMID3** | 0.8872572105 | 0.2735737476 |
| **TLX3 amplification** | 1.862480248 | 0.2751978934 |
| **SMAD2 amplificatio** | 0.4869679903 | 0.2756273123 |
| **HNF1A deletion** | 1.681430628 | 0.277234679 |
| **NPRL3 deletion** | 0.3156146521 | 0.2775444034 |
| **FAS amplification** | 0.002485563618 | 0.2776306504 |
| **CCND1 deletion** | 0.3209625121 | 0.2783781537 |
| **Lab value: TP** | 0.7870246319 | 0.2791211953 |
| **Lab value: EGFR** | 0.9917603753 | 0.2791809483 |
| **PRS: PRSWEB_PHE** | 1.120260115 | 0.2802865687 |
| **MET deletion** | 0.4098036097 | 0.2803308685 |
| **SNV in TSC1** | 5.949158491 | 0.2807935251 |
| **CADM2 deletion** | 0.5440864788 | 0.2810195963 |
| **FGFR1 deletion** | 1.373848304 | 0.2815100984 |
| **SQSTM1 deletion** | 1.872703081 | 0.2821977343 |
| **KDM6A amplificatio** | 0.06604511659 | 0.2823198986 |
| **JAK1 amplification** | 0.02660882724 | 0.2834691678 |
| **FANCE amplificatio** | 0.6808762457 | 0.2835410997 |
| **ERCC1 deletion** | 2.860465194 | 0.2835828134 |
| **CSF3R amplificatio** | 0.3919159337 | 0.2847950311 |
| **RINT1 amplification** | 0.5406436159 | 0.2855681872 |
| **NFKBIE amplificatio** | 0.6545895035 | 0.2859236807 |
| **NF1 amplification** | 0.5921352188 | 0.2862547481 |
| **DIS3L2 deletion** | 1.950025707 | 0.2864633189 |
| **RB1 deletion** | 0.6739276861 | 0.2889180468 |
| **TOPBP1 amplificati** | 0.2951863387 | 0.2900227785 |
| **ARHGEF12 amplific** | 0.1969998029 | 0.290443196 |
| **CDK6 amplification** | 0.7066298676 | 0.2905802368 |
| **HMBS amplification** | 0.1974054411 | 0.2910567662 |
| **BUB1B amplificatio** | 0.4054664182 | 0.2918036014 |
| **CDK2 amplification** | 1.481899589 | 0.2918657374 |
| **PRS: RCC** | 0.8779401163 | 0.2920411154 |
| **EPHA7 deletion** | 0.4346586675 | 0.2924940144 |
| **SLX1B deletion** | 2.416620278 | 0.2938313136 |

| | | |
|---|---|---|
| **SNV in VHL** | 2.067500268 | 0.2939917781 |
| **SDHD deletion** | 0.7263936587 | 0.2942849462 |
| **SNV in BRCA2** | 2.323958942 | 0.2949855337 |
| **Lab value: MMONO** | 1.122978935 | 0.2950423369 |
| **PRS: UKB_460K.blo** | 0.8944796799 | 0.2956965045 |
| **MTOR deletion** | 0.675797022 | 0.2961210148 |
| **STAG2 amplificatior** | 0.1345420099 | 0.29690737 |
| **H3F3B amplificatior** | 0.6170508574 | 0.2974494248 |
| **CUX1 amplification** | 0.6902845296 | 0.2984116046 |
| **PALB2 deletion** | 0.3129717277 | 0.2989837108 |
| **PRS: PMID2786325** | 0.8871690128 | 0.2997668118 |
| **CCND1 amplificatio** | 0.6677919091 | 0.2997836927 |
| **DDR2 deletion** | 4.681141837 | 0.3007882406 |
| **SDHC deletion** | 4.681141837 | 0.3007882406 |
| **RARA amplification** | 0.6181391064 | 0.3029813357 |
| **PDGFRB deletion** | 0.6181919952 | 0.3040485368 |
| **ATR deletion** | 0.4085612464 | 0.3041494179 |
| **KIT deletion** | 1.476884354 | 0.3050158413 |
| **KLLN amplification** | 0.02941788368 | 0.305202283 |
| **CDKN1B deletion** | 1.443462893 | 0.305778283 |
| **QKI amplification** | 0.3106271335 | 0.3059936392 |
| **Lab value: UWBC** | 0.9531313778 | 0.306153542 |
| **SNV in MSH2** | 6.29106312 | 0.3076423905 |
| **SMC3 amplification** | 0.001200544225 | 0.3088032685 |
| **HRAS deletion** | 0.5946840595 | 0.3088775634 |
| **RHOT1 amplificatior** | 0.5862985839 | 0.3103892519 |
| **PRS: PMID2786325** | 0.8805437272 | 0.3107462782 |
| **PDGFRA deletion** | 1.47164388 | 0.3119178987 |
| **ELANE deletion** | 0.2334155325 | 0.3122181246 |
| **IL7R deletion** | 0.4888053919 | 0.3126295858 |
| **RAD50 deletion** | 0.4798768231 | 0.3142875441 |
| **CALR deletion** | 0.1025326734 | 0.3145532931 |
| **ARID1B amplificatio** | 0.3364351264 | 0.3145899358 |
| **ERCC6 amplificatio** | 0.1329991098 | 0.3148190819 |
| **GSTM5 deletion** | 0.533391705 | 0.3186599725 |
| **PHF6 deletion** | 0.1732884207 | 0.3196527458 |
| **PRS: PRSWEB_PHE** | 1.122051841 | 0.321794617 |
| **ETV1 deletion** | 2.328230441 | 0.3225713503 |
| **PARK2 amplificatior** | 0.3543322699 | 0.3226638322 |
| **CRLF2 deletion** | 0.2752381409 | 0.3234114981 |
| **RAD51C deletion** | 2.726166473 | 0.3235130846 |
| **SF3B1 amplification** | 0.6092216849 | 0.3239878237 |
| **SLC25A13 amplifica** | 0.59926649 | 0.3248894161 |

| | | |
|---|---|---|
| **Lab value: MLYMPH** | 1.045190513 | 0.324920372 |
| **PTPN14 amplificatio** | 0.6728987111 | 0.3258147596 |
| **EME1 amplification** | 0.5803683583 | 0.3264963119 |
| **PTPN11 deletion** | 1.618525976 | 0.3266841724 |
| **MAP2K2 amplificati** | 0.1717437309 | 0.3270023534 |
| **KLLN deletion** | 0.6251486351 | 0.3282794653 |
| **PPM1D deletion** | 2.879977866 | 0.3284965639 |
| **CDK1 deletion** | 0.559924387 | 0.3286557627 |
| **PDGFRB amplificati** | 1.64441642 | 0.3287811128 |
| **PRS: MEDS_PMID3** | 1.112738751 | 0.329212862 |
| **STAG2 deletion** | 0.09880030443 | 0.3301297121 |
| **GALNT12 deletion** | 2.072150907 | 0.3303212548 |
| **Lab value: SGPT** | 0.9945987103 | 0.3304923334 |
| **SUFU amplification** | 0.002262188989 | 0.3306968549 |
| **PRS: UKB_460K.cov** | 0.8686280379 | 0.3308994821 |
| **ATM amplification** | 0.2640272499 | 0.3323108148 |
| **BUB1B deletion** | 0.7001504271 | 0.3328468371 |
| **POLH amplification** | 0.6835957882 | 0.3328521592 |
| **PRS: UKB_460K.bo** | 1.118670603 | 0.3332091459 |
| **POT1 deletion** | 0.3641115218 | 0.333529881 |
| **PRF1 amplification** | 0.1054682565 | 0.3337005788 |
| **KDR deletion** | 1.433292288 | 0.3340614753 |
| **CBL amplification** | 0.3652875708 | 0.335233964 |
| **Lab value: MONO** | 1.034242597 | 0.3357354663 |
| **SLX4 deletion** | 0.08374491936 | 0.335764289 |
| **BCORL1 deletion** | 0.1480888447 | 0.3360260228 |
| **SETD2 amplificatio** | 1.853584451 | 0.3366910022 |
| **FOXL2 amplificatio** | 0.358563273 | 0.3369025866 |
| **BRD3 amplification** | 0.4712079691 | 0.3370576533 |
| **NOTCH3 deletion** | 0.02247080579 | 0.3371118783 |
| **AR deletion** | 0.0534364609 | 0.3372021117 |
| **PRS: PRSWEB_PHE** | 1.113207603 | 0.33827655 |
| **MET amplification** | 0.7312530742 | 0.338780465 |
| **FAH deletion** | 0.3910636024 | 0.3389550694 |
| **PRS: Stratified_mal** | 0.9009270913 | 0.3405364733 |
| **HNF1A amplificatio** | 1.372195223 | 0.341072612 |
| **PNRC1 deletion** | 0.4590608012 | 0.3412096994 |
| **TRAF7 deletion** | 0.09000721047 | 0.3421471345 |
| **PMS2 amplification** | 0.772565303 | 0.3422495631 |
| **RSPO3 amplificatio** | 2.079740844 | 0.3428158115 |
| **TET2 deletion** | 1.347325682 | 0.3433967255 |
| **TRAF7 amplificatio** | 2.049248967 | 0.3436510248 |
| **PSMD13 deletion** | 0.01399546177 | 0.3439108357 |

| | | |
|---|---|---|
| PRS: PMID27863252 | 0.8911789398 | 0.344315832 |
| HLA_DRB1_0701 | 1.256677366 | 0.3444227705 |
| IDH2 amplification | 0.5571700928 | 0.3444683195 |
| FUS deletion | 0.2494557579 | 0.345168764 |
| HLA_C_0303 | 0.7012153858 | 0.3461249075 |
| RHOH deletion | 1.665784986 | 0.3462130756 |
| PRS: PMID27863252 | 0.8917221619 | 0.3473208156 |
| PRS: TRICL.ade1_ns | 1.132411648 | 0.3480398954 |
| USP8 deletion | 0.6131346343 | 0.3480618951 |
| CDK12 amplification | 0.5704370042 | 0.349327255 |
| SNV in SMAD4 | 0.08730591715 | 0.350308733 |
| SNV in ASXL1 | 1.984344666 | 0.3503791053 |
| XRCC3 amplification | 0.1547168152 | 0.3504840144 |
| Lab value: PLT | 0.9988997381 | 0.3514306569 |
| RB1 amplification | 0.4209950923 | 0.3517021565 |
| EP300 amplification | 0.6062135083 | 0.3523608262 |
| PIK3C2B deletion | 2.076009186 | 0.3523863044 |
| NEIL1 deletion | 0.4092816285 | 0.3525002001 |
| FAM175A deletion | 1.905151665 | 0.3527105366 |
| USP28 amplification | 0.2372668017 | 0.3528950819 |
| NEGR1 deletion | 0.4944538636 | 0.353010491 |
| GNAQ deletion | 1.345923348 | 0.3542108352 |
| H3F3A amplification | 0.7794786106 | 0.3547429855 |
| RNF43 deletion | 2.54695779 | 0.3551111576 |
| NEIL1 amplification | 2.127888014 | 0.3554772763 |
| PRS: PRSWEB_PHE | 0.9021185124 | 0.3561969058 |
| ARID1B deletion | 0.744572962 | 0.3563621008 |
| KCNIP1 deletion | 1.679275998 | 0.3574524124 |
| PRS: UKB_460K.dis | 0.8991843983 | 0.3580124024 |
| PRS: UKB_460K.pig | 1.10448834 | 0.3580569153 |
| ITK deletion | 0.4798005915 | 0.3591838549 |
| PRS: Stratified_prox | 0.9108582795 | 0.3594339847 |
| JAK2 deletion | 1.251844968 | 0.361054435 |
| Lab value: ALK | 0.9984385704 | 0.3618962762 |
| PRS: UKB_460K.bod | 0.9009687196 | 0.3622428533 |
| KMT2A amplificatio | 0.2748536636 | 0.3651064411 |
| HLA_DQB1_0604 | 1.690715928 | 0.3652046691 |
| EGFR deletion | 2.00869932 | 0.3669390586 |
| HLA_DQB1_0202 | 1.296332531 | 0.3684680889 |
| MITF amplification | 0.5890856881 | 0.3684910921 |
| CRKL amplification | 0.6550008539 | 0.370158984 |
| PNKP amplification | 0.2702876518 | 0.3701630769 |
| SNV in NF1 | 1.280860456 | 0.3709328261 |

| | | |
|---|---|---|
| **MUS81 amplificatio** | 0.202815878 | 0.371233371 |
| **SNV in FAT1** | 2.689319447 | 0.3716454742 |
| **PRS: BreastCancer_** | 1.13369127 | 0.3719538599 |
| **RUNX1 amplificatio** | 0.658065282 | 0.3720964851 |
| **NT5C2 amplificatior** | 0.0005644113314 | 0.3732765211 |
| **BCOR amplification** | 0.05948357644 | 0.3740710261 |
| **RHBDF2 amplificati** | 0.6633978279 | 0.3770512829 |
| **NF2 deletion** | 1.415891946 | 0.3794566227 |
| **TNFAIP3 amplificati** | 0.5791024045 | 0.3795783835 |
| **SNV in RB1** | 0.005466486977 | 0.3810461751 |
| **ATRX deletion** | 0.03851494441 | 0.3811961853 |
| **MSH6 amplification** | 0.611499279 | 0.3814496967 |
| **ERCC4 deletion** | 0.1870204644 | 0.3815531807 |
| **SNV in ATM** | 0.4481498317 | 0.3815697178 |
| **SDHC amplification** | 0.8001153276 | 0.3844762656 |
| **ENG amplification** | 0.5006230055 | 0.384740901 |
| **TRIM37 amplificatio** | 0.6622151557 | 0.3850178853 |
| **Lab value: URBC** | 1.052878965 | 0.3872658724 |
| **SNV in NOTCH1** | 1.683567984 | 0.3877095166 |
| **EWSR1 deletion** | 1.404040823 | 0.3878222149 |
| **CHEK1 amplificatio** | 0.198978903 | 0.3882435318 |
| **XRCC5 amplificatio** | 2.222432095 | 0.3888452148 |
| **HLA_C_0304** | 1.30700056 | 0.3890380071 |
| **TMB_binned1** | 0.7945635592 | 0.3891970577 |
| **ATM deletion** | 0.7740987304 | 0.3899897666 |
| **NPRL2 deletion** | 0.6660915765 | 0.3906113755 |
| **CHEK1 deletion** | 0.6422445544 | 0.3911012826 |
| **ABL1 amplification** | 0.5745809104 | 0.391400854 |
| **Lab value: FRT4M** | 1.836621986 | 0.3915633938 |
| **TCEB1 deletion** | 0.1095071809 | 0.3922367004 |
| **BAP1 amplification** | 0.3625213029 | 0.3927812127 |
| **CUX1 deletion** | 0.4685070477 | 0.3929519332 |
| **SMAD4 deletion** | 1.299008154 | 0.3932266344 |
| **HLA_B_1401** | 0.001954841398 | 0.3935513531 |
| **PRS: PMID2786325** | 1.09104877 | 0.3936464434 |
| **TAZ amplification** | 0.2924534457 | 0.3942537966 |
| **HFE amplification** | 0.6852424438 | 0.3960193741 |
| **PRS: PRSWEB_PHE** | 0.9160719334 | 0.3961214369 |
| **ALK amplification** | 0.6100214266 | 0.3974065009 |
| **CD274 deletion** | 1.231691098 | 0.3988140039 |
| **CDK4 deletion** | 1.532835304 | 0.3988517438 |
| **RUNX1T1 deletion** | 2.505844971 | 0.3988817539 |
| **NOTCH3 amplificati** | 0.08782611136 | 0.3992809704 |

| | | |
|---|---|---|
| **FBXW7 amplificatio** | 0.1933382885 | 0.3995237078 |
| **KDM5C amplificatio** | 0.1174761463 | 0.399606827 |
| **HLA_B_1501** | 0.6788343536 | 0.4006125205 |
| **GNB2L1 amplificati** | 2.130413888 | 0.4008833981 |
| **PRS: UKB_460K.rep** | 1.120997356 | 0.4018209575 |
| **HLA_A_3001** | 1.802785869 | 0.402112953 |
| **PHF6 amplification** | 0.3238813775 | 0.4021393675 |
| **GPC3 deletion** | 0.2252503707 | 0.4021500569 |
| **UIMC1 deletion** | 0.5186771598 | 0.4022817606 |
| **Lab value: GGTL** | 0.974619683 | 0.4044061122 |
| **NFKBIA amplificatio** | 0.6007467402 | 0.4049122519 |
| **RPA1 deletion** | 0.731689467 | 0.405505163 |
| **ARAF amplification** | 0.04278916491 | 0.4067397699 |
| **BABAM1 deletion** | 0.1621573857 | 0.4069662194 |
| **DDR2 amplification** | 0.8074966432 | 0.4074347286 |
| **RBL2 amplification** | 0.08882347368 | 0.4096201724 |
| **HLA_B_5101** | 0.6932806766 | 0.4111056418 |
| **EGLN1 deletion** | 3.686610908 | 0.411434798 |
| **FAN1 amplification** | 0.06368745459 | 0.412569869 |
| **ESR1 amplification** | 0.5839646959 | 0.4133107683 |
| **PRS: PMID2786325** | 0.9177833805 | 0.413329351 |
| **NT5C2 deletion** | 0.6962823718 | 0.4141880874 |
| **PRS: PRSWEB_PHE** | 0.9135887825 | 0.4162859749 |
| **DCLRE1C deletion** | 0.6905082046 | 0.4164473131 |
| **PRS: PRSWEB_PHE** | 0.9149945921 | 0.4174516312 |
| **CCNE1 amplificatio** | 0.7629518488 | 0.4181365788 |
| **MTAP deletion** | 0.7529348518 | 0.4183534194 |
| **DDB1 amplification** | 0.1904520844 | 0.4184645698 |
| **VEGFA amplificatior** | 0.7331866978 | 0.4185727779 |
| **PRS: UKB_460K.rep** | 1.096308459 | 0.4203316194 |
| **SH2B3 deletion** | 1.489388922 | 0.4207050125 |
| **ARHGEF12 deletion** | 0.6402357939 | 0.4207592199 |
| **HABP2 deletion** | 0.682382063 | 0.4217549095 |
| **SMARCB1 deletion** | 1.383359224 | 0.4224957407 |
| **AXL deletion** | 1.691725521 | 0.4234140407 |
| **TAL2 deletion** | 1.831501005 | 0.4239990411 |
| **MITF deletion** | 0.7622300268 | 0.4241152659 |
| **SNV in SETD2** | 1.491993814 | 0.4247949476 |
| **EPHA3 deletion** | 0.6890039338 | 0.4249086486 |
| **EPCAM amplificatio** | 1.956253194 | 0.4249666301 |
| **U2AF1 amplificatior** | 0.6214195074 | 0.4262389044 |
| **TSC1 amplification** | 0.6076141682 | 0.4262779301 |
| **AKT2 deletion** | 1.66415995 | 0.4263864762 |

| | | |
|---|---|---|
| **PRS: PMID27863252** | 0.9103739484 | 0.4277195902 |
| **CDK2 deletion** | 2.140105363 | 0.4284445577 |
| **RELA amplification** | 0.237530616 | 0.4285144717 |
| **PRS: PRSWEB_PHE** | 0.9106740424 | 0.4292077716 |
| **BRE deletion** | 0.0171086929 | 0.4299394471 |
| **APC amplification** | 1.603254505 | 0.4299457723 |
| **PRS: MEDS_PMID3** | 0.9163977161 | 0.430082262 |
| **CDC73 amplification** | 0.8164544443 | 0.4303360018 |
| **FLT3 deletion** | 0.7282594616 | 0.4326015241 |
| **IDH1 amplification** | 0.6125718233 | 0.4335419895 |
| **PRS: PMID27863252** | 0.923028857 | 0.4357013787 |
| **PRS: TRICL.ade.HM** | 0.9104779391 | 0.4361872281 |
| **Lab value: MEOS** | 1.297024852 | 0.4363400994 |
| **DDB2 amplification** | 0.2107723461 | 0.4365283618 |
| **KLF2 deletion** | 0.2290971297 | 0.4371737003 |
| **PRS: PMID27863252** | 0.9066257077 | 0.4374636943 |
| **HLA_DPB1_0402** | 0.7355995955 | 0.4376666465 |
| **MAP3K1 amplificati** | 1.622631438 | 0.4380519526 |
| **SDHD amplification** | 0.4471520925 | 0.4381623349 |
| **NSD1 deletion** | 0.5459883718 | 0.4401778072 |
| **RNF43 amplification** | 0.6787395513 | 0.4412293962 |
| **PARK2 deletion** | 0.7908906934 | 0.4413157368 |
| **FAT1 deletion** | 0.5497951782 | 0.4413957966 |
| **CHEK2 deletion** | 1.351107475 | 0.4433811771 |
| **PRS: UKB_460K.blo** | 1.093824042 | 0.4433824886 |
| **CARD11 deletion** | 1.863307334 | 0.4439837028 |
| **MYD88 amplificatio** | 1.546763423 | 0.4455969153 |
| **EXO1 deletion** | 3.490069952 | 0.4458428552 |
| **TCF3 deletion** | 0.6112553247 | 0.4463586209 |
| **NFE2L2 amplificatio** | 0.6605335653 | 0.446680591 |
| **RAD51C amplificati** | 0.6961043944 | 0.4469240525 |
| **PRS: PRSWEB_PHE** | 0.930056913 | 0.4478798625 |
| **NSD1 amplification** | 1.490597613 | 0.4485334012 |
| **BARD1 amplification** | 2.553053083 | 0.4488056368 |
| **ZNRF3 deletion** | 1.572331874 | 0.4488220997 |
| **SPOP amplification** | 0.6573359936 | 0.4505687428 |
| **Lab value: CEA** | 0.9513954206 | 0.4507646994 |
| **PRS: X20171017_MV** | 1.085273778 | 0.4518114703 |
| **CDK4 amplification** | 1.252661485 | 0.4520271662 |
| **PRS: PGS000194_pl** | 0.9207601597 | 0.4536327676 |
| **PRS: PGS000195_pl** | 0.9207601597 | 0.4536327676 |
| **PRS: PGS000196_pl** | 0.9207601597 | 0.4536327676 |
| **PRS: PGS000197_pl** | 0.9207601597 | 0.4536327676 |

| | | |
|---|---|---|
| **PRS: PGS000198_pl** | 0.9207601597 | 0.4536327676 |
| **PRS: PGS000199_pl** | 0.9207601597 | 0.4536327676 |
| **Lab value: MCV** | 1.012267503 | 0.4542981389 |
| **PRS: UKB_460K.dis** | 1.08430652 | 0.4548202703 |
| **MDM2 amplification** | 1.248432073 | 0.4557625292 |
| **SNV in ROS1** | 2.442877528 | 0.4571739809 |
| **PRS: PRSWEB_PHE** | 0.9188157453 | 0.4578563849 |
| **IKZF3 amplification** | 0.6053258359 | 0.4585442315 |
| **WHSC1 amplificatio** | 1.563680283 | 0.4591876629 |
| **SBDS deletion** | 0.486247859 | 0.4596616078 |
| **PPM1D amplificatio** | 0.6983609457 | 0.46033837 |
| **CYLD amplification** | 0.6003826911 | 0.4606137136 |
| **AXIN2 amplification** | 0.7015561816 | 0.4608124328 |
| **HABP2 amplification** | 0.009578735356 | 0.4613305723 |
| **CBFB deletion** | 0.6526433787 | 0.4621167953 |
| **JAZF1 deletion** | 0.04614446882 | 0.462192506 |
| **KLF4 deletion** | 1.662616415 | 0.4624839378 |
| **MAPK1 deletion** | 1.390076317 | 0.4625455931 |
| **PRS: UKB_460K.pig** | 1.124387197 | 0.4627762646 |
| **SH2D1A amplificati** | 0.278016956 | 0.4645039993 |
| **HMBS deletion** | 0.677814291 | 0.4659621493 |
| **WT1 amplification** | 0.3190673664 | 0.4663146154 |
| **CDK5 deletion** | 0.06285669593 | 0.4663966792 |
| **HLA_DRB1_0401** | 0.7544641441 | 0.4668592594 |
| **PML deletion** | 0.5014907613 | 0.4675133608 |
| **HIST1H3B amplifica** | 0.7325183135 | 0.4677048236 |
| **SNV in ARID1A** | 1.165697183 | 0.4680352196 |
| **RSPO3 deletion** | 0.752050842 | 0.4684456897 |
| **XPO1 amplification** | 0.6774288465 | 0.4696388592 |
| **TDG deletion** | 0.5467734124 | 0.4701329417 |
| **KDM6A deletion** | 0.2749417127 | 0.4727755253 |
| **TP53BP1 deletion** | 0.7064478194 | 0.4730749037 |
| **EXO1 amplification** | 0.7370747276 | 0.4742355682 |
| **ITK amplification** | 1.760467506 | 0.4744998009 |
| **ESR1 deletion** | 0.797282359 | 0.4761012958 |
| **FANCD2 amplificati** | 0.6595407777 | 0.476131827 |
| **SLX4 amplification** | 1.707588021 | 0.4775936513 |
| **TERT amplification** | 1.215329243 | 0.4778936174 |
| **CD58 deletion** | 0.648788785 | 0.4779631186 |
| **ETV1 amplification** | 0.8252992991 | 0.4781860949 |
| **PRS: UKB_460K.blo** | 1.09666187 | 0.4782447703 |
| **MGA deletion** | 0.713228832 | 0.4787152665 |
| **MYB deletion** | 0.7989924415 | 0.4789478299 |

| | | |
|---|---|---|
| **PRS: PMID2786325[illegible]** | 0.9300409783 | 0.4794882964 |
| **DEPDC5 amplificati** | 0.009277465758 | 0.4795708169 |
| **SNV in APC** | 1.303182924 | 0.4805563455 |
| **HLA_DRB1_0301** | 0.8469789734 | 0.480674541 |
| **FANCB deletion** | 0.2826160364 | 0.4815823902 |
| **SNV in TET2** | 2.123278272 | 0.4819772935 |
| **PHOX2B amplificati** | 0.5830680177 | 0.482015225 |
| **NOTCH1 amplificati** | 0.6162783957 | 0.4831254553 |
| **KEAP1 amplificatior** | 0.7025267022 | 0.4837355836 |
| **FLT1 deletion** | 0.7551756849 | 0.4839123044 |
| **TRAF3 amplificatior** | 0.1185543729 | 0.4845586594 |
| **Lab value: ABASO** | 16.23699443 | 0.4849639629 |
| **TRIM37 deletion** | 1.956431453 | 0.4853059064 |
| **SLX1A deletion** | 1.970505511 | 0.4860722389 |
| **LIG4 amplification** | 0.5750793387 | 0.4863785483 |
| **NTRK3 amplificatior** | 0.6150640731 | 0.4893593909 |
| **NEIL3 amplification** | 0.6308151731 | 0.4899612738 |
| **MEN1 deletion** | 0.591231799 | 0.4917849717 |
| **SMC3 deletion** | 0.7892933345 | 0.4920702843 |
| **PDCD1LG2 deletion** | 1.18681532 | 0.4932446322 |
| **HLA_C_1701** | 0.06511811093 | 0.4936692462 |
| **SUFU deletion** | 0.8070440657 | 0.4940441719 |
| **AKT3 amplification** | 0.8301198492 | 0.4975541126 |
| **PBRM1 amplificatio** | 0.442867119 | 0.4977732326 |
| **CEBPA deletion** | 1.469268012 | 0.4985805831 |
| **PRDM1 amplificatio** | 1.33074568 | 0.4991489037 |
| **PRS: Stratified_dist[illegible]** | 0.9256840765 | 0.4993417516 |
| **GPC3 amplification** | 0.3961807265 | 0.4994061633 |
| **KIF1B deletion** | 1.347804612 | 0.5000028785 |
| **SLITRK6 deletion** | 0.6726222648 | 0.5001047769 |
| **RAD51 deletion** | 0.7239500187 | 0.5003619231 |
| **TSHR deletion** | 0.5916363549 | 0.5016684531 |
| **HLA_C_0501** | 1.249941374 | 0.5028720915 |
| **EXT2 deletion** | 0.6646085542 | 0.5031339424 |
| **NTHL1 amplificatior** | 1.613824471 | 0.5033676543 |
| **SOX9 deletion** | 2.044981267 | 0.5034108629 |
| **CDKN1B amplificati** | 1.232709768 | 0.5037952382 |
| **TLX3 deletion** | 0.5974628962 | 0.5040226489 |
| **KMT2A deletion** | 0.7004692727 | 0.5045460356 |
| **SOCS1 deletion** | 0.6062023717 | 0.505273278 |
| **SRSF2 deletion** | 2.062162723 | 0.5063775333 |
| **MAP2K2 deletion** | 0.3553569386 | 0.5068916026 |
| **MRE11A amplificati** | 0.4650936023 | 0.5100149378 |

| | | |
|---|---|---|
| DIS3 amplification | 0.7145912364 | 0.5102303613 |
| GATA3 amplification | 0.6848014827 | 0.510373062 |
| KDM5A deletion | 0.5981454788 | 0.5111617684 |
| RARA deletion | 1.377430003 | 0.5118635414 |
| PRS: GSCAN.Drinks | 1.069156069 | 0.5123087978 |
| SNV in BCOR | 0.002869419024 | 0.5124516019 |
| USP28 deletion | 0.7160144998 | 0.5149284803 |
| PTPRD deletion | 1.283710955 | 0.5151089134 |
| ASXL1 deletion | 0.0003367107326 | 0.5160008858 |
| PRS: PMID27863252 | 0.9270204622 | 0.5165610507 |
| CSF3R deletion | 0.7404095517 | 0.5185626933 |
| DKC1 amplification | 1.733808434 | 0.5188521368 |
| AKT1 amplification | 1.340243434 | 0.5188977072 |
| HLA_C_0102 | 0.7629590591 | 0.5192547043 |
| HLA_B_3501 | 0.7280163771 | 0.5192721784 |
| PRS: PMID27863252 | 0.93789413 | 0.5196874083 |
| NFKBIZ amplificatio | 0.738063591 | 0.5197155167 |
| PRS: UKB_460K.cov | 1.072779234 | 0.5197595315 |
| PPP2R1A amplificat | 0.629938108 | 0.5198541171 |
| PRS: TRICL.sqc.HM | 1.101116885 | 0.5201444425 |
| LMO1 deletion | 1.334484535 | 0.5202749122 |
| MCL1 deletion | 2.518404057 | 0.5220322437 |
| SOX2 amplification | 1.182657875 | 0.5227926601 |
| AXL amplification | 0.7670315458 | 0.5243993315 |
| ERCC2 deletion | 0.6654094424 | 0.5248004753 |
| INSIG1 deletion | 0.1079199324 | 0.5255097735 |
| PRS: PRSWEB_PHE | 0.9376562361 | 0.5258484823 |
| MAF deletion | 0.7165442534 | 0.5267266937 |
| MAP2K1 amplificati | 0.6774841856 | 0.5278766873 |
| NEIL3 deletion | 0.5748475947 | 0.5301338164 |
| XRCC6 deletion | 0.5786658646 | 0.5307119204 |
| MDM4 amplification | 0.8521455334 | 0.5312310594 |
| TCF7L2 deletion | 0.80702615 | 0.5312815503 |
| ATRX amplification | 0.04433854743 | 0.5322049159 |
| PRS: GSCAN.AgeOf | 1.07894249 | 0.5324785012 |
| FOXA1 amplification | 1.395523417 | 0.5330693789 |
| Lab value: CA125 | 0.9979316624 | 0.5334206721 |
| MYBL1 deletion | 1.570166191 | 0.5338429611 |
| ENG deletion | 1.551285115 | 0.5342022731 |
| ZNF217 amplificatio | 0.8239505158 | 0.5347124293 |
| PRS: MEDS_PMID3 | 1.070955218 | 0.5352695118 |
| Lab value: MALYM | 1.938081042 | 0.5355580193 |
| PRAME amplificatio | 0.04765516035 | 0.5356273891 |

| | | |
|---|---|---|
| HLA_A_2402 | 0.8345949662 | 0.5356886036 |
| IGF1R amplification | 0.6692678808 | 0.5366177922 |
| TAL2 amplification | 0.6018361143 | 0.5373938214 |
| SF1 amplification | 0.0881491503 | 0.5374221784 |
| Lab value: NEUT | 0.9926974719 | 0.5376377104 |
| MTOR amplification | 0.6456125947 | 0.539514222 |
| IKZF3 deletion | 1.537603935 | 0.5403128788 |
| RIF1 deletion | 0.1614415145 | 0.5406804938 |
| NR0B1 amplificatio | 0.01998630971 | 0.5408549958 |
| PRPF8 deletion | 1.335992952 | 0.5411327122 |
| BMPR1A deletion | 0.8177301041 | 0.5415533143 |
| PRS: Asthma.adults | 0.9341774615 | 0.541598915 |
| XRCC4 deletion | 0.6837833174 | 0.5418443646 |
| PRS: BRCA.ERNEG | 1.069175946 | 0.5420088061 |
| EPHA7 amplificatio | 1.392389873 | 0.5443419458 |
| STAG1 deletion | 0.001259157264 | 0.5454174056 |
| TDG amplification | 1.383720319 | 0.5459963501 |
| VHL amplification | 0.7035537814 | 0.5463094039 |
| ARHGAP35 amplific | 1.582033396 | 0.5469783554 |
| TMEM127 amplifica | 0.5912726364 | 0.5471562682 |
| DMC1 amplification | 0.6987509462 | 0.5474961872 |
| PRS: PRSWEB_PHE | 0.9361774136 | 0.5475959512 |
| MAP3K1 deletion | 1.219561182 | 0.5476721902 |
| BRIP1 deletion | 1.776377975 | 0.5480725707 |
| PRS: MEDS_PMID3 | 1.077188726 | 0.5511773327 |
| RIF1 amplification | 2.17304831 | 0.5519262941 |
| VHL deletion | 0.7813170961 | 0.5526727993 |
| PTEN deletion | 0.823939506 | 0.5530535959 |
| RAF1 deletion | 0.7788948441 | 0.5535276066 |
| PRS: PRSWEB_PHE | 0.9286002573 | 0.5539864329 |
| PRS: PMID2786325 | 1.068843782 | 0.5540805273 |
| XRCC4 amplificatio | 1.675292228 | 0.5545578215 |
| PRS: PMID2786325 | 0.940589635 | 0.5548029795 |
| PRPF40B deletion | 1.664949875 | 0.5549296934 |
| HLA_DPB1_0301 | 1.211583537 | 0.5549851569 |
| MTA1 amplification | 0.3007602258 | 0.5551459532 |
| SLX1A amplificatior | 0.01604681575 | 0.5555184618 |
| DOCK8 amplificatio | 0.5877092035 | 0.5556246063 |
| MRE11A deletion | 0.6791278829 | 0.556094356 |
| PRS: UKB_460K.rep | 1.076255101 | 0.5574767048 |
| DMC1 deletion | 0.5730363937 | 0.5600138486 |
| FANCD2 deletion | 0.7857687454 | 0.5614771179 |
| RASA1 amplificatior | 1.676347348 | 0.5629063157 |

| | | |
|---|---|---|
| **PRS: PRSWEB_PHE** | 1.050689296 | 0.564728347 |
| **XRCC6 amplificatio** | 0.7041273681 | 0.5654386191 |
| **MTAP amplification** | 0.46297413 | 0.5660344763 |
| **KLF4 amplification** | 0.6472360984 | 0.5663772253 |
| **BRCA2 deletion** | 0.7987750693 | 0.5677069023 |
| **HLA_B_5301** | 0.002261315501 | 0.5680603323 |
| **PRS: UKB_460K.blo** | 0.9454199101 | 0.5684231884 |
| **INSIG1 amplificatio** | 1.299068151 | 0.5689994079 |
| **BCOR deletion** | 0.3055448326 | 0.5692694953 |
| **TNFAIP3 deletion** | 0.8355421096 | 0.5703460902 |
| **SDHAF2 deletion** | 0.6407202649 | 0.5706961886 |
| **Lab value: SGOT** | 0.9960510585 | 0.5723797929 |
| **SNV in ATR** | 1.598545501 | 0.5726649837 |
| **SUZ12 deletion** | 1.386199552 | 0.573595019 |
| **PRS: MEDS_PMID3** | 0.9429416836 | 0.5737514263 |
| **BCL2L12 amplificat** | 1.308539083 | 0.5777694117 |
| **FANCB amplificatio** | 0.07999436792 | 0.5793568691 |
| **Lab value: BASO** | 1.18097913 | 0.5793570545 |
| **RASA1 deletion** | 0.7029870203 | 0.5793665297 |
| **FLCN deletion** | 1.166420778 | 0.5794130618 |
| **FGFR1 amplificatio** | 0.8151892573 | 0.579486554 |
| **YAP1 amplification** | 0.5638351617 | 0.5801170596 |
| **MED12 deletion** | 0.1552334262 | 0.5802466536 |
| **PTPN14 deletion** | 2.114441914 | 0.5803140077 |
| **Lab value: DBILI** | 2.528704847 | 0.5803973242 |
| **PRS: OVCA** | 0.947836938 | 0.5813243828 |
| **PMS1 deletion** | 0.7411398001 | 0.5816487613 |
| **PRS: MEDS_PMID3** | 1.064154726 | 0.5816681207 |
| **PRS: MEDS_PMID3** | 0.9466331133 | 0.5833074114 |
| **APC deletion** | 0.8134171332 | 0.5834306688 |
| **POLD1 deletion** | 1.409252665 | 0.5834338802 |
| **LMO2 amplification** | 0.3713357461 | 0.5841328628 |
| **ERG amplification** | 0.6097268583 | 0.586669524 |
| **CALR amplification** | 0.07016401241 | 0.587508988 |
| **ERBB2 deletion** | 1.289055745 | 0.5885701841 |
| **PRS: PMID2786325** | 0.9457671905 | 0.5909800076 |
| **HLA_B_0702** | 1.139810092 | 0.5912217736 |
| **NTRK2 amplificatio** | 0.6816333534 | 0.5914324733 |
| **Lab value: CA199C** | 1.013585751 | 0.5941240986 |
| **BCORL1 amplificati** | 0.4072065752 | 0.5962584941 |
| **FGFR2 deletion** | 0.8463783559 | 0.5963784903 |
| **RUNX1 deletion** | 0.7346194973 | 0.5978012511 |
| **MSH2 amplification** | 0.7295212278 | 0.5981471989 |

| | | |
|---|---|---|
| **PRS: UKB_460K.car** | 1.071010618 | 0.5982883464 |
| **PPARG deletion** | 1.546433537 | 0.5996950656 |
| **HLA_DQB1_0201** | 0.8910561489 | 0.5998764778 |
| **PDCD1LG2 amplific** | 0.7929880988 | 0.6004427228 |
| **PRS: PMID27863252** | 0.9420663169 | 0.6004656114 |
| **BCL2 deletion** | 1.185030692 | 0.6005787358 |
| **FANCF deletion** | 1.281179346 | 0.6016080406 |
| **PRS: PMID27863252** | 1.055181513 | 0.6034929899 |
| **IGF2 amplification** | 3.044332531 | 0.605869979 |
| **Lab value: TSH** | 0.9737946027 | 0.6062001033 |
| **Lab value: CHOL** | 1.009974438 | 0.6074208365 |
| **H19 amplification** | 3.177488504 | 0.6075398865 |
| **FH amplification** | 0.8717710902 | 0.6087914722 |
| **SNV in ARID2** | 1.484151145 | 0.6096116912 |
| **PRS: UKB_460K.bod** | 1.062111741 | 0.6109504427 |
| **WRN deletion** | 1.1427041 | 0.6145724633 |
| **MEF2B deletion** | 1.316227464 | 0.615147737 |
| **MCM8 deletion** | 0.72962379 | 0.615352183 |
| **EPHA5 amplification** | 0.6852613425 | 0.6155874019 |
| **Lab value: MBASO** | 1.508101926 | 0.6164176284 |
| **FANCI deletion** | 0.6904673223 | 0.6175168796 |
| **SRC amplification** | 1.281366429 | 0.617944181 |
| **SMAD2 deletion** | 1.176934702 | 0.6189084534 |
| **PRS: PMID27863252** | 0.9493058291 | 0.6205465949 |
| **PAXIP1 deletion** | 0.6466359186 | 0.6209912421 |
| **HLA_B_5701** | 0.8225744599 | 0.622189905 |
| **CSF1R amplification** | 1.518201759 | 0.6247045196 |
| **FLT3 amplification** | 0.6338544172 | 0.6248942274 |
| **SF3B1 deletion** | 1.356353071 | 0.6265361141 |
| **Lab value: LYMPH** | 1.006285941 | 0.6281139903 |
| **CCND2 amplificatio** | 1.162923072 | 0.6303667251 |
| **RAC1 amplification** | 0.8420082372 | 0.630942074 |
| **PRS: PRSWEB_PHE** | 0.9505951131 | 0.6322857822 |
| **SNV in KMT2A** | 0.6171373722 | 0.6330293361 |
| **HLA_B_4501** | 0.0004885550283 | 0.6340372446 |
| **ZNF708 amplificatio** | 1.200685958 | 0.6346888338 |
| **Lab value: NRBC** | 0.5939603969 | 0.6361560435 |
| **COL7A1 amplificati** | 0.587359212 | 0.6372195151 |
| **NR0B1 deletion** | 0.2733920943 | 0.637294755 |
| **ARHGAP35 deletion** | 1.330262275 | 0.6380784007 |
| **KCNQ1 amplificatio** | 2.720453921 | 0.6386904943 |
| **PRS: TRICL.ade1.HI** | 0.9466606529 | 0.640362277 |
| **FAH amplification** | 1.525510205 | 0.640811819 |

| | | |
|---|---|---|
| **PRS: PRSWEB_PHE** | 1.05139931 | 0.6411449162 |
| **RBL2 deletion** | 0.7722444378 | 0.643035341 |
| **GLI2 deletion** | 0.6580090939 | 0.6436923047 |
| **PRS: MEDS_PMID3** | 1.049312694 | 0.6438673526 |
| **CRTC2 amplificatio** | 0.8649401883 | 0.6442297838 |
| **HLA_B_1801** | 0.8231449554 | 0.6445754453 |
| **PRS: CD_deLange2** | 1.061206252 | 0.6458796854 |
| **Lab value: AMONO** | 1.182327737 | 0.6463965072 |
| **MBD4 amplification** | 0.6916820046 | 0.6476777957 |
| **ZRSR2 amplificatio** | 0.6227799813 | 0.6483683384 |
| **PRKCZ amplificatio** | 1.507510064 | 0.6492185744 |
| **PRS: PRSWEB_PHE** | 1.046580651 | 0.6504872563 |
| **PRS: UKB_460K.bm** | 0.9539435636 | 0.6506309036 |
| **XRCC2 deletion** | 0.6609422098 | 0.65197013 |
| **HOXB13 amplificati** | 0.7827359667 | 0.6521909154 |
| **UBE2T deletion** | 2.01026691 | 0.6526984265 |
| **FANCI amplification** | 0.7006861495 | 0.6527687045 |
| **FAT1 amplification** | 0.7327094748 | 0.6535871052 |
| **PRS: PRSWEB_PHE** | 1.051161275 | 0.6548624488 |
| **CDKN1C amplificati** | 0.3671148131 | 0.6572930394 |
| **SLITRK6 amplificati** | 1.335235844 | 0.6578638352 |
| **MEF2B amplificatio** | 1.169569686 | 0.6589317564 |
| **BLM amplification** | 0.7586862965 | 0.6596722083 |
| **PRS: IBD_deLange2** | 0.9515871621 | 0.6609583495 |
| **WHSC1 deletion** | 1.29717279 | 0.6611449666 |
| **PRS: MEDS_PMID3** | 0.9503702673 | 0.6615677729 |
| **FLT4 deletion** | 0.8243292176 | 0.6632966159 |
| **PRS: PRSWEB_PHE** | 0.9518223736 | 0.6644948617 |
| **LINC00894 amplifica** | 0.3369511917 | 0.6656455848 |
| **DKC1 deletion** | 3.048493258 | 0.6670356018 |
| **DNMT3A amplificati** | 0.7576889491 | 0.6670749907 |
| **SETBP1 deletion** | 1.153361773 | 0.670982803 |
| **SNV in KMT2D** | 1.288485136 | 0.6711507356 |
| **JAK3 deletion** | 1.286776908 | 0.6712570922 |
| **PRS: ProstateCance** | 1.052283388 | 0.6714345861 |
| **TMEM127 deletion** | 0.2237694801 | 0.6718253888 |
| **ERCC6 deletion** | 0.8247096207 | 0.6719805155 |
| **FLCN amplification** | 0.04694641351 | 0.6721174424 |
| **PMS1 amplification** | 0.7917347025 | 0.6728555214 |
| **GNA11 deletion** | 0.7659437818 | 0.6734659829 |
| **PRS: PMID2786325** | 1.050506819 | 0.675306195 |
| **SNV in ARID1B** | 2.129565057 | 0.6761470761 |
| **BCL11B amplificatio** | 0.004864418212 | 0.6774663204 |

| | | |
|---|---|---|
| **BRD4 amplification** | 1.154376841 | 0.6778628545 |
| **EXT2 amplification** | 0.6920793328 | 0.6780104179 |
| **SMARCA4 deletion** | 1.25707504 | 0.6782486948 |
| **WHSC1L1 amplifica** | 0.7510682811 | 0.6791833483 |
| **ACVR1 deletion** | 0.3834228359 | 0.6800198988 |
| **CDKN2A deletion** | 0.9126743649 | 0.6807895337 |
| **Lab value: PTT** | 1.022739029 | 0.6810327876 |
| **HIST1H3C deletion** | 1.344432898 | 0.6813074369 |
| **XPA amplification** | 0.7420688765 | 0.6815074374 |
| **AURKB deletion** | 1.11472416 | 0.6821578402 |
| **MYCN amplification** | 0.7618841481 | 0.683209428 |
| **PDGFRA amplificati** | 0.8198535838 | 0.6836534877 |
| **UIMC1 amplificatior** | 1.25182875 | 0.6844844436 |
| **SNV in STK11** | 1.295283482 | 0.6850582892 |
| **PRS: PRSWEB_PHE** | 0.9609630792 | 0.6860253773 |
| **SNV in TP53** | 0.8855354241 | 0.6865909235 |
| **GEN1 deletion** | 0.2794955832 | 0.6876137644 |
| **SERPINA1 amplifica** | 0.3841481823 | 0.6896587117 |
| **PRS: UKB_460K.pig** | 0.9516588807 | 0.6899628246 |
| **CIITA deletion** | 0.7810141647 | 0.6925428379 |
| **KEAP1 deletion** | 0.7141079399 | 0.6928128401 |
| **PRS: TRICL.ade_s.H** | 0.9542215169 | 0.6930176275 |
| **MLH3 amplification** | 1.670753318 | 0.6941790145 |
| **RAD52 amplificatior** | 0.7706634452 | 0.6954413232 |
| **ID4 amplification** | 0.8438287415 | 0.6970021379 |
| **HELQ deletion** | 1.271790105 | 0.6972815555 |
| **PRS: PMID2806790** | 0.9375001177 | 0.698037036 |
| **PRS: PMID2806790** | 0.9375001177 | 0.698037036 |
| **PRS: PMID2806790** | 0.9375001177 | 0.698037036 |
| **TCF3 amplification** | 1.154645396 | 0.698815139 |
| **FANCE deletion** | 0.7537678646 | 0.6989583015 |
| **PRS: UKB_460K.dis** | 1.044975652 | 0.699382249 |
| **PRS: MEDS_PMID3** | 1.042946878 | 0.69975299 |
| **BABAM1 amplificati** | 0.06283391301 | 0.7000429234 |
| **SRC deletion** | 3211.65304 | 0.7026017386 |
| **NPRL3 amplificatior** | 0.2465269501 | 0.7030356507 |
| **HIST1H3B deletion** | 1.33888702 | 0.7036132023 |
| **CREBBP amplificati** | 1.243899388 | 0.7058989198 |
| **RBM10 amplificatio** | 0.2240182886 | 0.7061676285 |
| **PRS: TRICL.sma_s.l** | 1.044963443 | 0.7066468788 |
| **HRAS amplification** | 1.307184252 | 0.7099126796 |
| **Lab value: ALYMPH** | 1.063676091 | 0.710564752 |
| **Lab value: AEOS** | 1.385912906 | 0.7125144526 |

| | | |
|---|---|---|
| **Lab value: URIC** | 0.9004774116 | 0.7137374941 |
| **KDM5A amplificatio** | 0.7811970939 | 0.7144143469 |
| **USP8 amplification** | 0.536963194 | 0.7153369179 |
| **PRS: GSCAN.Cigare** | 0.9603401247 | 0.7154519392 |
| **VEGFA deletion** | 1.331224453 | 0.716039656 |
| **CTCF deletion** | 0.8222708059 | 0.7168969272 |
| **WT1 deletion** | 1.194158023 | 0.718865296 |
| **RHOA amplification** | 0.5867622208 | 0.7208358467 |
| **FLT1 amplification** | 0.7213293314 | 0.7209644275 |
| **KDR amplification** | 0.8372063149 | 0.721113837 |
| **SNV in PRKDC** | 1.219729527 | 0.7218359396 |
| **RNF8 deletion** | 1.33583999 | 0.7220494406 |
| **SOCS1 amplificatio** | 1.224581189 | 0.7221183483 |
| **GNB2L1 deletion** | 1.225746476 | 0.7222252183 |
| **PRS: UKB_460K.blo** | 1.03226687 | 0.7231884638 |
| **Lab value: UPH** | 1.114338516 | 0.7233351249 |
| **RAD50 amplificatio** | 1.292170478 | 0.7235129125 |
| **PRS: PRSWEB_PHE** | 1.042148243 | 0.7241807996 |
| **JAK2 amplification** | 0.8617751235 | 0.724189751 |
| **FANCC amplificatio** | 0.7932402968 | 0.7244112742 |
| **RAC1 deletion** | 0.2901411207 | 0.7247421576 |
| **PIK3R1 deletion** | 1.130307537 | 0.7265721124 |
| **BCL2L1 amplificatic** | 1.121637586 | 0.7283363985 |
| **HLA_A_3201** | 1.177433997 | 0.7284706704 |
| **MUTYH deletion** | 0.8591873735 | 0.7290501547 |
| **Lab value: SQUAM** | 1.102033153 | 0.7291775201 |
| **CDKN1C deletion** | 0.8567196723 | 0.7295263478 |
| **Lab value: EOS** | 1.024214219 | 0.7310265939 |
| **ABCB11 amplificati** | 0.6589230184 | 0.7310619151 |
| **Lab value: PHOS** | 0.8311406374 | 0.7317363956 |
| **TSHR amplification** | 1.972827906 | 0.7324556065 |
| **HLA_A_0301** | 0.9262468066 | 0.7329761769 |
| **CTNNB1 amplificati** | 1.213719281 | 0.7378251478 |
| **BCL2L12 deletion** | 0.8560702434 | 0.7386647455 |
| **KCNIP1 amplificatic** | 1.31706179 | 0.7388373365 |
| **NRAS deletion** | 0.8652547688 | 0.7405484237 |
| **Lab value: FRT4** | 1.780479016 | 0.7410180065 |
| **NFKBIE deletion** | 1.298518641 | 0.7416984115 |
| **MYCN deletion** | 0.759519667 | 0.7452575621 |
| **XPC deletion** | 0.8735005062 | 0.746331173 |
| **Lab value: FRT3** | 0.7964095113 | 0.7471705187 |
| **DNMT3A deletion** | 0.7751389764 | 0.7497821758 |
| **GNA11 amplificatio** | 0.8887338265 | 0.751027718 |

| | | |
|---|---|---|
| **PIK3C2B amplificati** | 0.9212567589 | 0.7525092868 |
| **KMT2D deletion** | 0.7536753558 | 0.7536727773 |
| **LMO1 amplification** | 0.5904118965 | 0.7553863899 |
| **POLH deletion** | 1.278973793 | 0.7554206343 |
| **PRS: PRSWEB_PHE** | 0.9670779338 | 0.7559229143 |
| **EGFR amplification** | 0.9146065707 | 0.7566605721 |
| **CIITA amplification** | 1.123256704 | 0.7602264065 |
| **PRS: UKB_460K.blo** | 1.034378465 | 0.7609321143 |
| **HFE deletion** | 1.263876231 | 0.7631490579 |
| **IKZF1 deletion** | 1.278744385 | 0.7634026585 |
| **Lab value: PLCO2** | 1.014115876 | 0.7643091492 |
| **SNV in POLQ** | 1.68600881 | 0.7650683504 |
| **PRS: TRICL.all.HM3** | 1.036270307 | 0.7658563578 |
| **PRS: UKB_460K.oth** | 1.033992228 | 0.7663704859 |
| **Lab value: PNA** | 1.011881226 | 0.7666403903 |
| **HLA_C_0702** | 1.070911259 | 0.7670835421 |
| **SNV in COL7A1** | 1.623427521 | 0.7697495741 |
| **ETV6 deletion** | 1.127213122 | 0.7705220015 |
| **KAT6A amplificatior** | 0.8467717306 | 0.7709535027 |
| **PRS: MEDS_PMID3** | 1.031069377 | 0.7716958401 |
| **JAZF1 amplification** | 0.8951450483 | 0.7717601026 |
| **CDK1 amplification** | 0.0009070566823 | 0.7718593615 |
| **KRAS amplification** | 1.082230114 | 0.7733126534 |
| **CBFA2T3 deletion** | 0.8763763716 | 0.7772260771 |
| **MDM2 deletion** | 1.188180294 | 0.7781245686 |
| **PRKCI amplificatior** | 0.9224818759 | 0.7788111355 |
| **HLA_DPB1_0201** | 1.085159518 | 0.7815486517 |
| **HLA_B_4001** | 1.098570375 | 0.7826114107 |
| **FOXL2 deletion** | 0.8498883004 | 0.7851527751 |
| **CASP8 amplificatio** | 1.209774284 | 0.7863285838 |
| **PRS: UKB_460K.blo** | 1.033809738 | 0.7865076311 |
| **B2M deletion** | 0.9146839559 | 0.7874174506 |
| **AURKA amplificatio** | 1.082997683 | 0.7878114814 |
| **NEIL2 amplification** | 0.8138732241 | 0.7881837412 |
| **NBN deletion** | 0.7986906785 | 0.789526348 |
| **ERCC5 deletion** | 0.8887075397 | 0.789649342 |
| **CTLA4 amplificatior** | 0.8346163579 | 0.7899948324 |
| **RFWD2 deletion** | 1.455295927 | 0.790569485 |
| **MLH3 deletion** | 1.173435678 | 0.7921566288 |
| **BCL6 amplification** | 1.07650384 | 0.7927261047 |
| **ETV6 amplification** | 1.090748793 | 0.7928044278 |
| **PRS: PRSWEB_PHE** | 0.9709849928 | 0.7936519634 |
| **PRSS1 deletion** | 1.186748096 | 0.7952568224 |

| | | |
|---|---|---|
| U2AF1 deletion | 1.161155282 | 0.7961987205 |
| NPM1 deletion | 1.12149267 | 0.7963640868 |
| SNV in EP300 | 0.6747685896 | 0.7977906192 |
| OGG1 deletion | 1.222779221 | 0.79783618 |
| HLA_DQA1_0301 | 1.060678624 | 0.7981214044 |
| CCND2 deletion | 0.8971887999 | 0.7984005272 |
| Lab value: HCT | 1.00571482 | 0.7987651132 |
| SH2D1A deletion | 0.5243336547 | 0.7991119367 |
| ABCB11 deletion | 0.5710032208 | 0.8010108837 |
| FKBP9 amplificatior | 0.8781421825 | 0.8018780209 |
| GLI3 amplification | 0.8749251203 | 0.8024278671 |
| PIK3CA amplificatio | 0.93158975 | 0.8029398425 |
| POLB amplification | 0.8671041847 | 0.8051507106 |
| SNV in BRCA1 | 1.110440111 | 0.8053805931 |
| MECOM amplificati | 0.9357377738 | 0.8074712842 |
| STAT6 deletion | 1.151166944 | 0.8075271937 |
| CDK6 deletion | 0.7982994302 | 0.8094470451 |
| PRS: MEDS_PMID3 | 0.9712453767 | 0.8101408809 |
| PRS: MEDS_PMID3 | 1.025847127 | 0.8101920595 |
| LMO3 amplification | 1.090639994 | 0.8102139086 |
| MAP2K4 deletion | 1.068317438 | 0.8110900234 |
| PRS: PRSWEB_PHE | 0.9741595436 | 0.8119883527 |
| CRTC2 deletion | 1.602940656 | 0.8120800306 |
| ERG deletion | 1.213684033 | 0.8121897937 |
| RAF1 amplification | 1.118919705 | 0.8122122179 |
| ACVR1 amplificatior | 1.229111016 | 0.8127713964 |
| NRG1 deletion | 0.9101364037 | 0.8129745947 |
| Lab value: RETCP | 1.829705071 | 0.814489402 |
| PRS: PMID2786325 | 0.9705846162 | 0.814781847 |
| MYC deletion | 0.6760600359 | 0.8148164289 |
| RFWD2 amplificatio | 1.07501738 | 0.8154463049 |
| RHOH amplification | 1.225272086 | 0.8167458296 |
| Lab value: MCH | 1.010170846 | 0.8186534766 |
| ERCC3 deletion | 0.832612745 | 0.8209795257 |
| SDHA amplification | 1.081916424 | 0.82225106 |
| Lab value: TBILI | 0.9361569788 | 0.822768435 |
| PRS: UKB_460K.dis | 1.024860619 | 0.822943161 |
| CTNNA1 amplificati | 1.18261966 | 0.8245444123 |
| POLB deletion | 0.9063899223 | 0.8254342312 |
| REL amplification | 0.9059714185 | 0.8257026731 |
| PRS: UKB_460K.dis | 1.023225441 | 0.8260880137 |
| PRS: PRSWEB_PHE | 1.02612489 | 0.8271195622 |
| SERPINA1 deletion | 1.147698705 | 0.8279569912 |

| | | |
|---|---|---|
| **PRAME deletion** | 1.1358027 | 0.8286819105 |
| **PRS: PRSWEB_PHE** | 1.023231382 | 0.8292229184 |
| **PRS: PRSWEB_PHE** | 1.022409033 | 0.8293603064 |
| **CDKN2B deletion** | 0.9530909663 | 0.8312876928 |
| **TSC2 amplification** | 1.129639418 | 0.8314362934 |
| **CD274 amplificatior** | 1.078577475 | 0.8315159454 |
| **FAM46C deletion** | 1.094502507 | 0.8324043525 |
| **PRS: PMID27863252** | 0.9767397593 | 0.8336503845 |
| **PRS: UC_deLange2** | 1.023680353 | 0.8339470234 |
| **FGFR2 amplificatior** | 0.8445939477 | 0.8347988133 |
| **HELQ amplification** | 0.4589264876 | 0.8362505474 |
| **PPARG amplificatio** | 1.142344767 | 0.8374108158 |
| **CIC deletion** | 0.585136413 | 0.8375720232 |
| **SETD2 deletion** | 1.065609869 | 0.8411000272 |
| **PRS: Asthma.age_o** | 0.9766632843 | 0.841874855 |
| **PRS: PMID27863252** | 1.022948496 | 0.8418806835 |
| **SLC34A2 deletion** | 1.118750717 | 0.842183371 |
| **MTA1 deletion** | 1.135165829 | 0.8424067517 |
| **PRS: GSCAN.Smoki** | 1.021328075 | 0.8447288379 |
| **OGG1 amplification** | 0.8385040152 | 0.8448666823 |
| **KIT amplification** | 0.9196917123 | 0.8453269216 |
| **SDHA deletion** | 0.8912342783 | 0.8460030867 |
| **Lab value: MCHC** | 0.976061677 | 0.8460668845 |
| **DICER1 deletion** | 1.066390572 | 0.8467341975 |
| **PRS: UKB_460K.imp** | 0.9789487335 | 0.8479490512 |
| **FANCL amplificatior** | 0.9011988022 | 0.8480527711 |
| **Lab value: MAMON** | 1.409754077 | 0.8480608472 |
| **FOXA1 deletion** | 1.178205792 | 0.8483553745 |
| **RELA deletion** | 1.348990876 | 0.8499241248 |
| **PRS: MEDS_PMID3** | 0.9725226259 | 0.8502774654 |
| **PRS: UKB_460K.ca** | 0.9782582745 | 0.8503620313 |
| **ELANE amplificatior** | 0.6878713428 | 0.850555612 |
| **DICER1 amplificatio** | 1.123631076 | 0.8511850109 |
| **GNAS amplification** | 1.057692587 | 0.8514664635 |
| **HLA_C_0602** | 1.052179743 | 0.8518722001 |
| **NPM1 amplification** | 1.077401312 | 0.851998235 |
| **PRS: MEDS_PMID3** | 1.021092307 | 0.8524344085 |
| **HLA_A_6802** | 1.369006682 | 0.8533779039 |
| **MAX amplification** | 0.8677412208 | 0.853625169 |
| **PRS: UKB_460K.dis** | 0.9775080202 | 0.8545168319 |
| **CADM2 amplificatio** | 0.6414549286 | 0.8546550838 |
| **TRAF3 deletion** | 1.121966603 | 0.8552981074 |
| **RECQL4 deletion** | 0.6794739592 | 0.8554282464 |

| | | |
|---|---|---|
| CRTC1 amplificatio | 1.068625373 | 0.855900989 |
| PRS: UKB_460K.dis | 1.022015259 | 0.8559349979 |
| MLH1 deletion | 0.9381553603 | 0.8564322591 |
| RAD51 amplificatio | 1.296218774 | 0.8571125726 |
| TET1 amplification | 0.6641668475 | 0.8573197803 |
| FAS deletion | 0.9456473437 | 0.8575612224 |
| SBDS amplification | 0.9414353131 | 0.8581314583 |
| PTCH1 amplificatio | 0.8873606301 | 0.8583013815 |
| RHOT1 deletion | 1.177839824 | 0.8593048907 |
| FAM175A amplificat | 0.5006545553 | 0.8614629797 |
| GLI1 deletion | 1.102111319 | 0.8614774198 |
| HLA_A_6801 | 1.080708863 | 0.8620239844 |
| PRS: MEDS_PMID3 | 1.020016145 | 0.8625851102 |
| CDH4 amplification | 0.9138193963 | 0.8626849048 |
| PRS: UKB_460K.dis | 1.02187455 | 0.863393788 |
| GNAQ amplification | 0.8876018341 | 0.8640882666 |
| DIS3L2 amplificatio | 0.8526820852 | 0.8646509686 |
| PRS: UKB_460K.dis | 1.019316646 | 0.8651448179 |
| HLA_A_2601 | 1.086897663 | 0.8652625584 |
| PALB2 amplificatior | 0.8796065247 | 0.8661124616 |
| PTK2 deletion | 0.6810072686 | 0.8663291756 |
| KAT6B amplificatior | 1.157956781 | 0.8663664765 |
| FANCM amplificatio | 1.118554811 | 0.8666806243 |
| SMARCA4 amplifica | 1.060414717 | 0.8671163273 |
| KAT6A deletion | 0.9292588306 | 0.8686345337 |
| STK11 amplificatior | 0.9429098537 | 0.8687540855 |
| PIK3R1 amplificatio | 1.121106562 | 0.8704065623 |
| HLA_DRB1_0404 | 1.124991749 | 0.8720665595 |
| MUS81 deletion | 1.292039243 | 0.8720837655 |
| ARAF deletion | 0.6592663846 | 0.8742648112 |
| EXT1 deletion | 1.244611239 | 0.8758664195 |
| CDK5 amplification | 0.9343096315 | 0.8760113083 |
| ARID2 deletion | 0.9159036176 | 0.876060842 |
| PRS: PRSWEB_PHE | 0.979767746 | 0.8768666727 |
| ERCC2 amplificatio | 1.056650288 | 0.8804448185 |
| TMPRSS2 amplifica | 0.9137962617 | 0.8807886415 |
| DAXX deletion | 1.132465665 | 0.8811471502 |
| HLA_DQB1_0302 | 1.040485877 | 0.8828525726 |
| PRS: PMID2786325 | 0.9842105236 | 0.8846694223 |
| GREM1 deletion | 0.9352296985 | 0.8851730902 |
| CDH1 amplification | 1.073817231 | 0.8853653053 |
| ERCC4 amplificatio | 1.081642513 | 0.8857412982 |
| PRS: UKB_460K.me | 1.016919267 | 0.8857901548 |

| | | |
|---|---|---|
| **HLA_A_1101** | 0.944176739 | 0.8861528674 |
| **RAD21 deletion** | 0.7627852566 | 0.8878771864 |
| **PRS: UKB_460K.lun** | 1.017045761 | 0.889781711 |
| **DCLRE1C amplifica** | 1.122758262 | 0.8898736935 |
| **IKZF1 amplification** | 0.9580918318 | 0.8900404374 |
| **MPL deletion** | 1.060891152 | 0.8914886167 |
| **PRS: UKB_460K.rep** | 1.018078208 | 0.8923595428 |
| **PRS: UKB_460K.pig** | 1.016501719 | 0.8955980173 |
| **SOS1 amplification** | 0.9132156477 | 0.8963083235 |
| **FANCM deletion** | 1.114908499 | 0.8963466733 |
| **HLA_B_1302** | 0.9205037981 | 0.8966658671 |
| **RHPN2 amplificatio** | 0.9404668167 | 0.8967941771 |
| **BAP1 deletion** | 1.042145599 | 0.8980491607 |
| **FGFR3 amplificatior** | 0.9378618459 | 0.8984243793 |
| **PRS: Stratified_fem** | 0.9841052464 | 0.8989845937 |
| **CXCR4 deletion** | 0.8166517216 | 0.8993023653 |
| **GATA2 amplificatior** | 1.080101623 | 0.9000427566 |
| **TET2 amplification** | 1.117142207 | 0.9000695078 |
| **PRS: UKB_460K.bo** | 0.9852123008 | 0.9011296014 |
| **Lab value: PCL** | 1.004138976 | 0.9041233345 |
| **DOCK8 deletion** | 0.9586818551 | 0.9048523751 |
| **GATA4 deletion** | 0.9692712284 | 0.906660159 |
| **Lab value: USPG** | 49.89897742 | 0.9100286331 |
| **TP53 deletion** | 1.030132137 | 0.9105712917 |
| **MGA amplification** | 1.176580437 | 0.9111489718 |
| **TCF7L2 amplificatio** | 0.909634789 | 0.9119864744 |
| **TMPRSS2 deletion** | 1.100669571 | 0.9124427083 |
| **PRS: MEDS_PMID3** | 1.013905816 | 0.9128909341 |
| **PRS: TRICL.sqc_s.H** | 1.01549496 | 0.9132008442 |
| **CDK8 amplification** | 0.8852453463 | 0.9139687841 |
| **WHSC1L1 deletion** | 0.9552579876 | 0.9141132813 |
| **PRS: PMID2786325** | 0.9886818225 | 0.9151607652 |
| **AXIN2 deletion** | 1.103871686 | 0.9171141415 |
| **PRS: PRSWEB_PHE** | 1.01109454 | 0.9177487393 |
| **PRS: Asthma.childr** | 0.9866831645 | 0.9178051725 |
| **HLA_DQB1_0402** | 0.8822900064 | 0.9182607996 |
| **PRS: UKB_460K.bo** | 1.0109969 | 0.9186430515 |
| **PRS: UKB_460K.rep** | 1.01255359 | 0.9187849823 |
| **CARD11 amplificati** | 1.026882206 | 0.920559039 |
| **RINT1 deletion** | 1.109003559 | 0.9210333 |
| **PML amplification** | 1.111524354 | 0.9218438752 |
| **CXCR4 amplificatio** | 0.7977341388 | 0.9225179943 |
| **Lab value: HGB** | 1.005999533 | 0.9232369657 |

| | | |
|---|---|---|
| ERBB3 deletion | 0.9454188693 | 0.9236397072 |
| TFE3 amplification | 1.212860534 | 0.9242402103 |
| Lab value: ANION | 1.006630782 | 0.9244927867 |
| PRS: PRSWEB_PHE | 0.9902563101 | 0.9245261871 |
| PRS: UKB_460K.pig | 0.988785433 | 0.9251329382 |
| MYD88 deletion | 0.9678321507 | 0.9258786039 |
| Lab value: RDW | 0.9956179122 | 0.9275528667 |
| Lab value: RBC | 1.016444942 | 0.9276385994 |
| PTK2B deletion | 1.035127667 | 0.927737634 |
| PNRC1 amplificatio | 0.9421724306 | 0.9277756897 |
| XRCC3 deletion | 1.058000615 | 0.9283064766 |
| XPC amplification | 1.044343031 | 0.928380255 |
| Lab value: WBC | 1.003233158 | 0.9292914297 |
| EP300 deletion | 1.04059046 | 0.9293436358 |
| PRS: PRSWEB_PHE | 0.9901162172 | 0.929489311 |
| PRS: X25HydroxyVit | 0.9867066137 | 0.929634423 |
| HLA_B_2705 | 1.050157231 | 0.9302007918 |
| TERT deletion | 0.9601511187 | 0.9312546804 |
| FAN1 deletion | 1.041533464 | 0.9312626473 |
| HLA_A_2501 | 1.064971934 | 0.9312940204 |
| RSPO2 deletion | 0.7788751325 | 0.9316568253 |
| FUS amplification | 0.9533883127 | 0.9330640406 |
| XRCC1 deletion | 0.8721804247 | 0.9346384082 |
| FBXW7 deletion | 1.03024272 | 0.9350180911 |
| MAFB amplification | 1.039796409 | 0.9368976229 |
| LMO2 deletion | 0.9602970199 | 0.9386807464 |
| PTK2B amplificatior | 1.060524203 | 0.9387134098 |
| HLA_B_5501 | 0.9529822168 | 0.9388820296 |
| CASP8 deletion | 1.186006329 | 0.939147981 |
| PRS: UKB_460K.dis | 0.991839716 | 0.9410934955 |
| ETV5 amplification | 0.9805125387 | 0.9425076061 |
| CDKN2C deletion | 0.9722553525 | 0.9431604443 |
| PIM1 deletion | 1.050298212 | 0.9433692363 |
| PRS: MEDS_PMID3 | 1.007959765 | 0.9434872195 |
| PRS: PMID2786325 | 0.9928799005 | 0.9443774613 |
| MCM8 amplificatior | 0.9523068509 | 0.9446426599 |
| PRS: PRSWEB_PHE | 0.9928907689 | 0.9455508489 |
| WAS amplification | 1.143779377 | 0.9461718757 |
| NOTCH2 deletion | 1.0327587 | 0.9462624285 |
| BRCC3 deletion | 0.8479509128 | 0.9472904566 |
| PRS: PRSWEB_PHE | 1.007008544 | 0.9475741129 |
| GALNT12 amplificat | 1.052871444 | 0.9484675007 |
| ERCC3 amplificatio | 0.9465802652 | 0.9490746485 |

| | | |
|---|---|---|
| **ERBB4 amplificatio** | 1.046139216 | 0.9498103085 |
| **CDKN1A deletion** | 0.9572393461 | 0.9511647825 |
| **ROS1 amplification** | 1.037330267 | 0.952085043 |
| **TP53BP1 amplificat** | 0.9086419426 | 0.953019076 |
| **HLA_DPB1_0401** | 1.01102774 | 0.9533150429 |
| **HLA_B_3801** | 0.9657659762 | 0.9534384333 |
| **LIG4 deletion** | 1.039169356 | 0.9545427461 |
| **FGFR4 deletion** | 1.022251565 | 0.9558034848 |
| **CRKL deletion** | 1.027761725 | 0.9561657938 |
| **HLA_C_1203** | 1.017811158 | 0.9572343143 |
| **DDB1 deletion** | 0.9017106886 | 0.9584762643 |
| **PRS: TRICL.sqc_ns.** | 1.006422957 | 0.9588598321 |
| **PRS: PRSWEB_PHE** | 1.005413486 | 0.9609496517 |
| **TFE3 deletion** | 1.236337867 | 0.961256813 |
| **WAS deletion** | 1.236337867 | 0.961256813 |
| **ERCC5 amplificatio** | 0.9794591705 | 0.9622038461 |
| **JAK3 amplification** | 0.983489919 | 0.9623304233 |
| **SQSTM1 amplificati** | 0.9727597252 | 0.9626838868 |
| **CD79B deletion** | 1.040727525 | 0.963176758 |
| **PRS: UKB_460K.dis** | 0.9937640441 | 0.9634619015 |
| **PRS: MEDS_PMID3** | 1.005431894 | 0.9634964093 |
| **FGFR3 deletion** | 0.9797652367 | 0.9642436164 |
| **NEIL2 deletion** | 0.9833653187 | 0.9653591837 |
| **SNV in MGA** | 0.8810024004 | 0.9660283411 |
| **PRS: PMID2786325** | 0.9957154121 | 0.9660345909 |
| **PRS: TRICL.ade1_s.** | 1.005172127 | 0.9662163906 |
| **BRD4 deletion** | 1.028261243 | 0.9679296962 |
| **Lab value: DMANUT** | 0.9950376721 | 0.9679735419 |
| **PRS: PMID2786325** | 0.9949741607 | 0.9691883538 |
| **POLD1 amplificatio** | 0.9695343327 | 0.9706371056 |
| **AKT1 deletion** | 0.9878851725 | 0.9724581222 |
| **FANCA amplificatio** | 0.9757392811 | 0.9735522011 |
| **PBRM1 deletion** | 0.9890473518 | 0.9738792719 |
| **Lab value: ANEUT** | 0.9984341336 | 0.9740599929 |
| **QKI deletion** | 1.009683417 | 0.9747382126 |
| **BCL11B deletion** | 1.019066003 | 0.9761271058 |
| **PRS: GSCAN.Smoki** | 0.9968347461 | 0.9769677884 |
| **NRG1 amplification** | 0.9783910664 | 0.9780479073 |
| **STK11 deletion** | 1.013344112 | 0.9785940132 |
| **DIS3 deletion** | 0.9896211661 | 0.9790869234 |
| **DDB2 deletion** | 0.9871682522 | 0.9794243105 |
| **PRS: PRSWEB_PHE** | 0.9972161363 | 0.9796718144 |
| **CCND3 deletion** | 0.9801229889 | 0.9799068865 |

| | | |
|---|---|---|
| **Lab value: PK** | 1.008492781 | 0.9802235491 |
| **SNV in CREBBP** | 1.01987343 | 0.9816829083 |
| **PRS: UKB_460K.blo** | 0.997840337 | 0.9831493676 |
| **PRS: UKB_460K.cov** | 0.9972730873 | 0.9834955395 |
| **PRS: TRICL.smoker** | 0.9977510042 | 0.9843790908 |
| **PRS: PRSWEB_PHE** | 0.9981591882 | 0.9864028363 |
| **RBM10 deletion** | 1.077440162 | 0.9864123903 |
| **HLA_A_2301** | 1.005499337 | 0.9899979977 |
| **PRS: PRSWEB_PHE** | 0.9986267771 | 0.990026711 |
| **CTLA4 deletion** | 1.029057733 | 0.9901551282 |
| **PRS: UKB_460K.dis** | 0.9988039266 | 0.9912128252 |
| **PHOX2B deletion** | 1.004834881 | 0.9916671588 |
| **CBLB amplification** | 1.003719273 | 0.9920268895 |
| **CTNNB1 deletion** | 0.9969669751 | 0.992889449 |
| **TAZ deletion** | 1.021980304 | 0.9929292481 |
| **NTRK1 deletion** | 1.011562655 | 0.9930403156 |
| **PRS: UKB_460K.blo** | 0.9991668495 | 0.9944127287 |
| **PRS: PRSWEB_PHE** | 0.9992649344 | 0.9946009711 |
| **SLC34A2 amplificat** | 1.005635564 | 0.9950440776 |
| **EPHA5 deletion** | 1.002364646 | 0.9958365143 |
| **ASXL1 amplificatior** | 1.001288142 | 0.9969929254 |
| **AKT2 amplification** | 1.000996708 | 0.9976540872 |
| **SLC25A13 deletion** | 1.002809485 | 0.9978765417 |
| **Lab value: LIPASE** | 1.000025315 | 0.9985065475 |
| **MED12 amplificatio** | 0.7247086996 | 0.9992168184 |
| **LINC00894 deletion** | 0.3793687701 | 0.9994770059 |
| **SNV in STAG2** | 0.4685790378 | 0.9998049289 |
| **CSF1R deletion** | 0.9998719684 | 0.9998238112 |
| **AURKB amplificatio** | 0.6827227119 | 0.9998480575 |
| **GBA deletion** | 0.52536077 | 0.9998524322 |
| **AR amplification** | 0.8188294093 | 0.9999173731 |
| **RIT1 deletion** | 0.1809994975 | 0.9999200459 |
| **PVRL4 deletion** | 0.2007445671 | 0.9999781239 |

| **Search terms** | **Restriction of results** |
|---|---|
| due to antineoplastic therapy | glucocorticoids and synthetic analogues |
| allergic reaction to drug | anemia |
| adverse effect | hepatitis c |
| toxic | immunosuppressive |
| autoimmune | infectious |
| drug-induced | anticoagulants |
| hepatitis | due to inhalation of food and vomit |
| endocrinopathy | insomnia |
| nephritis | constipation |
| pancreatitis | monitoring |
| uveitis | diagnostic agents |
| muscolosceletal | opium |
| carditis | opioid |
| cystitis | pancytopenia |
| athritis | without complication |
| adrenal insufficiency | leukopenia |
| mucositis | agranulocytosis |
| myositis | clostridium difficile |
| pruritis | without esophagitis |
| sialadenitis | fever |
| bronchiolitis | non-steroid |
| myocarditis | malignant |
| ketoacidosis | nontoxic |
| colitis | chemo |
| pneumonitis | radi |
| dermatitis | chronic |
| neuropathy | cancer |
| hypothyroidism due to medication | nodule |
| hypothyroidism due to medicamen | history |
| | contact |
| | melanoma |
| | infection |
| | test |
| | cyst |
| | antibody |
| | viral |
| | bacteria |
| | noninflammatory |
